# Social network and household exposure explain the use of malaria prevention measures in rural communities of Meghalaya, India

**DOI:** 10.1101/2023.04.23.23288997

**Authors:** Elisa Bellotti, Andras Voros, Mattimi Passah, Quinnie Doreen Nongrum, Carinthia Balabet Nengnong, Charishma Khongwir, Annemieke van Eijk, Anne Kessler, Rajiv Sarkar, Jane M. Carlton, Sandra Albert

**Affiliations:** Department of Sociology, University of Manchester, Manchester, UK; School of Social Policy, University of Birmingham, Birmingham, UK; Center for Genomics and Systems Biology, Department of Biology, New York University, USA; Indian Institute of Public Health Shillong, Shillong, Meghalaya, India

## Abstract

Malaria remains a global concern despite substantial reduction in incidence over the past twenty years. Public health interventions to increase the uptake of preventive measures have contributed to this decline but their impact has not been uniform. To date, we know little about what determines the use of preventive measures in rural, hard-to-reach populations, which are crucial contexts for malaria eradication. We collected detailed interview data on the use of malaria preventive measures, health-related discussion networks, individual characteristics, and household composition in ten tribal, malaria-endemic villages in Meghalaya, India in 2020-2021 (n=1,530). Employing standard and network statistical models, we found that social network and household exposure were consistently positively associated with preventive measure use across villages. Network and household exposure were also the most important factors explaining behaviour, outweighing individual characteristics, opinion leaders, and network size. These results suggest that real-life data on social networks and household composition should be considered in studies of health-behaviour change.

## INTRODUCTION

Following a substantial reduction in the global incidence of malaria between 2001 and 2015, the World Health Organization set a goal of global eradication by 2030 [WHO, 2015]. While large-scale interventions played a pivotal role in controlling and eliminating malaria in some countries, their impact has varied across the malarious world, resulting in little change for poor, migrant, and hard-to-reach populations [WHO, 2015]. Prevention in residual epicentres is challenging unless local populations are engaged [Dhiman, 2019]. Treating difficult pockets of transmission requires tailored and targeted approaches suited to local contexts [Gosling et al., 2020]. This is true for Meghalaya state in Northeastern India, a hilly and mountainous area of subtropical rain forests interspersed with rice-agroecosystems. The wet climate makes Meghalaya an optimal setting for malaria, with multiple *Anopheles* mosquito species transmitting *Plasmodium falciparum* and *Plasmodium vivax* malaria parasites [Kessler et al., 2018; Kessler et al., 2021]. Meghalaya is inhabited mostly by tribal peoples, with 86% of its population of approximately 3.2 million belonging to an indigenous ethnicity or Scheduled

Tribes as referred to by the government of India [Rajagopal, 1976; Lele et al., 2009; Ministry of Tribal Affairs, 2021]. ‘Tribals’ often experience geographical marginalization, poor access to health care, and low socio-economic status [Rajagopal, 1976; Sharma et al. 2015; Kessler et al., 2018], environmental and social factors that pose a challenge to malaria prevention efforts. In Meghalaya, large-scale malaria prevention efforts are coordinated by modern, government-run public health programmes managed through local Primary Health Centres (PHC) and facilitated at the village level by Accredited Social Health Activists (ASHAs). These operate alongside largely undocumented traditional tribal treatments provided by Traditional Healers [Albert et al., 2015].

Large-scale malaria prevention measures in Meghalaya have mainly focused on reducing mosquito bites [Debboun et al., 2013] through the use of Indoor Residual Spraying (IRS) and the distribution of Long-Lasting Insecticidal Nets (LLINs), which repel and reduce the longevity of indoor-feeding and resting of *Anopheles* vector mosquitoes [Mishra et al., 2021]. Both methods have been associated with a substantial reduction in malaria incidence [Sahu et al., 2020] but are used only indoors [Lek et al., 2020; Sangoro, 2015; Rubio-Palis et al., 1992]. Villagers working or socializing outside their dwellings, especially in the evening, night, and early morning, may use other measures to prevent mosquito bites [Sangoro, 2015; Wilson et al., 2014], such as protective clothing, insect repellent spray or body cream. IRS is contested in many Meghalayan villages [Sarkar et al., 2021; Passah et al., 2022], where households could turn to other indoor measures such as air-borne repellents (coils and vaporizers) instead.

Studies that measure the adoption of – or resistance to – mosquito bite prevention methods usually evaluate how characteristics of individuals or households (gender, education, socio-economic background, number of children, living conditions, etc.) relate to knowledge and practice of malaria preventive and treatment behaviours [Nlinwe et al., 2021; Matin et al., 2020; Van Eijk et al., 2016; Shahandeh et al, 2012]. What these studies do not consider is that information and attitudes regarding both effective and ineffective preventive behaviours are likely to spread through social networks. These may facilitate support for but also resistance to new practices. Such often-overlooked network processes can help explain why some effective practices, like the use of vaporisers, may become common in certain areas [Van Eijk et al., 2016] while others, like IRS, may be opposed [Sarkar et al., 2021; Passah et al., 2022], or why practices whose effectiveness is unproven, like burning egg trays and jute bags, may thrive.

Social network research is increasingly becoming common in the study of adoption of health behaviours, including eating habits [Salvy et al., 2012], physical activity [Proestakis et al., 2018], family planning [Gayen et al., 2010; Stoebenau et al., 2003], healthy practices [Kim et al., 2015], and general diffusion of information and innovations [Rogers Everett, 1962; Coleman et al., 1957; Valente, 1996; Valente 2010; Centola; 2010; Centola, 2018]. Threshold models postulate that individuals adopt innovative behaviours depending on the proportion of the population already engaged in such behaviours [Granovetter, 1978]. They further propose that individuals comfortable with a low threshold adopt a behaviour earlier than others, while late adopters require a higher threshold. Threshold models are efficient in predicting simple contagion, for which proximity – being close to infected subjects – is enough to trigger diffusion [Watts et al., 1998; Centola, 2010]. However, behaviours are complex phenomena that do not spread across networks in the same way an airborne disease might [Centola and Macy, 2007]. Adopting a behaviour like burning coils or jute bags is influenced both by locals who use these preventive measures as well as those who do not use them because they consider them ineffective or explicitly oppose them [Centola; 2010; Centola, 2018]. To be diffused, innovations need to be adopted and reinforced within local neighbourhoods of social networks [Coleman et al., 1957; Valente, 1996; Valente 2010; Centola; 2010; Centola, 2018].

A network exposure model of diffusion [Coleman et al., 1957; Valente, 1996; Valente 2010; Centola, 2018] does not measure the threshold level of the entire population but rather, the threshold level of the proportion of adopters within the population that individuals are directly related to. It postulates that innovative behaviour spreads relatively quickly within highly clustered and homophilous regions of the networks [Christakis and Fowler, 2007; Smith et al., 2008; Crosnoe et al., 2008; Valente et al., 2009] but the spreading slows down where such clusters are disconnected [Centola, 2011]. Central actors such as persons with numerous contacts [Freeman 1979] may not be the best located actors to promote diffusion of new practices. They are usually considered opinion leaders and champions of the status quo who are reluctant to change the way things are done within a community [Valente and Pumpang 2007]. Innovations are more likely to spin off from peripheries of social networks, where people are less constrained by collective social norms, are more likely to be connected through non-redundant weak ties, and therefore are potentially more exposed to norms of different groups [Granovetter 1978, Burt 2004].

Most studies that model diffusion of diseases or information use secondary [Weston et al., 2018] or synthetic data [Horsevad et al., 2022] to calibrate or validate models [Verelst et al., 2016; Bedson et al. 2021]. Social network information is at best inferred from physical proximity [Levy et al., 2021]. Studies that collect primary social network data, especially from remote, hard-to-reach tribal villages, are very rare [Gayen et al., 2010; Stoebenau et al., 2003; Kim et al. 2015; Holly et al., 2014; Koster, 2011]. This is so because social network data need to be gathered from the entire population of interest [Smith et al., 2008; Perkins et al., 2015] with minimal missing data, which is a huge challenge in hard-to-reach populations where individuals spend most of the day working outdoors. To guarantee coverage, the population of interest must be identified and bounded [Smith et al., 2008; Marsden, 2002], while acknowledging that boundaries are always arbitrary. In practice, an individual’s social network extends beyond the village they live in, even in remote areas [Stoebenau and Valente 2003; Koster 2011].

Once populations have been identified and bounded, and data have been collected, social network analysis can be used to study the links between the structure of networks and the behaviours of its actors. This requires specialised statistical techniques [Steglich et al., 2010], since dependencies in networks and between networks and behaviour violate the independence assumption at the core of most standard (non-network) statistical methods [Bartels et al., 1979; Doreian 1980; Dow et al., 1983; Dow et al., 1984; Ord 1975; Leenders 2002]. The Stochastic Actor-oriented Model (SAOM) is one of the few methods suitable to analyse the dynamics of social networks and individual behaviour [Snijders et al., 2010]. The SAOM is increasingly popular in the social sciences and has been applied in the context of various health behaviours in recent years [adams and Schaefer, 2017; de la Haye et al., 2019; Long and Valente, 2019; Franken et al., 2023]. Despite successful applications of network models, empirical data in the area of disease prevention, and specifically malaria prevention, are still predominantly analysed with non-network methods [Nlinwe et al., 2021; Matin et al., 2020; Van Eijk et al., 2016; see Musoke et al. 2023 for a review].

In this study, we assess the importance of social networks in explaining the use of eight mosquito bite preventive measures in ten hard-to-reach tribal villages in northeast India. We compare the role of five groups of factors affecting villagers’ use of the preventive measures: individual characteristics, influence from village opinion leaders, social network size (within and outside villages), network exposure, and household exposure. We first examine which factors explain the use of each preventive measure by fitting village-level logistic regression models. In the results, we present the meta-analysis of all models (Snijders et al. 2012), which shows the significant variables, across villages, in explaining the use of each preventive measure. We then apply stationary Stochastic Actor-oriented Models [Snijders and Steglich, 2015], an extension of SAOMs for cross-sectional data observed at a single time point, to jointly model individuals’ use of preventive measures and their social relations with other villagers. In this step, we consider villagers’ use of any measure, treating the system of social ties and measure use as a multilevel network (Lazega and Snijders, 2015:36-37]. While our data is cross-sectional, the SAOM is dynamic in nature and allows inference to social processes that can explain the observed patterns of social ties, preventive behaviour, individual attributes, and household structure (see the Methods section for further details).

To our knowledge, this is the first study where a social network analysis approach is used in the context of mosquito bite prevention. To date, stationary SAOMs have not been fitted to multilevel network data in the scholarly literature, while we know of only two applications of the method to a single network, both outside of the area of health-related behaviours [Snijders and Steglich, 2015; Simpson 2022]. Similar to the logistic regression models, in the results we present the meta-analysis of all SAOM models. Details of the meta-analysis procedure can be found in the Method section, while full logistic and SAOM models results are reported in the Supplementary Information, Tables 12-19 and Appendix E, Tables 41-44.

We apply the above models to data we collected in the state of Meghalaya in India between January 2020 and August 2021. Data is available from ten villages in three districts: three villages in West Khasi Hills (WK1, WK2, WK3), three villages in West Jaintia Hills (WJ1, WJ2, WJ3), and four villages in South Garo Hills (SG1, SG2, SG3, SG4). In these areas, detailed malaria epidemiological and behavioural information has only recently been generated [Kessler et al., 2018; Kessler et al., 2021]. See Figure 1 for the geographic context of our study sites.

**Figure 1.**
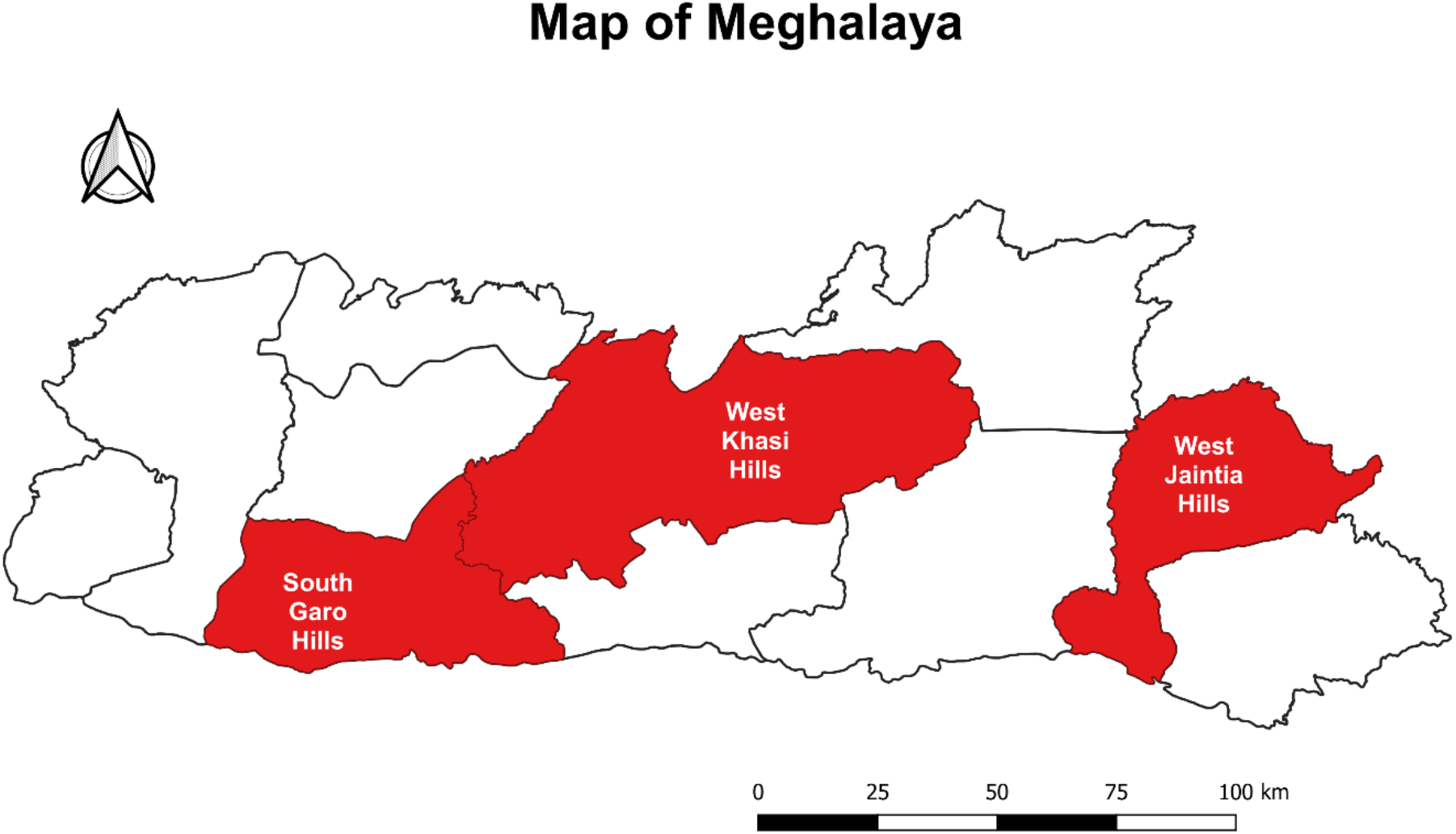
The map of Meghalaya state in Northeast India with the three studied districts highlighted.

A structured questionnaire was administered to every reachable adult (≥18 years old) in the studied villages, resulting in information from 1,530 villagers residing in 766 households.

From each participant, we collected information about: (1) use of mosquito bite preventive measures (LLINs, covering clothes, boots, gloves, insecticide cream, coils, vaporizers, burning materials), (2) demographic characteristics and role(s) within household and village, (3) health-related discussion network within and outside village, and (4) household membership. The ASHA was interviewed in all 10 villages, and in the six villages where present, the Traditional Healer was also interviewed. Both the ASHA and the Traditional Healer are considered opinion leaders for their health-related expertise and their role in administering healthcare within villages. The full list of questions, details on coding and variable descriptives are reported in the Supplementary Information, Appendix B, Tables B1-B15.

By interviewing most adults in each village, we can quantify how many of the people a villager talks to about health-related matters use a given preventive measure – we refer to this as ***network exposure***. Similarly, the number of household members using a preventive measure defines a villager’s ***household exposure*** to this measure. **Figure 2** provides an illustrative example for network exposure to the use of coils, based on data from one of the villages; household exposure may be conceived similarly. We use this conceptualisation of exposure in the first part of our analysis, as it is compatible with standard logistic regression models. Figure 3 provides a different view on exposure that allows to consider all preventive measures at once: for the same village, it presents measure use and discussion ties as a ***multilevel network***. In this network, ties representing usage connect villagers to any measure they report using and discussion ties connect villagers to each other. We use this conceptualisation in the second part of our analysis, as it allows to study patterns of use and discussion across measures in SAOMs.

**Figure 2.**
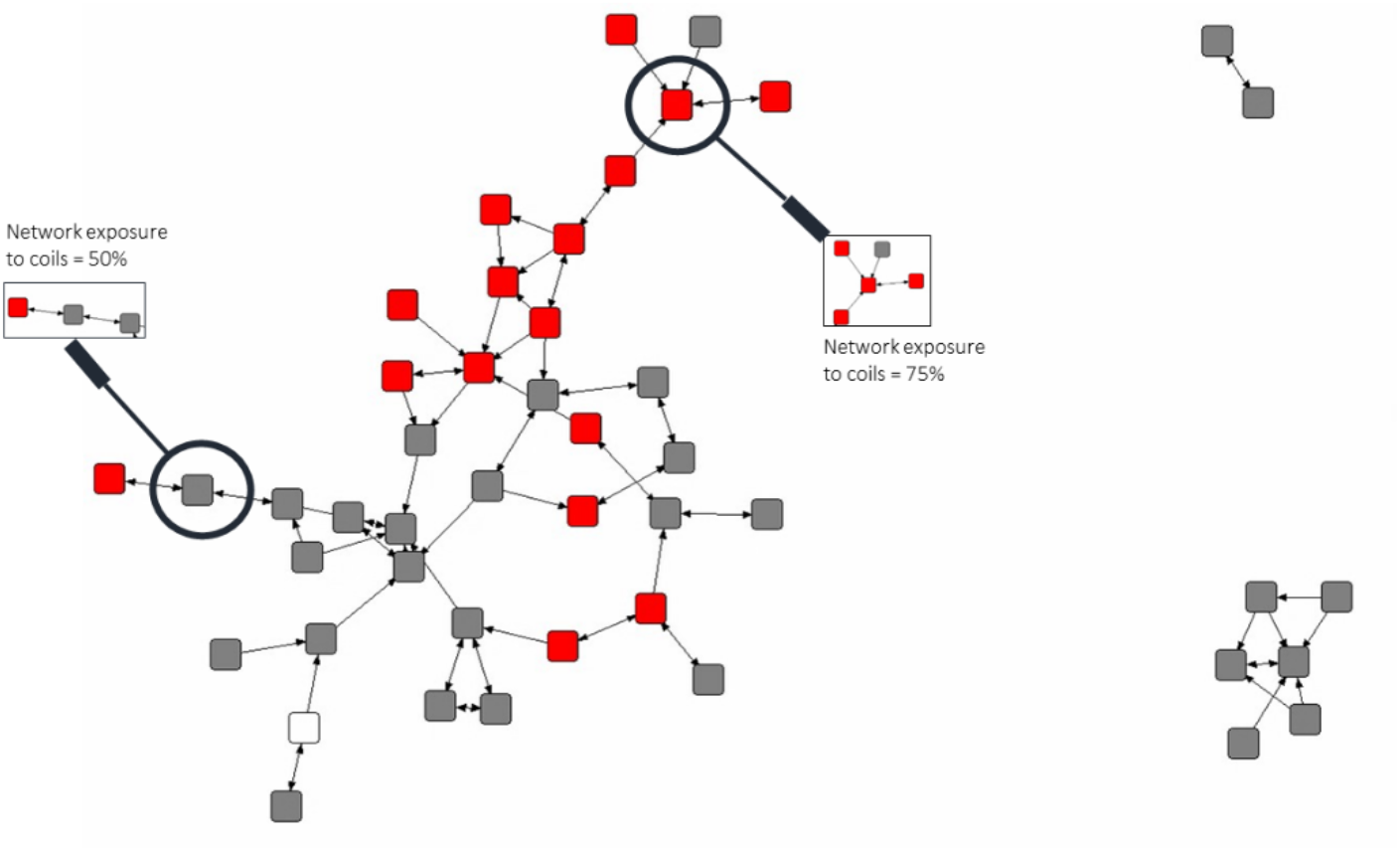
Example of percentage of exposure to the use of coils in WK2. Nodes represent interviewed villagers; lines represent whom the villagers talk to about health-related matters; red nodes indicate villagers who use coils; grey nodes indicate villagers who do not use coils. Network exposure to coils of the node highlighted at the top right = 75%. Network exposure to coils of the node highlighted at the left = 50%.

**Figure 3.**
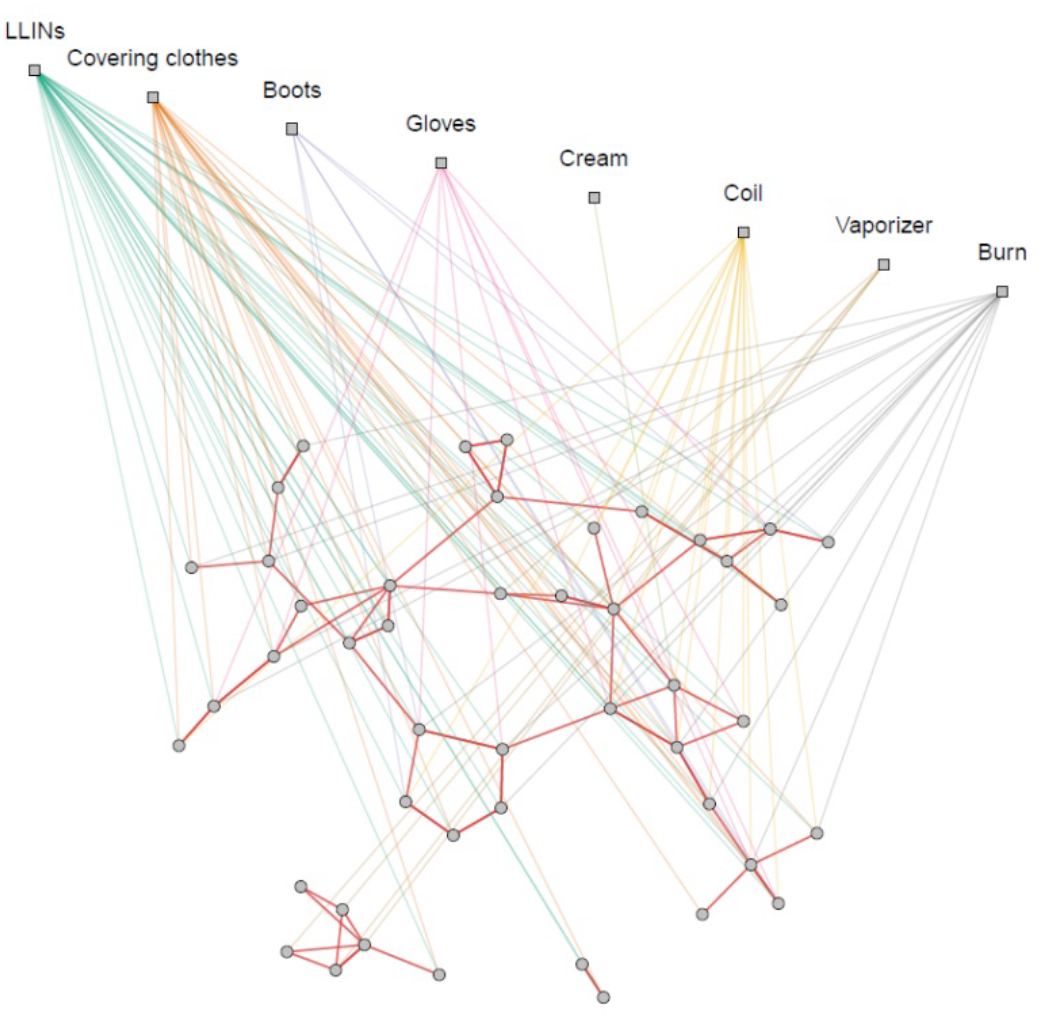
Multilevel network of people by people talking about health-related matters and using preventive measures in WK2. Appendix C in the Supplementary information (Figures C1-C10) reproduces multilevel network visualizations for each village.

## RESULTS

### Factors associated with the use of different preventive measures

Results of meta-analyses of village-level logistic regressions explaining the use of each preventive measure are reported in Table 1. The table presents estimates of population parameters (μ) and standard errors (SE) of variables that were retained through backward model selection performed separately for each preventive measure. See the Methods section for details about variable selection and meta-analysis models. Heterogeneity statistics for estimates are reported in the Supplementary Information (Appendix D, Tables D19-D26).

**Table 1:**
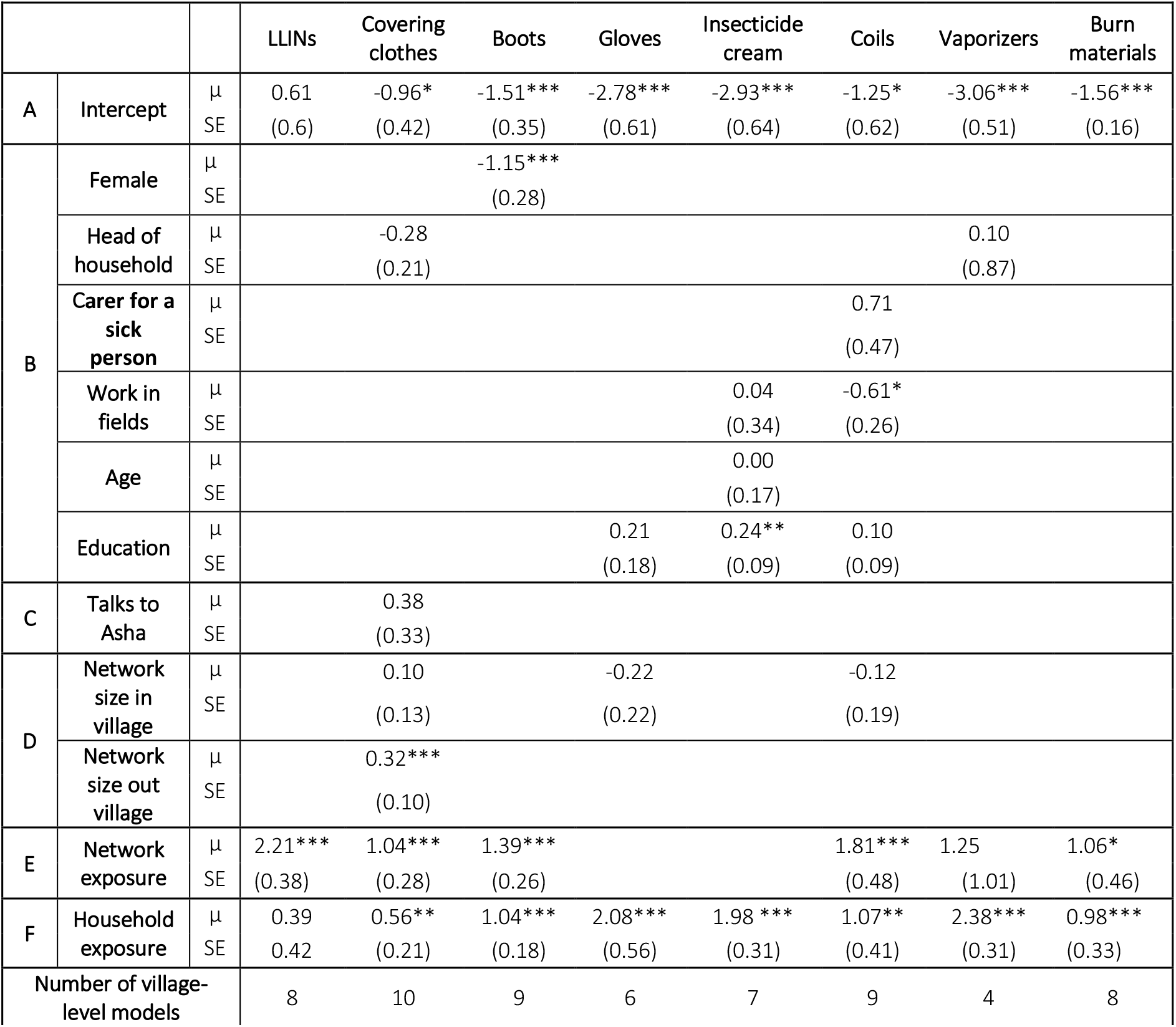
Results of meta-analyses of parameters from village-level logistic regression models explaining the use of each preventive measure. Variables were chosen for each measure by backward model selection as described in the Methods section; variables are grouped as A: intercept; B: Individual characteristics; C: Opinion leaders; D: Network size; E: Network exposure; F: Household exposure; μ – estimated population parameters (log odds ratios); SE – standard errors of estimates; p values: * <0.05, ** <0.01, *** <0.001.

**Table 1** shows that at least two explanatory factors were retained in the model selection process for every preventive measure. The meta-analysis of parameters highlights that there were two of these that were consistently associated with measure use across the villages.

First, network exposure significantly increased the probability of using five of the eight measures, except for gloves, insecticide cream, and vaporizers (odds ratios ranging from 2.8 to 9.1). Household exposure was significantly positively associated with the use of measures in seven cases, with the only exception LLINs (odds ratios for significant estimates between 2.7 and 10.8). These effect sizes are quite substantial, considering that the average number of discussion partners and the average household size are both 2 in the entire dataset. For example, our models predict that one additional discussion partner using boots would double the odds of a villager with an average-sized network using boots (network exposure log odds ratio 1.39).

Other explanatory variables did not have a consistent impact across preventive measures. Of the considered individual characteristics, only gender (boots), working in fields (coils), and education (insecticide cream) had a significant effect on measure use and only for one measure. The number of reported discussion partners outside of one’s village only had a significant positive effect on the use of covering clothes. Talking to village opinion leaders, ASHAs and Traditional Healers, did not have a significant impact on the use of any of the measures. These results do not indicate that individual variables, opinion leaders, and network size are not associated with the use of measure at all. However, the effects of these factors vary from village to village (see the Supplementary Information, Appendix D, Tables D10-D17 for village-level model results).

### Relevance of network and household exposure for explaining the use of measures

Next, we examine the accuracy of village-level models to assess the importance of different factors in explaining the use of preventive measures. To do so, we fit seven nested models for each preventive measure in each village, which include different subsets of the explanatory variables, as marked by the letters A-F in Table 1. The Empty Model contains an intercept term only (subset A) and serves as a baseline for accuracy per measure and village. The Individual Model includes the individual variables that were used for the given measure as well (A+B). The Opinion Leader Model further includes any effects of talking to the ASHA or the Traditional Healer (A+B+C). The Network Size Model considers any included variables related to the number of discussion ties (A+B+C+D). The Network Exposure Model (A+B+C+D+E) and the Household Exposure Model (A+B+C+D+F) add the relevant exposure term to the Network Size Model. The Full Model for each preventive measure is the same as the one reported in **Table 1**.

We compare the accuracy (correct classification rate) of the above models. The results are presented in **Figure 4**. The average accuracy of the Empty Model is 77%. Adding individual (77%), opinion leader (78%), and network size variables (79%) makes only a small difference. In contrast, including either network exposure or household exposure improves average accuracy by 2% and 3% compared to the Network Size Model. The Full Model, which includes both exposure terms beyond the Network Size Model, has an average accuracy of 83%. (For village- and measure-level classification rates, see the Supplementary Information, Appendix D, Tables D29-38).

**Figure 4.**
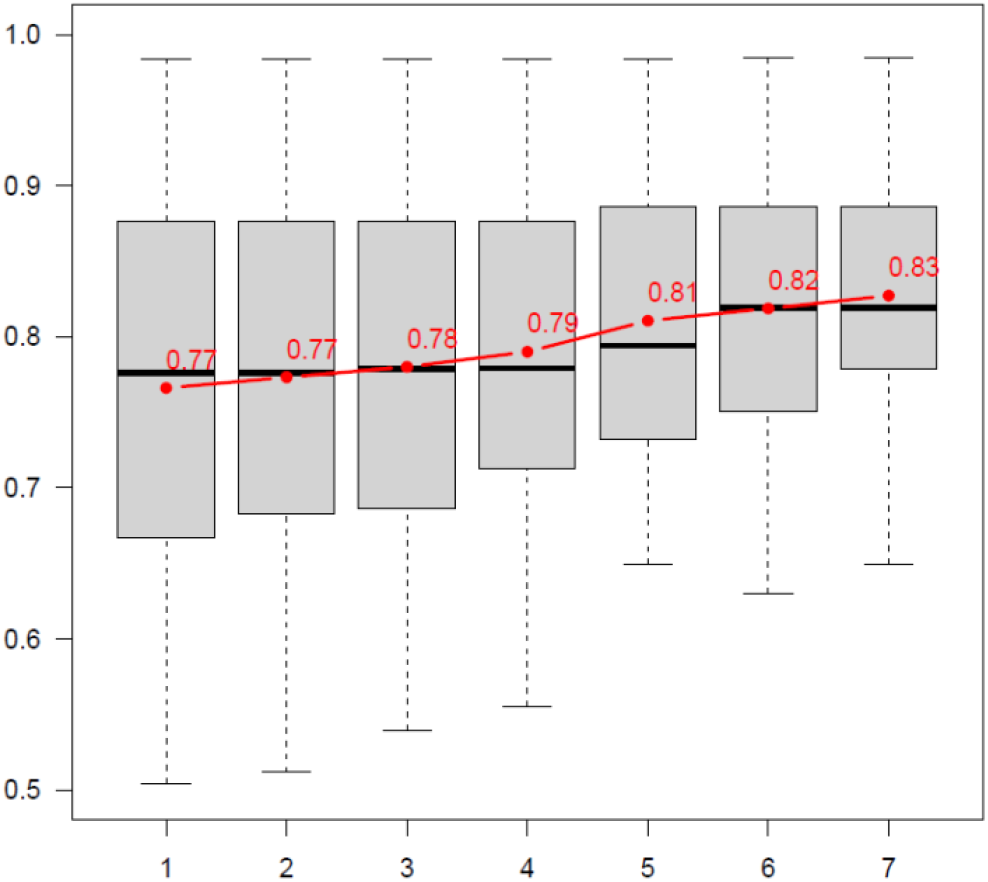
Distribution of the accuracy of different village-level logistic model specifications. Each bar represents models using different variable subsets of the models in Table 1, denoted by A-F; accordingly, each bar represents the accuracy of 61 models; the connected red points and values show the average accuracy of models; stars represent p-values for two-sided pairwise sample t-tests of the difference in the average accuracy of models adjacent in the figure: *p<0.05, **p<0.01, ***p<0.001 (for full t-test statistics, see the Supplementary Information, Tables D27-D20).

Overall, the 6 percentage point improvement in accuracy between the Empty Model and the Full Model, and the 4 percentage point increase from including network and household exposure alone, may not appear high. However, these numbers should be interpreted in the context of a high baseline accuracy due to the skewness of measure use in many cases (e.g., LLINS are adopted by nearly everyone). Considering this, it is noteworthy that network and household exposure reduce the classification error of the Network Size Model by 20%, from 0.21 to 0.17. These results suggest that the two exposure factors substantially contribute to explaining the use of preventive measures.

### Social mechanisms associated with preventive measure use

The results presented so far indicate that network and household exposure are key factors explaining the use of preventive measures in our villages. To explore the specific social mechanisms which may explain measure use, we fitted stationary SAOMs to the multilevel network of use and discussion ties, as defined earlier, in each village. We summarise the results of ten similarly specified SAOMs (one per village) by a meta-analysis of parameters in Table 2. As we are interested in the use of preventive measures, here we only discuss model effects that explain which measures are used by individuals. The models also simultaneously treat the village discussion network as an outcome that co-evolves with measure use (see the Methods section for further details on the model). This allows us to disentangle effect directions between discussion and use, and thus take a step closer to identifying likely causal impacts of social networks on the use of measures. The full results of the SAOM meta-analysis, all village-level model estimates, convergence statistics, and goodness-of-fit statistics are reported in the Supplementary Information, Appendix E, Tables E1-E6.

**Table 2:**
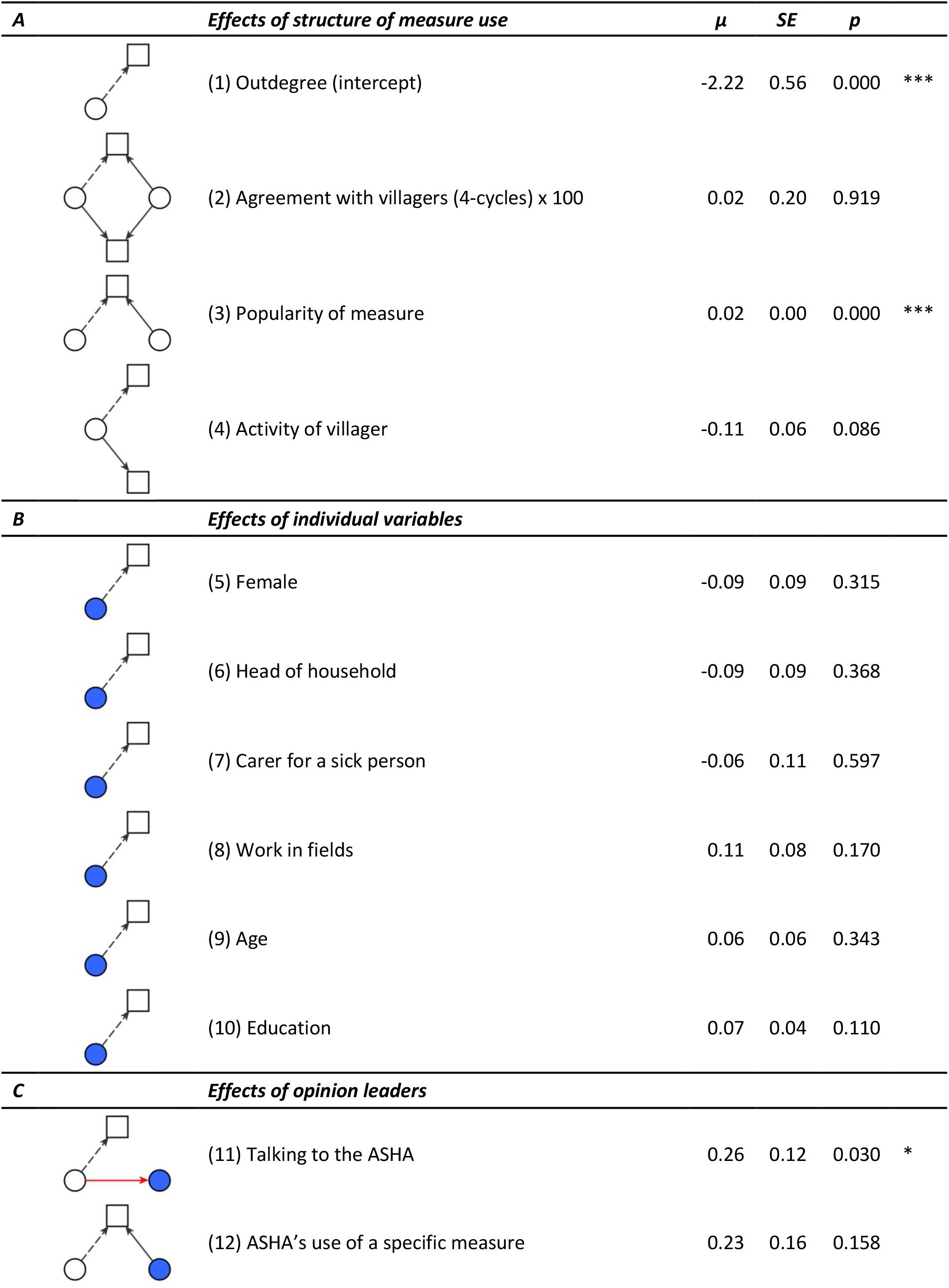
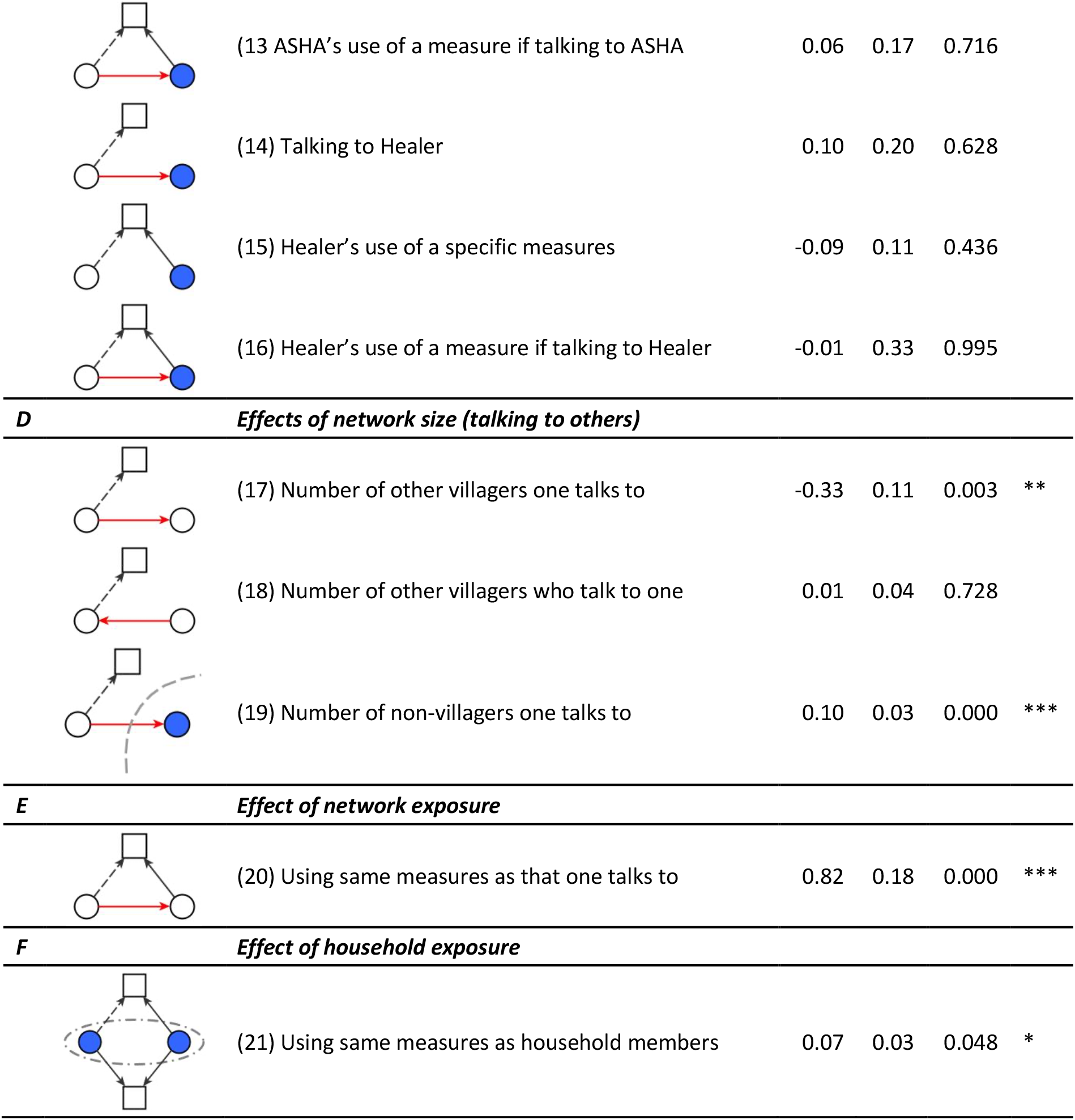
Meta-analysis of estimates from multilevel stationary SAOMs in each village. Circles represent villagers; in each effect (explanatory variable), blue circles indicate villagers with specific characteristics (e.g., gender), squares represent preventive measures, red arrows represent health-related discussion ties, black solid arrows represent preventive measure use, and black dashed arrows represent the dependent tie, the probability of which is assessed by the given effect; rate parameters were fixed at 2.5 for both networks in all models; all models converged according to the criteria set out in Ripley et al. (2022) – see the Supplementary Information, Appendix E, Tables E5-E6 for further details; μ – estimated population parameters (log odds ratios); SE – standard errors of estimates; p values: * <0.05, ** <0.01, *** <0.001.

| **A** | ***Effects of structure of measure use*** | **$\mu$** | ***SE*** | ***p*** |  |
| --- | --- | --- | --- | --- | --- |
|  | (1) Outdegree (intercept) | -2.22 | 0.56 | 0.000 | *** |
|  | (2) Agreement with villagers (4-cycles) x 100 | 0.02 | 0.20 | 0.919 |  |
|  | (3) Popularity of measure | 0.02 | 0.00 | 0.000 | *** |
|  | (4) Activity of villager | -0.11 | 0.06 | 0.086 |  |
| **B** | ***Effects of individual variables*** |  |  |  |  |
|  | (5) Female | -0.09 | 0.09 | 0.315 |  |
|  | (6) Head of household | -0.09 | 0.09 | 0.368 |  |
|  | (7) Carer for a sick person | -0.06 | 0.11 | 0.597 |  |
|  | (8) Work in fields | 0.11 | 0.08 | 0.170 |  |
|  | (9) Age | 0.06 | 0.06 | 0.343 |  |
|  | (10) Education | 0.07 | 0.04 | 0.110 |  |
| **C** | ***Effects of opinion leaders*** |  |  |  |  |
|  | (11) Talking to the ASHA | 0.26 | 0.12 | 0.030 | * |
|  | (12) ASHA's use of a specific measure | 0.23 | 0.16 | 0.158 |  |
|  | (13) ASHA's use of a measure if talking to ASHA | 0.06 | 0.17 | 0.716 |  |
|  | (14) Talking to Healer | 0.10 | 0.20 | 0.628 |  |
|  | (15) Healer's use of a specific measures | -0.09 | 0.11 | 0.436 |  |
|  | (16) Healer's use of a measure if talking to Healer | -0.01 | 0.33 | 0.995 |  |
| **D** *Effects of network size (talking to others)* |  |  |  |  |  |
|  | (17) Number of other villagers one talks to | -0.33 | 0.11 | 0.003 | ** |
|  | (18) Number of other villagers who talk to one | 0.01 | 0.04 | 0.728 |  |
|  | (19) Number of non-villagers one talks to | 0.10 | 0.03 | 0.000 | *** |
| **E** *Effect of network exposure* |  |  |  |  |  |
|  | (20) Using same measures as that one talks to | 0.82 | 0.18 | 0.000 | *** |
| **F** *Effect of household exposure* |  |  |  |  |  |
|  | (21) Using same measures as household members | 0.07 | 0.03 | 0.048 | * |

Like in any SAOM, the first set of parameters (A) control for the overall structure of the use network. The outdegree (density) effect (effect (1) in Table 2), which is significantly negative, is the intercept and highlights that using a measure is not very common on average: the estimated baseline probability is 11%. The 4-cycle effect (2) expresses tendencies for similarity of use between individuals, which is not significant. The positive popularity of measures effect (3) reflects that villagers are significantly more likely to use measures which are already used by several others. The non-significant villager activity effect (4) suggests that there are no differences in villagers’ tendency to use a measure based on how many other measures they are currently using.

The second set of parameters (B) models individual characteristics (5-10). None of these variables explain the use of preventive measures significantly, in line with the results of the logistic models. The third set of parameters (C) looks at the role of opinion leaders (11-16). Here, contrary to what we found for individual measures, talking to the ASHA significantly increases the odds of using any preventive measure, by 30% on average (11). However, talking to the Traditional Healer and considering the specific measures that the ASHA and the Traditional Healer uses do not seem to matter consistently across the villages. The fourth set of parameters (D) looks at effects of discussion network size (17-19). We find that talking to more people in one’s own village significantly reduces (17), while talking to more people outside of the village increases the odds of using any preventive measure (19) (odds ratios 0.72 and 1.10). Finally, being talked to by many within one’s village does not significantly influence the likelihood of using measures (18).

The last two sets of parameters (E-F) model network and household exposure (20-21). Both parameters are significant and positive. Network exposure (20) more than doubles the odds of use of a measure for each additional discussion partner who reports using the same measure (odds ratio 2.27). Further, if two members of a household use the same measure (21), it is more likely on average that they both use other measures as well (odds ratio 1.07). While both effects appear substantial in size, their relative importance may vary by individual, depending on the size of their discussion networks and households.

### Network and household exposure explain patterns of preventive measure use and health-related discussions

The above results highlight the importance of considering villagers’ discussion networks when we explain the use of preventive measures. To assess how well our village-level SAOMs capture the connections between the discussion and use networks, we compare the goodness of fit of different model specifications that include subsets of the effects in the full model in Table 2. **Figure 5** shows the distribution of SAOM goodness of fit p-values with regard to the mixed triad census of the two modelled networks (Hollway et al., 2017). We find that fit of a Baseline Model considering only endogenous structural effects in the discussion and use networks (effect group A) generally achieves a poor fit on these statistics. Sequentially adding effects of individual characteristics (A+B), opinion leaders (A+B+C), and network size (A+B+C+D) does not improve the fit. Adding network exposure (A+B+C+D+E) or household exposure (A+B+C+D+F) leads to a large improvement in model fit, while there are further gains from including both types of exposure effects (Full Model). We note that excluding network exposure from the Full Model has a statistically significant negative impact on fit (paired sample one-sided t-test: difference=-0.12, t=2.22, p=0.03, df=9), while excluding household exposure does not (difference=-0.08, t=1.63, p=0.07, df=9). These results provide a clear indication that effects from network exposure and household exposure are crucial in explaining the links between the use preventive measures and discussion networks. Village-level goodness of fit statistics are reported in the Supplementary Information, Appendix E, Tables E5-E6.

**Figure 5.**
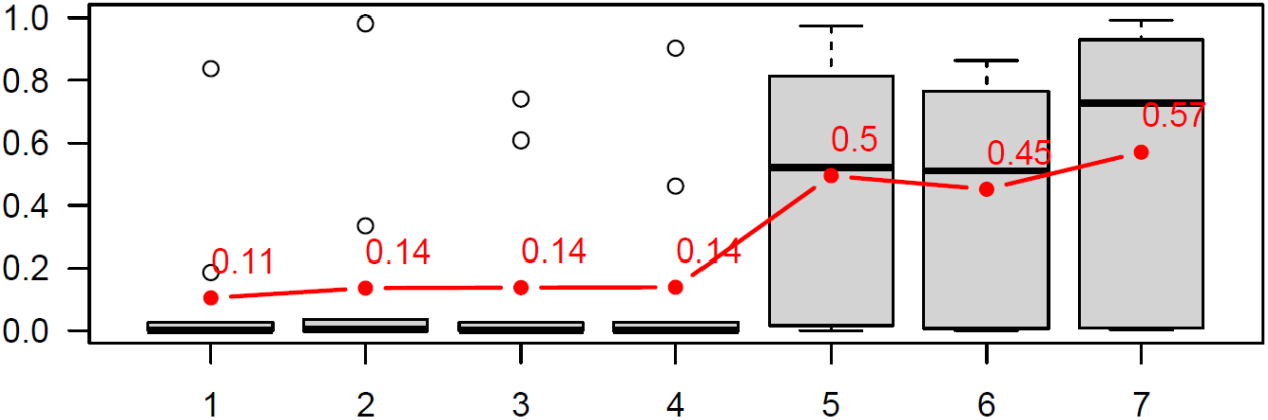
Comparison of village-level SAOM specifications by goodness fit (GoF) p-values regarding the combination of talk network and measure use ties. Each bar represents models using different variable subsets from Table 2, denoted by A-F; accordingly, each bar represents GoF results of 10 models; the considered GoF statistic is the mixed triad census of Hollway et al. (2017); the connected red points and values show the average GoF p-values of the models; stars represent p-values for one-sided pairwise sample t-tests of the difference in the average GoF p-value of models adjacent in the figure: *p<0.05, **p<0.01, ***p<0.001.

## DISCUSSION

Our results demonstrate that considering social networks contributes to the explanation of health-related behaviours and practices. We examined a variety of factors that may influence the uptake of eight malaria-preventive measures in ten hard-to-reach tribal communities in northeast India, using standard statistical methods and social network models. We found that exposure to the use of preventive measure in one’s social network was consistently associated with own use. In both standard and network models explaining measure use, we showed that network exposure significantly improves the explanatory power of the models. Exposure in households had a comparable role in our analyses, while factors typically considered in studies of health behaviour, such as individual characteristics [Nlinwe et al., 2021; Matin et al., 2020; Van Eijk et al., 2016; Shahandeh et al, 2012] and opinion leaders [Stoebenau and Valente 2003; Valente and Pumpang 2007], had little to no effect.

The findings about social network effects are theoretically relevant as they provide empirical support for core ideas of network exposure models [Coleman et al., 1957; Valente, 1996; Valente 2010; Centola, 2018]. According to these, information and attitudes regarding both effective and ineffective preventive behaviours should be less dependent on who people are and more on whom they talk to. This suggests that research on health behaviours and social influence could benefit from further theoretical work that explores specific social mechanisms of network exposure. The effect of household members’ behaviour is also important, especially because exposure in households does not necessarily have to rely on discussions. People may use preventive measures simply because they are available in the house where they live, where exposure can be tacit and reinforced by observing use rather than talking about it. Collecting data on social networks and household composition, information on which are both not immediately available in contexts where census data are lacking, requires considerable amount of time and resources, but our analyses clearly show that these can provide novel theoretical and empirical insight into social processes explaining health behaviours.

We found a positive effect of discussion ties extending outside villages on the likelihood of using malaria preventive measures. We know from our data that these ties are mainly constituted by family and in-law ties (Of the 841 ties to people from other villages, 74% are to family members, 11% to relatives, and 15% to non-blood contacts). In Meghalaya, a matrilinear society, upon marriage sons move out of their maternal homes to stay at theie spouse’ home, while daughters remain close to their families of origin. The move of sons across villages creates networks of partially weak ties [Granovetter 1978]: these relationships are weak, according to Granovetter’s definition [1978] in the sense that they are geographically distant and in infrequent contact. They are also potential sources of new information and attitudes about preventive techniques as they reach outside one’s own, closely-knit village community. At the same time, family and relatives in different villages are emotionally close enough – a characteristic normally associated with strong ties [Granovetter 1978] to rely on each other for health-related discussions, which increases the chance that they actually influence villagers’ health-related attitudes and behaviours. Therefore, intervillage ties complement the effects of network and household exposure on the use of preventive measures. The relatively small effect and explanatory importance of these ties may be due to the fact that they are informationally redundant: family and relatives likely all talk to each other, so they may end up sharing similar information or opinions. These findings point to the relevance of studying the extended family as a key social unit in shaping health-related behaviours and practices.

Interestingly, our analysis shows almost no effects from opinion leaders on using preventive measures. The literature suggests that they are rarely early adopters, but they could be instrumental in facilitating or hampering the spread of innovations [Valente and Pumpang 2007]. Our data does not allow to observe when exactly the ASHA and the Traditional Healer started using specific preventive measures, but results clearly indicate that their behaviour does not influence other villagers regardless if they talk to them or not. Talking to the ASHA does have an effect on using more measures, but this is small and it does not substantially contribute to model fit. This result may inform the planning of future interventions, as it indicates that relying solely on central actors like the ASHA may not be effective in improving disease prevention.

Similarly, we find little evidence that any of the individual characteristics we considered would consistently matter for use across villages and preventive measures. We did see indications in the logistic models that gender, type of work, and education are associated with measure use in case of a single measure. Other characteristics also appear to be relevant, but only in some villages and in case of a few preventive measures (see the Supplementary Information, Appendix D, Tables D10-D17). Overall, our analyses do not suggest that individual factors do not matter at all, only that their effects on the use of measures may differ from community to community. By exploring these differences in a localised geographic context, large-scale public health interventions can be effectively tailored to the attitudes and practices of relevant social groups.

Our study has a few key limitations that may inform the design of future research into health-related behaviours. First, we collected self-reported data on behaviours and social ties. Self-reports are known to be sensitive to recall bias [Bernard and Killworth 1977; Small 2017] as well as socially desirable responses [Edwards 1953]. We aimed to attenuate this issue by keeping questions about prevention practices and discussion ties general. We only asked participants to report on whether they use different preventive measures or not (we queried frequency of use regarding covering clothes and insecticide cream but disregarded this detail in the analyses for comparability; see the Supplementary Information, Appendix B1 for details). We measured social networks by asking people whom they talk to about health-related matters without reference to details of these exchanges. While these questions could reduce recall problems, they did not inform us about the frequency and contexts of measure use and the content of discussions. Employing field observations [Bernard and Killworth 1977; Curtis et al., 1995] and smart sensor technologies [Cattuto et al., 2010; Elmer et al., 2019; Voros et al. 2021] may help to gather richer data. However, trade-offs between data quality and resolutions should be carefully considered, as these methods are more intrusive and may lead to lower participation rates [Voros et al. 2021] and alter behaviour [Landsberger, 1958]. Our interview-based approach helped us to gain the trust of our participants and elicit meaningful responses about general patterns of measure use and discussion ties.

Second, the validity of our findings should be assessed in the context of coverage and sample size. Our overall response rate is 68%, which is considered sufficiently high for statistical network analyses [Kossinets 2006]. However, coverage was not equal across the three regions and was only just over 60% in West Jaintia Hills. This partially compensated with the higher coverage of households (80%). Overall, our data collection efforts have resulted in reasonably high response rates that indicate the adequate internal validity of our results. In turn, our findings should not be generalised beyond the studied contexts. We only had the resources to observe a small proportion of communities in just three districts of Meghalaya state. Nonetheless, we found some common patterns in the studied communities which suggest that processes from network and household exposure may help to explain the use of preventive measures. We need larger-scale, regionally representative datasets to confirm these findings and gain a more complete picture of the social mechanisms driving health-related behaviours. Our results further suggest that future work should focus on collecting detailed data on weak ties and inter-villages relationships. Combined with a large-scale approach, this could improve our understanding of how effectively preventive measure spread across malaria endemic regions.

Third, we interpret our key results in terms of social processes, while we collected and analysed cross-sectional data. Clearly, analysing one snapshot of networks and behaviours comes with limitations. We do not know how the use of measures changed over time. It is also possible that some measures were becoming popular, while others unpopular at the time of the data collection. We do not have information about the stability of health-related discussion ties either, although the network literature suggests that such relations should be stable in the short-term [Hogan et al., 2022]. In line with these concerns, our logistic regression results only highlight associations, without considering the effects of measure use on social networks. The Stochastic Actor-oriented Model is better suited to identify the social processes at play. Even when applied to cross-sectional data, the SAOM is dynamic: it assumes that villagers may still change their behaviour and network ties, but these changes keep the overall structure of the multilevel network in a stable state (in a “short-term dynamic equilibrium”; Snijders and Steglich 2015). This means that the model considers both the effects of network ties on measure use and vice versa (see full results the Supplementary Information, Appendix E, Tables E1-E2). By concurrently modelling the dependencies between network ties and behaviours, we may infer to the social mechanisms that are likely to maintain the network system [Lusher et al. 2012], such as network or household exposure. Any causal claims that are made on the basis of “multi-mechanistic” network models [Stadtfeld and Amati 2021], such as the SAOM, are stronger than those based on results from non-network models. At the same time, collecting longitudinal data and studying the actual changes of networks and behaviour will be required to gain a clear understanding of the dynamics of measure use. Our approach can be easily adapted to serve as a basis of such a longitudinal design.

For the first time since 2015, when malaria elimination efforts were initiated in India [PIB 2020], local elimination may be within reach. However, current control efforts rely solely on mass interventions. These include the large-scale distribution of LLINS that was interrupted in India in recent years [WHO 2022], and IRS which is met with high levels of refusal in Meghalaya [Sarkar et al., 2021; Passah et al., 2022]. Our study suggests that these interventions could be complemented with targeted efforts that leverage social influence in interpersonal relations and households to increase uptake. This could help to achieve downstream elimination goals. However, further research is needed to map the patterns of measure use across different areas and social groups and to understand the social mechanisms that may facilitate or hamper the adoption of various health behaviours. This line of work may contribute to increasing the acceptance of a malaria vaccine in the future.

Beyond the context of malaria, exploring the role of social networks in health behaviour could benefit the prevention of other serious mosquito-borne diseases, such as Dengue, Chikungunya and the newly emerging Zika virus [Gulland 2016], which are on the rise both globally and in India [Shepard et al. 2014].

## METHODS

### Data collection

From January 2020 through August 2021, we collected data from three villages in West Khasi Hills (WK1, WK2, WK3), three in West Jaintia Hills (WJ1, WJ2, WJ3) and four in South Garo Hills (SG1, SG2, SG3, SG4) in Meghalaya, India. Villages were selected based on their manageable size (<500 eligible adults), their known willingness to participate in similar studies (we avoided villages where previous epidemiological studies encountered high rates of refusal, see Kessler et al., 2018), and their accessibility by either car or foot. Village-level coverage ranged from 53% of residents in WJ3 (80% of households) to 88% in WK3 (97% of households), with an average of 68% of individuals (n=1,530) and 80% households (a total of 766) interviewed. The topic of coverage is addressed in the Discussion section. Characteristics of non-respondents are reported in the Supplementary Information, Appendix A, Table A2.

Structured interviews were conducted in the appropriate local language (Khasi, Pnar or Garo) and translated into English by the interviewing team. Respondents were asked a series of questions regarding their individual background, household and village roles, their use of malaria preventive measures, and the people in and outside of their village they talk to about health-related issues. In total, each interview consisted of 26 questions and lasted about 30 minutes. The full list of interview questions, details about data coding and processing, and descriptive statistics for variables are reported in the Supplementary Information, Appendix B, Tables B1-B15.

The ASHA was interviewed in all ten villages, and in the six villages where present, the Traditional Healer was also interviewed. As we consider the ASHA and the Traditional Healers as opinion leaders, whose behaviour and influence may be substantially different from those of other villagers, we removed these two actors as respondents from our analyses. We still utilize information about their use of measures and about who talks to them to explain the behaviour and social networks of other villagers.

Permission to conduct the study was granted by the Headman of each village, and all respondents also signed an individual informed consent form. Identifiers of the villages and individual participants were removed from the dataset and this article to protect anonymity. Ethical approval for the study was obtained from the Institutional Review Boards (IRBs) of Martin Luther Christian University, Shillong, Meghalaya, India and New York University, New York, NY, USA.

### Variables

The dependent variables in our analyses represent the use of eight malaria prevention measures by participants: Long-Lasting Insecticidal Nets (LLINs), covering clothes, boots, gloves, insecticide cream, coils, vaporisers, and burning materials. We considered how a variate of explanatory variables were associated with these outcomes in different multivariate statistical models. In total, 14 relevant explanatory variables were available in the dataset and were considered in our analyses. We reduced this set for the meta-analysis of logistic regressions by a model selection procedure described below. The fitted social network models estimated a larger number of parameters, because each variable can affect the outcome in multiple ways in these models.

The starting set of explanatory variables was the following:

- **ndividual characteristics and roles in the household:** (1) gender, (2) being the head of the household, (3) looking after family members when they are sick (4) working in
- fields, (5) age, and (6) educational background.
- **Contact with opinion leaders:** the participant or someone from their household talks to the Asha (7-8) or the Traditional Healer (9-10) of the village about health-related issues.
- **Social network size:** the number of people in and outside the village the participant talks to about health-related issues (11-12).
- **Social network exposure:** the amount of people the participant talks to who use a given preventive measure (13)
- **Household exposure:** the amount of people in the participant’s household who use a given preventive measure (14)

### Logistic regression model selection procedure

We considered 14 explanatory variables for each logistic model explaining the use of a given preventive measure in a specific village (see the Supplementary Information, Appendix B, for the complete list of variables). Given the size of our villages and the often skewed distribution of measure use, we found that in most cases models including all 14 explanatory factors could not be estimated. Therefore, we employed a two-stage model selection procedure aimed at reducing the set of explanatory variables while providing a single specification per measure that can be estimated in every village. First, we carried out backward model selection based on the p-values of parameter estimates for each of the 8 (measures) x 10 (villages) = 80 models. This resulted in different optimal model specifications across measures and villages. Second, considering each measure separately, we added variables back that had a significant estimate in at least two of the villages. This resulted in 8 different model specifications, one for each preventive measure, that were identical across the villages. The exact steps of our procedure are detailed in Procedure 1. Estimating the reduced models was still difficult in case of some villages and preventive measures. We omitted models from the meta-analyses reported in the text that had very large estimated parameters and/or standard errors for at least one explanatory variable. The presented results are based on 61 models. See the Supplementary Information, Appendix D, Table D18 for the excluded models.

**Procedure 1. Two-stage model selection approach**

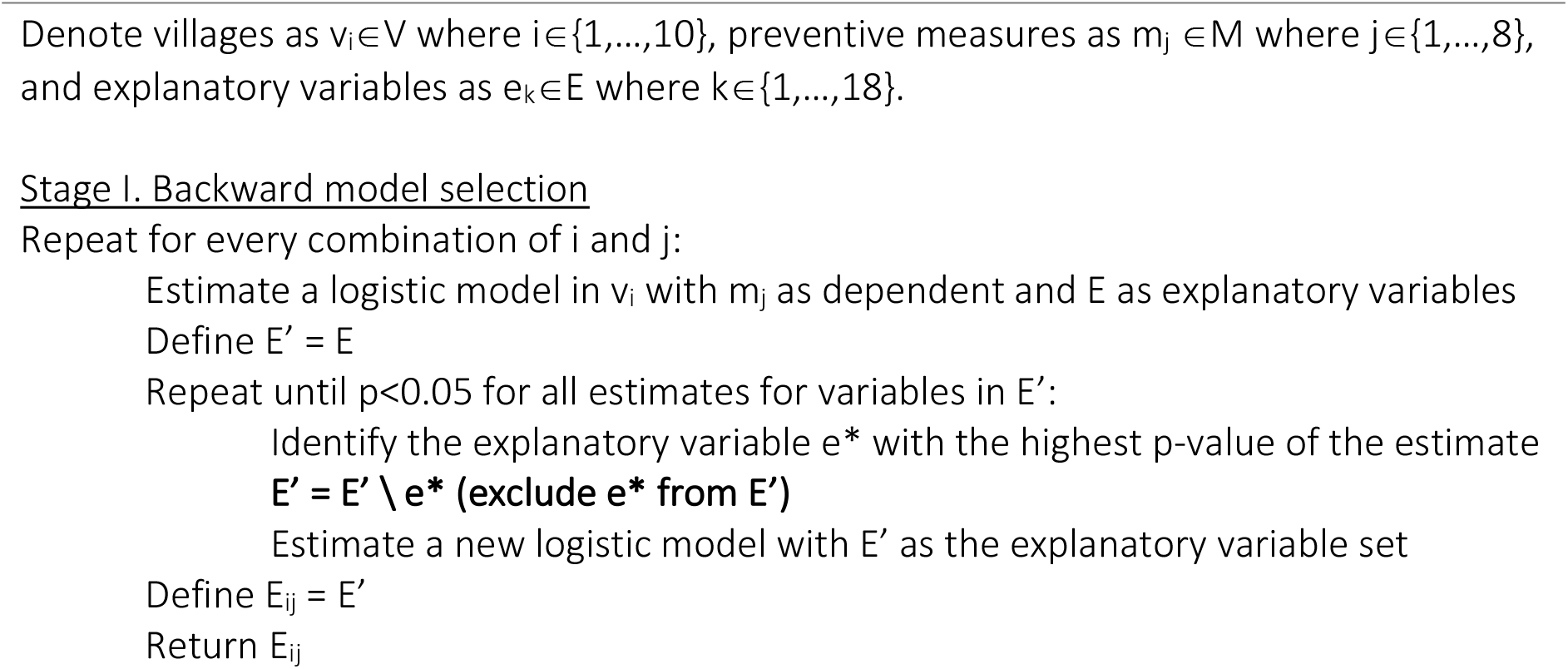

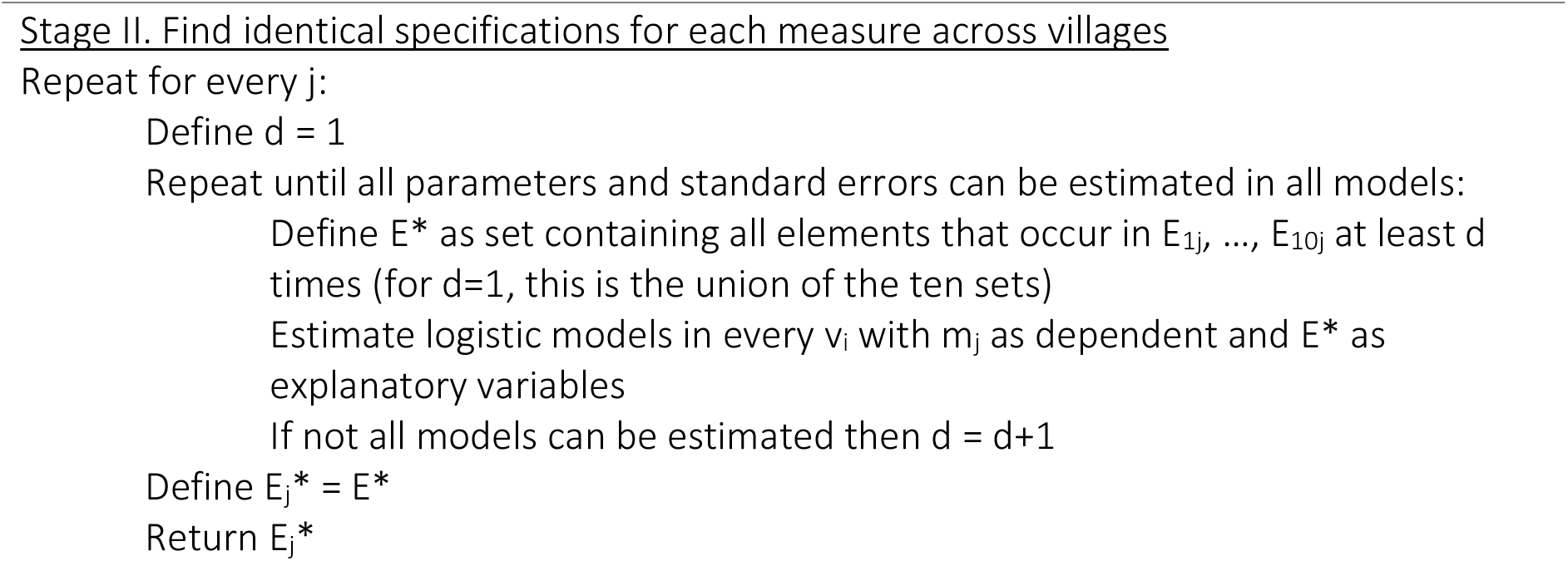

### Meta-analyses of village-level logistic regressions

To explore recurrent patterns of association between the use of different measures and our explanatory variables across villages, we meta-analysed the model results obtained from the model selection procedure described above. We perform parameter-wise meta-analyses with random-effects models estimated by restricted maximum likelihood using the metafor package (v3.8-1) in R (Viechtbauer 2010). Estimates provide information about the mean and standard error of parameters. Heterogeneity statistics are reported in the Supplementary Information, Appendix D, Tables D19-D26. The meta-analysis results are presented in Table 1.

### Comparison of the explanatory power of nested logistic regression models

We compared the explanatory power of the village-level logistic models described above and of nested models that contain different subsets of the explanatory variables. We calculated the accuracy for each model as the correct classification rate: the percentage of cases for which the observed value of the dependent variable (whether a villager uses a given measure or not) equals the estimated value obtained from the model. We used the standard 0.5 cut-off for transforming probabilities into estimated binary outcomes. The results of this analysis are presented in Figure 4, while the accuracy of the village-level models for each village are reported in Supplementary Information, Appendix D, Tables D29-D38.

### Meta-analysis of village-level stationary Stochastic Actor-oriented Models

For a more detailed analysis of the social mechanisms affecting the use of preventive measures, we employed stationary Stochastic Actor-oriented Models (SAOMs). Stationary SAOMs are an extension of the original model [Snijders et al. 2010; Snijders 1996] for the analysis of cross-sectional network data. In this case, it is assumed that the networks studied are in a short-term dynamic equilibrium: their structure (but not single network ties) is at least temporarily in a stable state [Snijders and Steglich 2015]. SAOMs are empirically calibrated simulation models that aim at identifying the relative strength of a set of social mechanisms, operating on network ties, individual and pairwise (dyadic) covariates, that over time could have generated an observed network. In a standard longitudinal SAOM, the model is conditional on the first observation of the network [Snijders et al. 2010]. In a stationary SAOM, the initial and final states of the network are identical, and the modelled social

### processes will, at least stochastically, maintain the existing structure of the network (hence the ‘stationary’ label) [Snijders and Steglich 2015]

To apply stationary SAOMs, we defined a multilevel network that consists of two interlinked networks in each village: 1) the network of health-related discussions, which is a one-mode network where villagers A and B are connected by a binary tie if A reports talking to B; and 2) the network of preventive measure use, which is a two-mode network where villager A is connected to measure M if A reports using M. An example of this network for one of the villages is presented in Figure 3. Similar visualisations for all villages can be found in the Supplementary Information, Appendix C.

We fit stationary SAOMs using the RSiena (v1.3.0) package in R (Ripley et al. 2022). We set the rate parameter to 2.5 for both networks in all villages, as this value enabled model convergence and led to good model fit in every case. The key results are interpreted in the main text. The full list of estimates, goodness of fit tests and robustness checks for rate parameters are presented in the Supplementary Information, Appendix E. Similar to the case of logistic regressions above, we perform parameter-wise meta-analyses of village-level SAOMs with random-effects models estimated by restricted maximum likelihood using the metafor package (v3.8-1) in R (Viechtbauer 2010). Estimates provide information about the mean and standard error of parameters. Heterogeneity statistics are reported in the Supplementary Information, Appendix E, Table E4. The meta-analysis results are presented in Table 2.

### Comparison of the explanatory power of nested SAOMs

We compared the explanatory power of the village-level SAOMs described above and of nested models that contain different subsets of the model effects. We calculated p-values of Mahalanobis distances for a mixed triad census fit test for different SAOM specifications in the results section. These reflect how well the distribution of simulated networks based on the fitted models represent the mixed triad census [Hollway et al., 2017] of the observed networks: a p-value close to 1 signifies very good fit and one close or equal to 0 marks poor fit [Ripley et al., 2022]. Calculations in each model are based on 5000 simulations and follow the steps described in Lospinoso and Snijders [2019]. The goodness of fit tests were carried out using the RSiena R package [Ripley et al., 2022]. The results are presented in Figure 5.

Further details of the goodness of fit analyses can be found in the Supplementary Information, Appendix E, Table E5.

## Data Availability

All data produced in the present work are contained in the manuscript

## ACKNOWLEDGMENTS

We thank the study participants for their time, our field teams for their data collection and transcribing efforts, and the Meghalaya Malaria State Control Programme for their support and ongoing collaboration. We also thank Drs. Mark Wilson and Steven Sullivan for manuscript editing and input. Research reported in this publication was supported by the National Institute of Allergy and Infectious Diseases of the National Institutes of Health under Award Number U19AI089676. The content is solely the responsibility of the authors and does not necessarily represent the official views of the National Institutes of Health.

## AUTHOR CONTRIBUTIONS

E.B., A.K., A.v.E., J.C., and S.A. designed the study and analyses. M.P. C.B.N, Q.D.N and C.K. conducted the questionnaires and collected the data, under the supervision of R.S. and S.A.

E.B. and A.V. analysed the data and wrote the first draft of the paper. All authors finalized the manuscript. J.C. obtained the funding.

## ADDITIONAL INFORMATION

## Supplementary information

**Correspondence and requests for materials** should be addressed to Elisa Bellotti and Sandra Albert.

## Data availability

TBA

## Code availability

TBA

