## Supplementary Information for "Social network and household exposure explain the use of malaria prevention measures in rural communities of Meghalaya, India"

##### Summary

This supplement provides additional information about the data collection and the analytical strategy of the study we conducted in ten indigenous hard to reach villages in three regions of Meghalaya state in India. We first provide an overview of the sampling strategy and details of the data collection (Appendix A). This is followed by an overview of the questionnaire and variables used in the main article (Appendix B). We then present visualisations the multilevel social networks of villagers discussion ties and use of malaria preventive measures for each village (Appendix C). We discuss details of the logistic regression analyses presented in the main article (Appendix D). Lastly, provide background and details for the Stochastic Actor Oriented Models presented in the main article (Appendix E).

**Appendix D. Results of the analyses using logistic regression models** ....... Error! Bookmark not defined.

D.1 Model selection I: Backward variable selection in each village .........**Error! Bookmark not defined.**

D.2 Model selection II: Identical model specifications across villages.....**Error! Bookmark not defined.**

D.3 Meta-analysis of village-level logistic regression models ... **Error! Bookmark not defined.**

D.4 Assessment of the goodness of fit of village-level logistic regression models..........**Error! Bookmark not defined.**

**Appendix E. Results of the analyses using Stochastic Actor-oriented Models (SAOMs)** .... Error! Bookmark not defined.

E.1 Overview of modelling approach ....................................... **Error! Bookmark not defined.**

E.2 Model specification and definition of effects ...................... **Error! Bookmark not defined.**

E.3 Village-level SAOM results.................................................. **Error! Bookmark not defined.**

E.4 Full SAOM meta-analysis results ........................................ **Error! Bookmark not defined.**

E.5 Assessment of the goodness of fit of village-level SAOMs.. **Error! Bookmark not defined.**

E.6 Robustness checks with different SAOM rate parameters . **Error! Bookmark not defined.**

**References**....................................................................... Error! Bookmark not defined.

### Appendix A. Sampling and data collection

From January 2020 through August 2021 we collected data from three villages in West Khasi Hills (WK1, WK2, WK3), three in West Jaintia Hills (WJ1, WJ2, WJ3) and four in South Garo Hills (SG1, SG2, SG3, SG4), as these were among the most isolated areas of Meghalaya with high malaria incidence. Villages were selected based on their manageable size (<500 eligible adults), their known willingness to participate to the study (avoiding villages where previous epidemiological studies encountered high rate of refusals, see Kessler et al., 2018), and their access by either car or foot.

A structured questionnaire was administered to every reachable adult (≥18 years old), resulting in information from 1,530 villagers residing in 766 households. Census data for every village was obtained from the Asha's register (**Figure A1**).

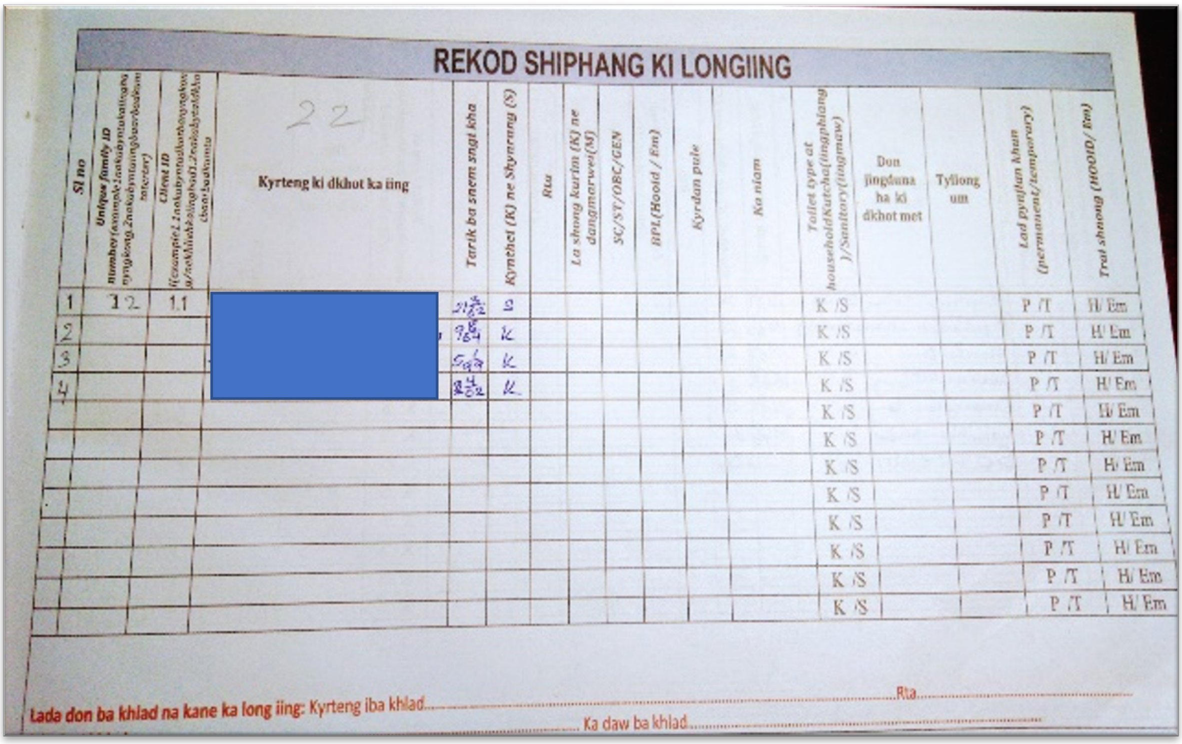

**Figure A1**. Example of Asha's register, which contains up to date record of who lives in each village in Meghalaya. Household members are reported in each page.

**Table A1** reports on the population of each village at the time of the data collection according to the Asha's register. The table further presents individual- and household-level response rates. The population figures include minors and people who recently moved out of the village but were still kept in the register. **Table A2** provides an overview of non-respondents by reason for non-response in each village. Whenever the information was available, we recorded the why eligible adults could not be interviewed. Reasons included being temporarily out farming, being temporarily out of the village, being unable to respond due to other reasons, having their door locked, or refusing to participate.

**Table A1.** Population size and response rates by village and in total.

| District | West Khasi Hills | | | West Jaintia Hills | | | South Garo Hills | | | | Total |
|---|---|---|---|---|---|---|---|---|---|---|---|
| Village | WK1 | WK2 | WK3 | WJ1 | WJ2 | WJ3 | SG1 | SG2 | SG3 | SG4 | |
| Population | 255 | 151 | 187 | 1206 | 1066 | 546 | 163 | 328 | 644 | 589 | 5135 |
| Eligible adults | 130 | 55 | 83 | 485 | 460 | 216 | 68 | 152 | 312 | 278 | 2239 |
| Adults interviewed | 102 | 48 | 73 | 317 | 293 | 115 | 44 | 117 | 209 | 212 | 1530 |
| % eligible adults interviewed | 78% | 87% | 88% | 65% | 64% | 53% | 65% | 77% | 67% | 76% | 68% |
| Eligible households | 69 | 25 | 31 | 226 | 190 | 98 | 30 | 56 | 106 | 121 | 952 |
| Households interviewed | 52 | 22 | 30 | 153 | 175 | 78 | 22 | 50 | 93 | 91 | 766 |
| % eligible households interviewed | 75% | 88% | 97% | 68% | 92% | 80% | 73% | 89% | 88% | 75% | 80% |

**Table A2.** Number of non-respondents by reasons of non-response in each village and in total.

| District | West Khasi Hills | | | West Jaintia Hills | | | South Garo Hills | | | | Total |
|---|---|---|---|---|---|---|---|---|---|---|---|
| Village | WK1 | WK2 | WK3 | WJ1 | WJ2 | WJ3 | SG1 | SG2 | SG3 | SG4 | |
| Eligible adults | 130 | 55 | 83 | 485 | 460 | 216 | 68 | 152 | 312 | 278 | 2239 |
| Temporarily in fields | 6 | 0 | 0 | 37 | 27 | 32 | 4 | 7 | 0 | 0 | 113 |
| Temporarily away | 13 | 5 | 3 | 33 | 31 | 18 | 6 | 10 | 35 | 14 | 168 |
| Unable to respond | 4 | 2 | 1 | 28 | 21 | 14 | 11 | 9 | 23 | 35 | 148 |
| Door locked | 0 | 0 | 0 | 6 | 4 | 2 | 0 | 0 | 5 | 0 | 17 |
| Refused to respond | 3 | 0 | 0 | 64 | 84 | 35 | 3 | 5 | 36 | 17 | 247 |
| Unknown reason | 2 | 0 | 6 | 0 | 0 | 0 | 0 | 4 | 4 | 0 | 16 |
| Total non-respondents | 28 | 7 | 10 | 168 | 167 | 101 | 24 | 35 | 103 | 66 | 709 |

The 1530 interviewed villagers named further 1195 people we could not reach for interviews for any of the reasons reported in Table 3. Of these people we do know their gender and the village they live in as we asked this information to the interviewees. We can thus compare the gender distribution of respondents and non-respondents. Of the people interviewed (1530) 57% were female, 43% were male, compared to the people named but not interviewed (1195) of which 47% were female, 53% were male. This means that we slightly capture more women than men in villages, possibly because women may be less frequently out farming. Looking at the people we interviewed (1530) 78% of men work in fields, compared to 64% of women.

Interviews were conducted in the appropriate local language (Khasi, Pnar or Garo) and translated into English by the interviewing team. Permission to conduct the study was granted by the Headman of each village, and all respondents also signed an individual informed consent form. The name of the village and information on individual participants was de-identified to protect anonymity.

Ethical approval for the study was obtained from the Institutional Review Boards (IRBs) of Martin Luther Christian University, Shillong, Meghalaya, India and New York University, New York, NY, USA.

### Appendix B. Questionnaire and variables

#### B.1 Use of preventive measures

Table B1 lists the questions we asked participants about their use of malaria preventive measures. Questions and response options are presented in English and Khasi, Pnar or Garo (depending on the tribe). In the analyses reported in the article, we did not utilise responses about the use of bednets including those without insecticide treatment (Q1), as nearly all interviewees said they used these. We also did not utilise responses about the use of mosquito mats, as none of the interviewees said they used these For the present paper, we recoded frequency of use of covering clothes (Q4) and insecticide cream (Q7) to use ("always", "sometimes" responses) and non-use ("rarely", "never" responses), in order to better match the response categories of the other items. **Table 2** reports the average number of preventive measures used in each village (range 0-8, average use in households and average use for individuals), while **Table 3** reports the percentage of individuals using each measure used in each village.

**Table B1.** Interview questions about the use of malaria preventive measures. Items and response options are described in English translation and in Khasi, Pnar or Garo.

| No. | Question (in English) | Response options | | |
|---|---|---|---|---|
| Q1 | Do you use a net at night? | Yes [ ] | No [ ] *(If no, go to Q4)* | |
| Q2 | If yes, is the net treated with insecticide (LLINS)? | Yes [ ] | No [ ] | Don't know [ ] |
| Q3 | If you do not use a net, what is the reason? | *[free response]* | | |
| Q4 | Do you cover your arms and legs during evening and early morning to prevent mosquito bites? | Always [ ]<br>Never [ ] | Sometimes [ ] | Rarely [ ] |
| Q5 | [If they work in fields:] Do you use boots when you go farming? | Yes [ ] | No [ ] | Don't know [ ] |
| Q6 | [If they work in fields:] Do you use gloves when you go farming? | Yes [ ] | No [ ] | Don't know [ ] |
| Q7 | Do you use insecticide cream to prevent mosquito bites? | Always [ ]<br>Never [ ] | Sometimes [ ] | Rarely [ ] |
| Q8 | Do you use any of the following to prevent mosquito bites? *(Tick all that apply)* | Use coils [ ]<br>Burn material (like neem leaves, cow dung) [ ]<br>Other (specify) [ ] | Use vaporizer [ ] | Mosquito mats [ ] |

**Table B2**. Average number of preventive measures used by households and individuals in each village.

| Village | Mean no. of measures used by households | Mean no. of measures used by individuals |
|---|---|---|
| WK1 | 2.8 | 2.8 |
| WK2 | 2.7 | 2.5 |

| | | |
|---|---|---|
| WK3 | 3.9 | 3.4 |
| WJ1 | 3.0 | 3.4 |
| WJ2 | 3.0 | 3.2 |
| WJ3 | 1.9 | 3.2 |
| SG1 | 2.3 | 2.4 |
| SG2 | 2.5 | 2.2 |
| SG3 | 2.9 | 2.7 |
| SG4 | 2.4 | 2.4 |
| Total | 2.8 | 2.9 |

**Table B3**: Percentage of interviewed individuals use of each preventive measure in each village. Cell colour represents level of use: higher levels marked by darker green.

| Village | LLINS | Covering clothes | Boots | Gloves | Insecticide cream | Coils | Vaporisers | Burning materials |
|---|---|---|---|---|---|---|---|---|
| WK1 | 79% | 65% | 26% | 21% | 13% | 38% | 4% | 31% |
| WK2 | 79% | 54% | 13% | 21% | 2% | 35% | 13% | 33% |
| WK3 | 89% | 73% | 29% | 16% | 23% | 58% | 12% | 37% |
| WJ1 | 98% | 78% | 49% | 3% | 9% | 52% | 22% | 30% |
| WJ2 | 99% | 69% | 24% | 4% | 15% | 77% | 3% | 26% |
| WJ3 | 91% | 79% | 18% | 0% | 22% | 75% | 0% | 36% |
| SG1 | 89% | 48% | 9% | 2% | 11% | 68% | 0% | 11% |
| SG2 | 100% | 50% | 4% | 0% | 3% | 33% | 0% | 27% |
| SG3 | 95% | 36% | 11% | 2% | 25% | 88% | 15% | 1% |
| SG4 | 94% | 48% | 3% | 1% | 6% | 85% | 1% | 0% |
| Total | 91% | 60% | 19% | 7% | 13% | 61% | 7% | 23% |

#### B.2 Individual characteristics

Table B4 lists the interview questions that were asked related to individuals' background, socio-demographic status, and roles in the household. We used only a binary indicator variable of occupation (Q14), representing whether a participant worked in fields (responses "cultivator" and "agricultural labourer") or not (all other responses). For the reported analyses, we recoded age to an ordinal variable with three categories: 18-29 years, 30-49 years, and 50 years and above. **Tables B5-B7** report the distribution of each individual characteristics in each village.

**Table B4.** Interview questions about individual characteristics. Items and response options are described in English translation and in Khasi, Pnar or Garo

| No. | Question (in English) | Response options | |
|---|---|---|---|
| Q9 | Gender | Male [ ] | Female [ ] |
| Q10 | Age | *[age recorded in years]* | |
| Q11 | Are you the head of the household? | Yes [ ] | No [ ] |

| Q12 | Are you in charge of looking after other members of the family when they are sick? | Yes [ ] | No [ ] |
|---|---|---|---|
| Q13 | Highest level of education | No schooling [ ]<br>Primary (1-5) [ ]<br>Secondary (X) [ ]<br>Graduate [ ]<br>Diploma [ ] | Below primary [ ]<br>Middle (6-8) [ ]<br>Higher Secondary (XII) [ ]<br>Post graduate [ ]<br>Other (specify) [ ] |
| Q14 | Usual occupation | Cultivator (independent farmer) [ ]<br>Agricultural labourer [ ]<br>Salaried service [ ]<br>Housewife [ ]<br>Child, not in school [ ]<br>Other (specify) [ ] | <br>Daily wage/labour [ ]<br>Self-employed/trade [ ]<br>Student [ ]<br>None [ ] |

**Table B5**: Percentage of respondents who are female, heads of households, carer for a sick person, and working in fields in each village.

| Village | Female | Head of household | Carer for a sick person | Work in fields |
|---|---|---|---|---|
| WK1 | 57% | 47% | 77% | 83% |
| WK2 | 50% | 54% | 69% | 79% |
| WK3 | 53% | 36% | 66% | 48% |
| WJ1 | 64% | 37% | 86% | 80% |
| WJ2 | 63% | 43% | 89% | 63% |
| WJ3 | 72% | 38% | 86% | 65% |
| SG1 | 52% | 66% | 89% | 77% |
| SG2 | 41% | 63% | 94% | 87% |
| SG3 | 51% | 58% | 96% | 46% |
| SG4 | 52% | 75% | 97% | 77% |
| Total | 56% | 52% | 85% | 71% |

**Table B6**: Distribution of age in each village.

| Village | 18-29 years | 30-49 years | Above 50 years |
|---|---|---|---|
| WK1 | 38% | 43% | 19% |
| WK2 | 46% | 35% | 19% |
| WK3 | 37% | 45% | 16% |
| WJ1 | 38% | 45% | 17% |
| WJ2 | 38% | 43% | 19% |
| WJ3 | 38% | 48% | 13% |
| SG1 | 61% | 32% | 7% |
| SG2 | 38% | 48% | 15% |
| SG3 | 36% | 56% | 8% |
| SG4 | 40% | 42% | 17% |

| | | | |
|---|---|---|---|
| Total | 41% | 44% | 15% |

**Table B7**: Distribution of educational background in each village.

| Village | No schooling | Below primary | Primary | Middle | Secondary | Higher secondary | Graduate and above |
|---|---|---|---|---|---|---|---|
| WK1 | 41% | 19% | 27% | 12% | 0% | 0% | 1% |
| WK2 | 38% | 15% | 31% | 2% | 2% | 6% | 6% |
| WK3 | 12% | 5% | 22% | 19% | 12% | 16% | 11% |
| WJ1 | 46% | 11% | 30% | 8% | 3% | 2% | 0% |
| WJ2 | 40% | 7% | 28% | 10% | 7% | 6% | 1% |
| WJ3 | 57% | 13% | 16% | 4% | 7% | 2% | 1% |
| SG1 | 32% | 9% | 11% | 32% | 16% | 0% | 0% |
| SG2 | 68% | 5% | 6% | 12% | 9% | 0% | 0% |
| SG3 | 25% | 8% | 9% | 25% | 23% | 6% | 2% |
| SG4 | 45% | 4% | 11% | 14% | 21% | 5% | 0% |
| Total | 40% | 10% | 19% | 14% | 10% | 4% | 2% |

#### B.3 Social networks and households

Health-related discussion networks were queried by two interview questions, as presented in **Table B8**. We use information about discussion ties within one's village (Q15) to define the network-related variables related to network size, opinion leaders, and network exposure, as described in the main text. As people mentioned in this question were living in the same village as the respondent, we could match their names with our participant data and create village-level social networks. These networks thus only contain information about the discussion ties reported between our participants. The question about out-of-village ties (Q16) is the basis for the variable of network size outside of one's village, which we define as the number of people mentioned by each respondent.

**Table B9** reports the descriptive statistics of the village-level discussion networks constructed from responses to question Q15 in **Table B8**. The table presents the following statistics for each village:

1. Number of nodes: the number of participants (who could name and be named as discussion partners)
2. Number of ties: the total number of discussion ties reported nodes
3. Average degree: the average number of discussion ties reported by a node
4. Indegree centralization: a normalized measure of the skewness of the distribution of the number of incoming ties of nodes (theoretical range: 0-1; smaller values reflect that the number of ties are close to equally distributed across nodes; larger values mean one or a few actors receive all of the ties)
5. Number of strong components: count of strong components in the network; a strong component is a set of nodes in which every node is reachable from every other node

by a directed path in the graph; e.g., node C is reachable from node A if A reports B and B reports C as a discussion partner

6. Average distance: the average length of the shortest paths connecting each pair of reachable nodes
7. Standard deviation of distances: the standard deviation of the shortest path lengths between nodes
8. Diameter: the length of the longest shortest path in the graph

**Table B10** presents village-level descriptive statistics for the number of reported discussion ties outside of one's own village.

Further, in each village we interviewed the Asha, and where available the Traditional Healer. We consider these individuals as important experts and opinion leaders on health-related matters, including malaria prevention, in the studied communities. **Table B11** reports the percentage of individuals who talk to the Asha or the Traditional Healer, as well as the percentage of household where at least one individual talks to the Asha or the Traditional Healer.

**Table B8.** Interview questions about health-related discussion networks. Items and response options are described in English translation and in Khasi, Pnar or Garo.

| No. | Question (in English) | Response options |
|---|---|---|
| Q15 | Please name the people in your village you talk to about health-related matters. Please indicate, if you know, the family name and their nick name. | *[free response, names recorded]* |
| Q16 | Are there any other people outside your village you talk to about health-related matters? | *[free response, names recorded]* |

**Table B9.** Descriptive statistics of health-related discussion networks in each village. The intuitive definition of each statistic is presented in this text; Avg. = Average, Std. = Standard error.

| Village | # of nodes | # of ties | Avg. degree | Indegree centralization | # of components | Avg. distance | Std. distance | Diameter |
|---|---|---|---|---|---|---|---|---|
| WK1 | 102 | 276 | 2.71 | 0.32 | 18 | 5.29 | 2.48 | 13 |
| WK2 | 48 | 105 | 2.19 | 0.21 | 20 | 3.80 | 1.87 | 10 |
| WK3 | 73 | 313 | 4.29 | 0.60 | 11 | 3.50 | 1.43 | 7 |
| WJ1 | 317 | 648 | 2.04 | 0.57 | 204 | 3.84 | 2.52 | 15 |
| WJ2 | 293 | 642 | 2.19 | 0.38 | 166 | 2.97 | 1.65 | 10 |
| WJ3 | 115 | 212 | 1.84 | 0.42 | 62 | 4.43 | 2.81 | 13 |
| SG1 | 44 | 111 | 2.52 | 0.49 | 17 | 1.97 | 0.99 | 5 |
| SG2 | 117 | 309 | 2.64 | 0.67 | 39 | 5.71 | 2.88 | 14 |
| SG3 | 209 | 276 | 1.32 | 0.20 | 137 | 1.53 | 0.73 | 4 |
| SG4 | 212 | 473 | 2.23 | 0.52 | 106 | 1.77 | 0.89 | 6 |

**Table B10.** Descriptive statistics of participants' discussion networks outside of their own village. Avg. = Average, Std. = Standard error.

| Avg. no. ties out of village | Std. no. ties out of village |
|---|---|
| 1.99 | 1.73 |
| 1.98 | 1.76 |
| 2.56 | 1.74 |
| 0.90 | 1.06 |
| 1.11 | 1.35 |
| 1.27 | 1.12 |
| 1.33 | 1.04 |
| 0.51 | 0.87 |
| 0.45 | 0.71 |
| 1.06 | 1.23 |

**Table B11**. Percentage of individuals and households talking to the Asha or the Traditional Healer about health-related matters in each village.

| Village | % participants talk to Asha | % participants talk to Healer | % households talk to Asha | % households talk to Healer |
|---|---|---|---|---|
| WK1 | 34% | 4% | 31% | 5% |
| WK2 | 25% | 23% | 33% | 27% |
| WK3 | 64% | 7% | 64% | 10% |
| WJ1 | 57% | 0% | 50% | 0% |
| WJ2 | 38% | 13% | 40% | 18% |
| WJ3 | 43% | 0% | 20% | 0% |
| SG1 | 52% | 0% | 45% | 0% |
| SG2 | 68% | 0% | 74% | 0% |
| SG3 | 21% | 0% | 28% | 0% |
| SG4 | 53% | 0% | 58% | 0% |
| Total | 45% | 5% | 44% | 6% |

Combining network information (Q15) with data on measure use (Q1-Q8), we compute the number of people each individual talk to who use each preventive measure. This constitutes what we call network exposure to preventive measures (Valente 1996). For example, **Figure B1** illustrates the percentage of exposure to the use of coils in WK2. It depicts the social network of whom villagers talk to regarding health-related issues, and who uses coils. In this case, network exposure may vary between 0% (none of the people named adopted) to 100% (everyone named adopted). To measure the exposure within the household, we compute how many of the other household members each person lives with use each preventive measure in a similar way. These definitions of exposure are used in the logistic regression analyses reported in the main text and in Appendix D of this document. Exposure is defined slightly differently in the network models, as discussed in the main text and in Appendix E here. **Table B12-13** report the average and median percentage of network exposure for each preventive measure in each village. **Table B14-15** present the average and median percentage of exposure within households.

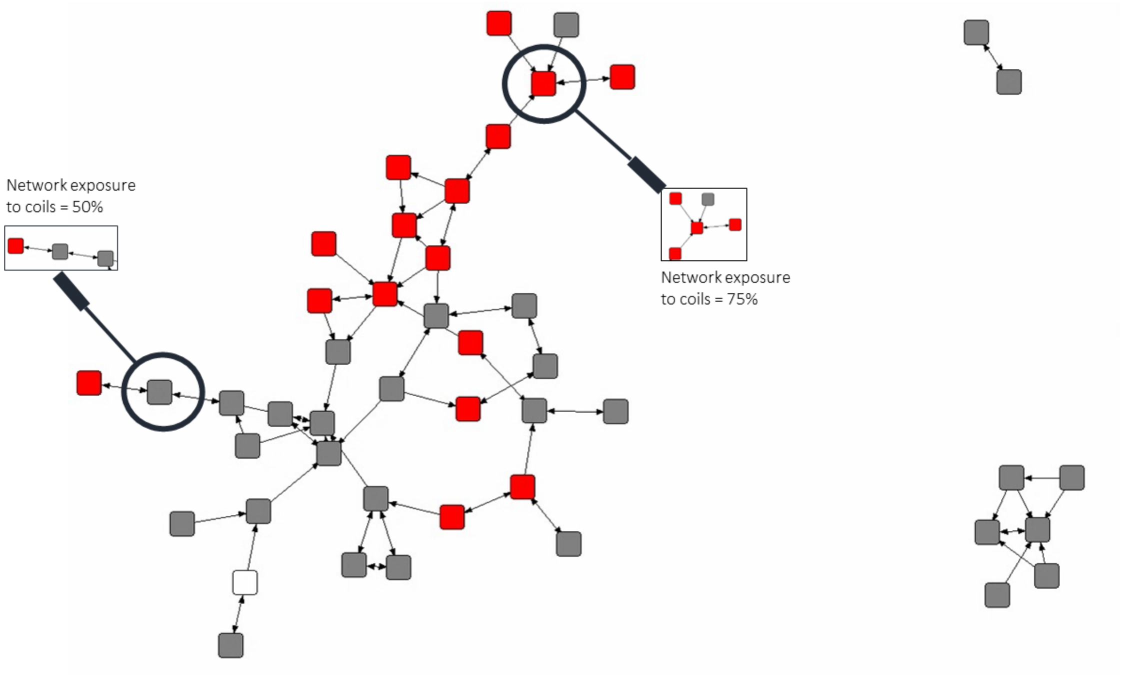

**Figure B1.** Visualisation of the exposure to the use of coils in the discussion network of village WK2. Nodes represent interviewed villagers; lines represent whom the villagers talk to about health-related matters; red nodes indicate villagers who use coils; grey nodes indicate villagers who do not use coils; network exposure to coils of the node highlighted at the top right = 75%; network exposure to coils of the node highlighted at the left: 50%.

**Table B12**. Average percentage of network exposure of individuals to each preventive measure in each village.

| Village | LLINS | Covering clothes | Boots | Gloves | Insecticide cream | Coils | Vaporisers | Burning materials |
|---|---|---|---|---|---|---|---|---|
| WK1 | 79% | 73% | 20% | 29% | 8% | 32% | 2% | 38% |
| WK2 | 77% | 54% | 14% | 28% | 3% | 37% | 19% | 28% |
| WK3 | 90% | 79% | 24% | 11% | 21% | 49% | 10% | 29% |
| WJ1 | 93% | 80% | 62% | 1% | 38% | 63% | 44% | 20% |
| WJ2 | 90% | 72% | 33% | 4% | 30% | 76% | 2% | 35% |
| WJ3 | 92% | 60% | 12% | 0% | 17% | 52% | 0% | 25% |
| SG1 | 82% | 65% | 9% | 3% | 33% | 81% | 0% | 11% |

| | | | | | | | | |
|---|---|---|---|---|---|---|---|---|
| SG2 | 98% | 60% | 3% | 0% | 2% | 45% | 0% | 20% |
| SG3 | 83% | 28% | 6% | 2% | 20% | 78% | 12% | 0% |
| SG4 | 92% | 52% | 2% | 1% | 25% | 86% | 1% | 0% |

**Table B13**. Median percentage of network exposure of individuals to each preventive measure in each village.

| Village | LLINS | Covering clothes | Boots | Gloves | Insecticide cream | Coils | Vaporisers | Burning materials |
|---|---|---|---|---|---|---|---|---|
| WK1 | 100% | 100% | 0% | 25% | 0% | 25% | 0% | 40% |
| WK2 | 100% | 50% | 0% | 12% | 0% | 33% | 0% | 0% |
| WK3 | 100% | 100% | 25% | 0% | 20% | 50% | 0% | 25% |
| WJ1 | 100% | 100% | 67% | 0% | 33% | 67% | 50% | 0% |
| WJ2 | 100% | 100% | 33% | 0% | 25% | 100% | 0% | 33% |
| WJ3 | 100% | 67% | 0% | 0% | 0% | 50% | 0% | 0% |
| SG1 | 100% | 67% | 0% | 0% | 25% | 100% | 0% | 0% |
| SG2 | 100% | 67% | 0% | 0% | 0% | 50% | 0% | 0% |
| SG3 | 100% | 0% | 0% | 0% | 0% | 100% | 0% | 0% |
| SG4 | 100% | 50% | 0% | 0% | 20% | 100% | 0% | 0% |

**Table B14**. Average percentage of network exposure of households for each preventive measure in each village.

| Village | LLINS | Covering clothes | Boots | Gloves | Insecticide cream | Coils | Vaporisers | Burning materials |
|---|---|---|---|---|---|---|---|---|
| WK1 | 73% | 60% | 26% | 23% | 14% | 45% | 6% | 33% |
| WK2 | 74% | 57% | 27% | 43% | 2% | 44% | 9% | 42% |
| WK3 | 83% | 72% | 49% | 24% | 32% | 63% | 14% | 55% |
| WJ1 | 75% | 65% | 50% | 3% | 11% | 46% | 20% | 28% |
| WJ2 | 82% | 64% | 28% | 6% | 18% | 68% | 5% | 29% |
| WJ3 | 52% | 46% | 14% | 0% | 15% | 39% | 0% | 20% |
| SG1 | 77% | 51% | 9% | 2% | 12% | 63% | 0% | 12% |
| SG2 | 90% | 59% | 10% | 0% | 7% | 47% | 0% | 38% |
| SG3 | 86% | 47% | 13% | 1% | 36% | 83% | 20% | 1% |
| SG4 | 88% | 56% | 7% | 1% | 8% | 83% | 1% | 0% |

**Table B15**. Median percentage of exposure within households to each preventive measure in each village.

| Village | LLINS | Covering clothes | Boots | Gloves | Insecticide cream | Coils | Vaporisers | Burning materials |
|---|---|---|---|---|---|---|---|---|
| WK1 | 100% | 100% | 0% | 0% | 0% | 0% | 0% | 0% |
| WK2 | 100% | 100% | 0% | 0% | 0% | 0% | 0% | 0% |

| | | | | | | | | |
|---|---|---|---|---|---|---|---|---|
| WK3 | 100% | 100% | 0% | 0% | 0% | 100% | 0% | 100% |
| WJ1 | 100% | 100% | 100% | 0% | 0% | 0% | 0% | 0% |
| WJ2 | 100% | 100% | 0% | 0% | 0% | 100% | 0% | 0% |
| WJ3 | 100% | 0% | 0% | 0% | 0% | 0% | 0% | 0% |
| SG1 | 100% | 100% | 0% | 0% | 0% | 100% | 0% | 0% |
| SG2 | 100% | 100% | 0% | 0% | 0% | 0% | 0% | 0% |
| SG3 | 100% | 0% | 0% | 0% | 0% | 100% | 0% | 0% |
| SG4 | 100% | 100% | 0% | 0% | 0% | 100% | 0% | 0% |

#### Appendix C. Visualisation of village-level discussion and measure use networks

**Figures C1-C10** visualise the multilevel network system of each village as a combination of two networks. The first is one mode network, connecting villagers to each other by their reported discussion ties. The second network connects villagers to any of the eight preventive measures that they use. This form of data is used in the Stochastic Actor-oriented Models reported in the main text and in Appendix E below.

In each figure here, villagers are represented by circle nodes and preventive measures by squared nodes. Thick red edges connect two villagers if at least one of them reports talking to the other about health-related matters (we consider tie directions in the statistical analyses). Thinner edges of various colour connect villagers to the preventive measures they use (edges are coloured by the preventive measure they connect to).

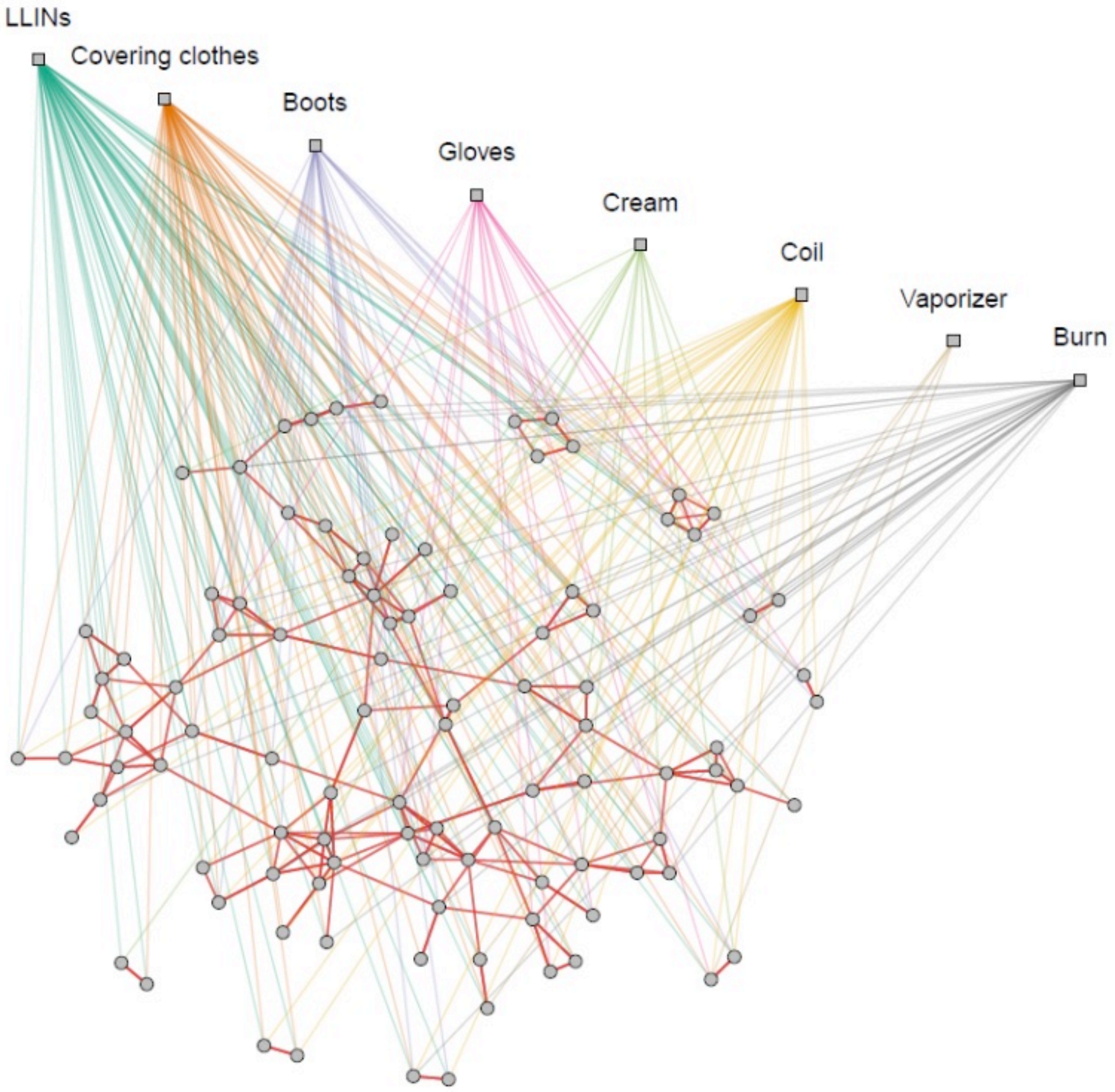

**Figure C1**. Visualisation of the multilevel network of discussion ties and measure use in WK1.

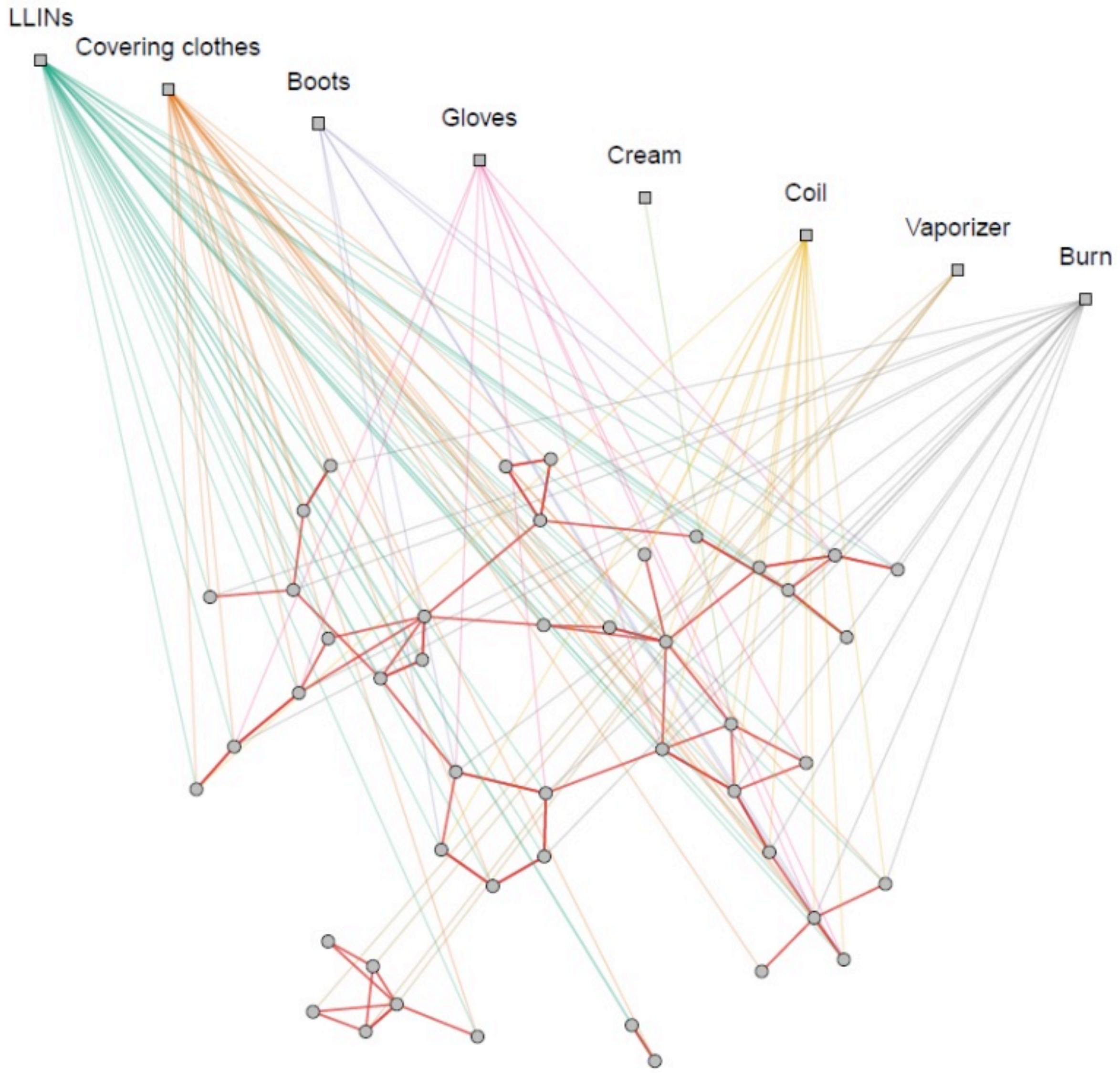

**Figure C2**. Visualisation of the multilevel network of discussion ties and measure use in WK2.

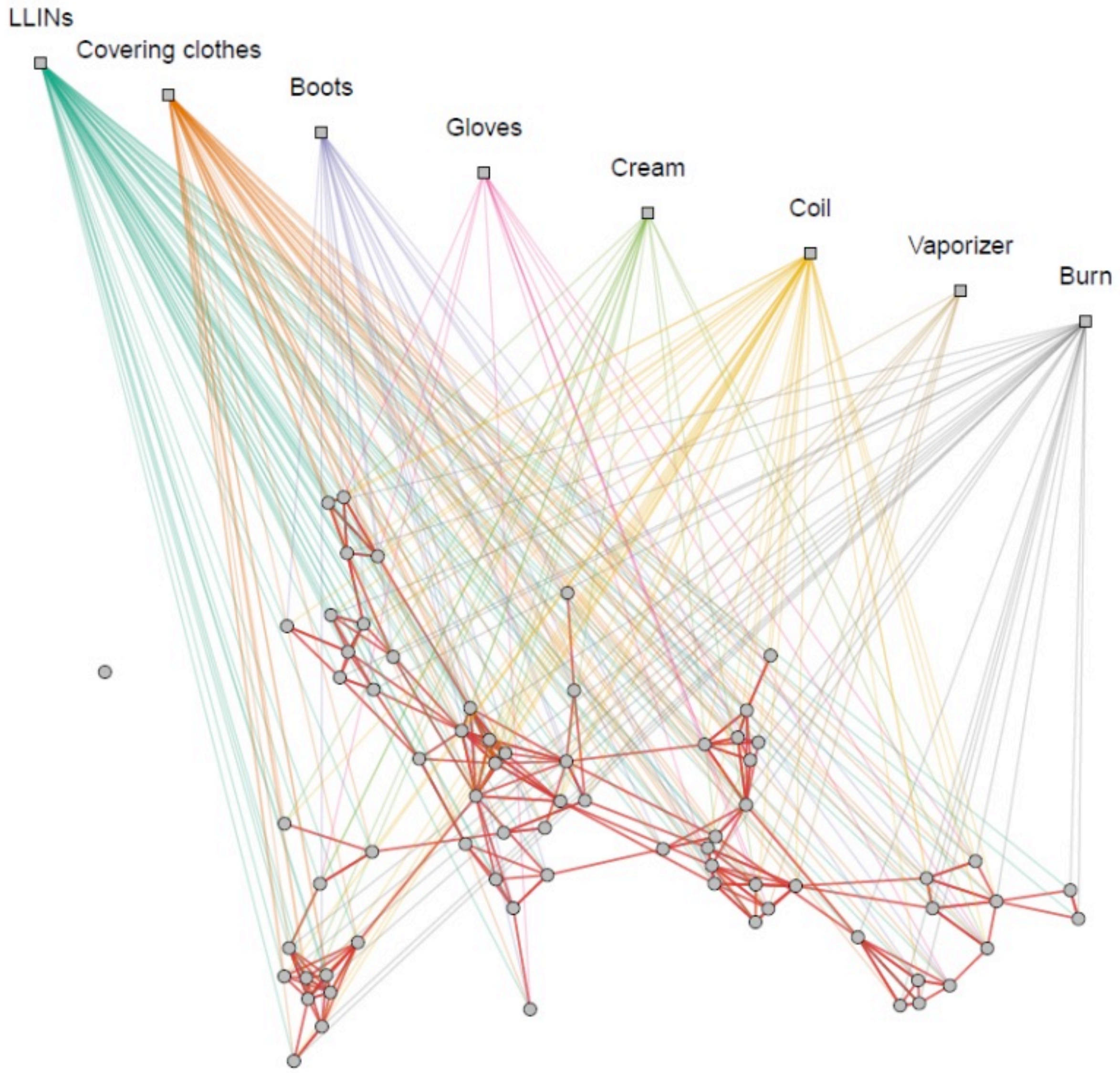

Figure C3. Visualisation of the multilevel network of discussion ties and measure use in WK3.

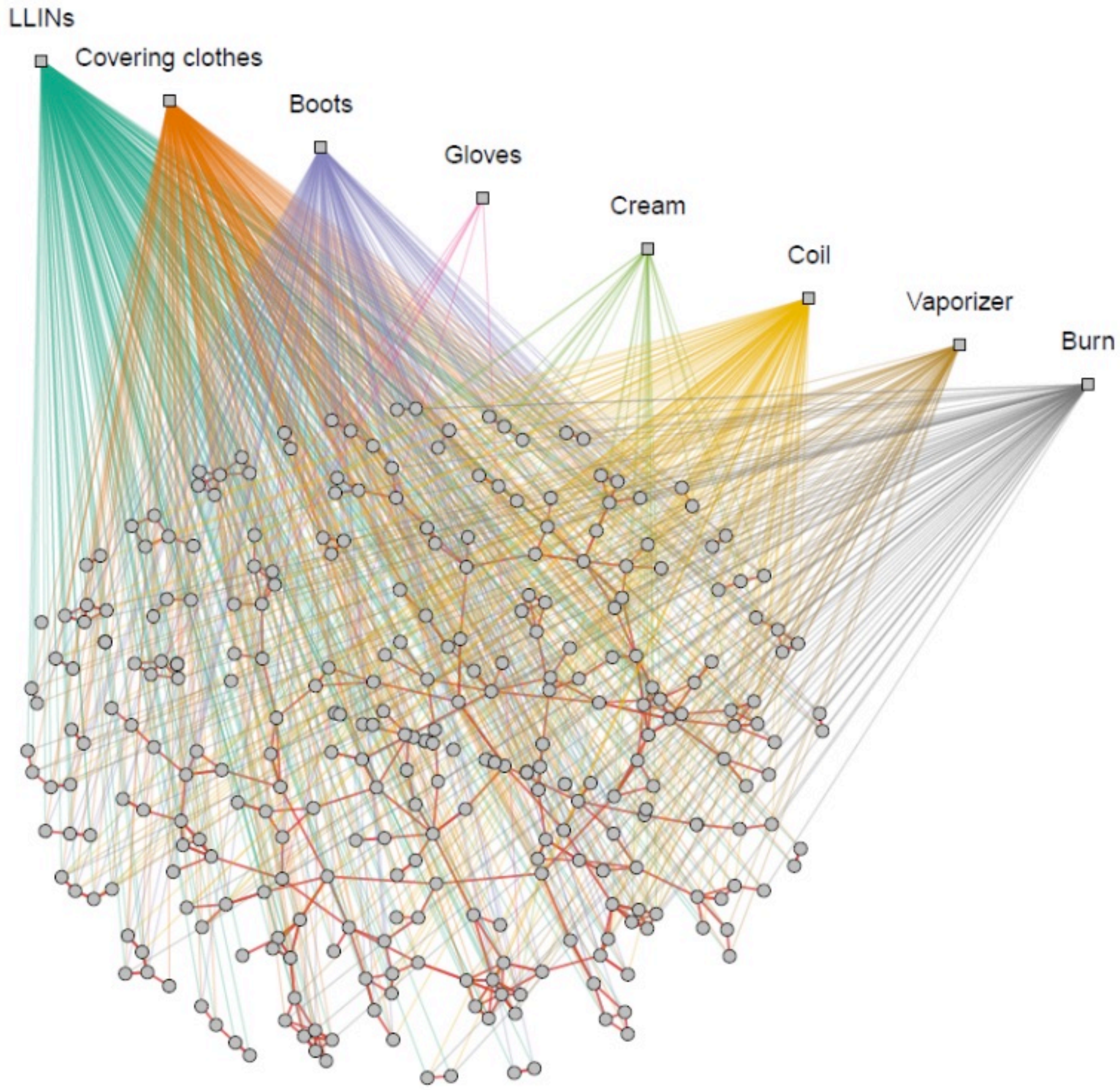

**Figure C4.** Visualisation of the multilevel network of discussion ties and measure use in WJ1.

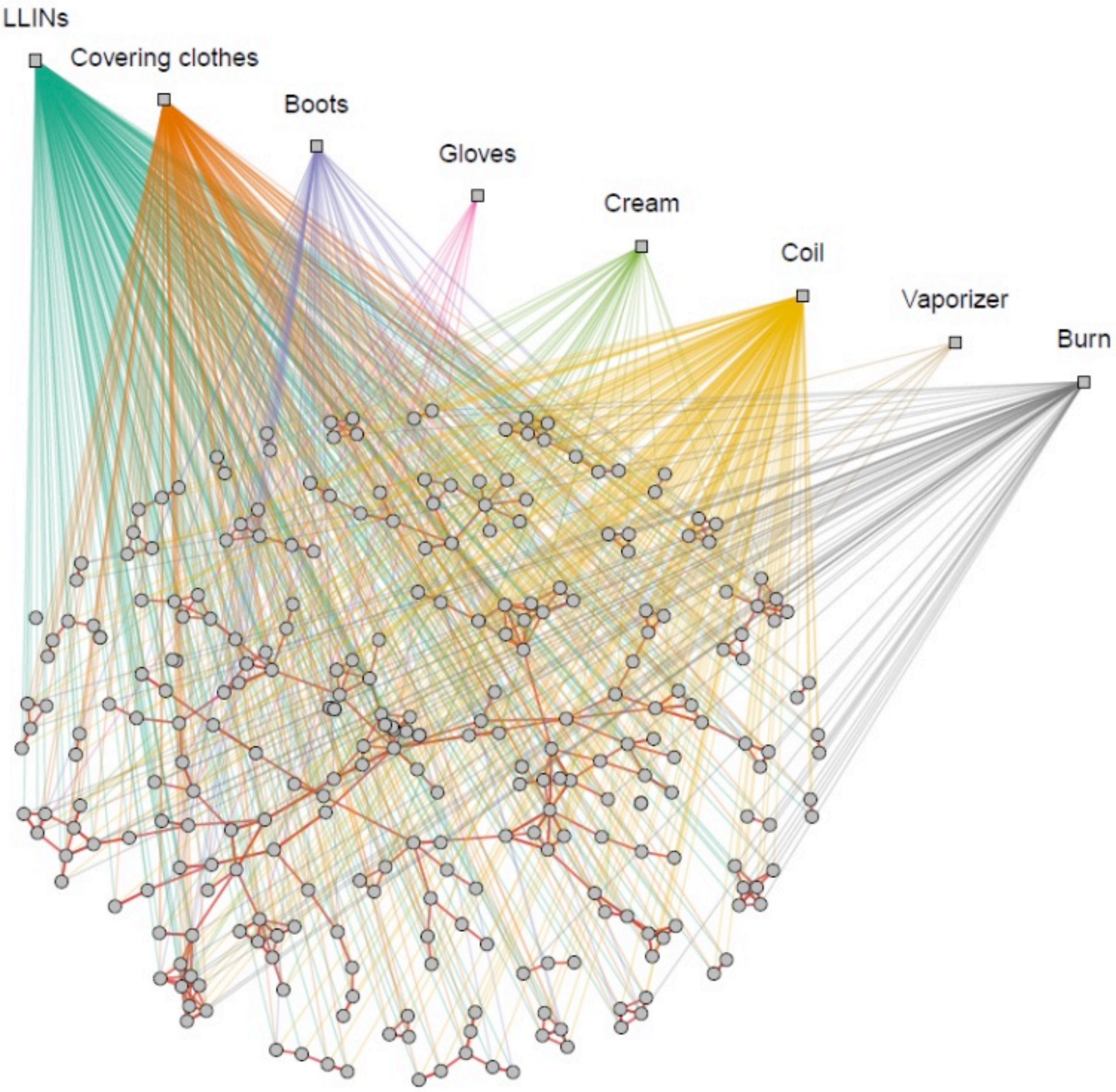

Figure C5. Visualisation of the multilevel network of discussion ties and measure use in WJ2.

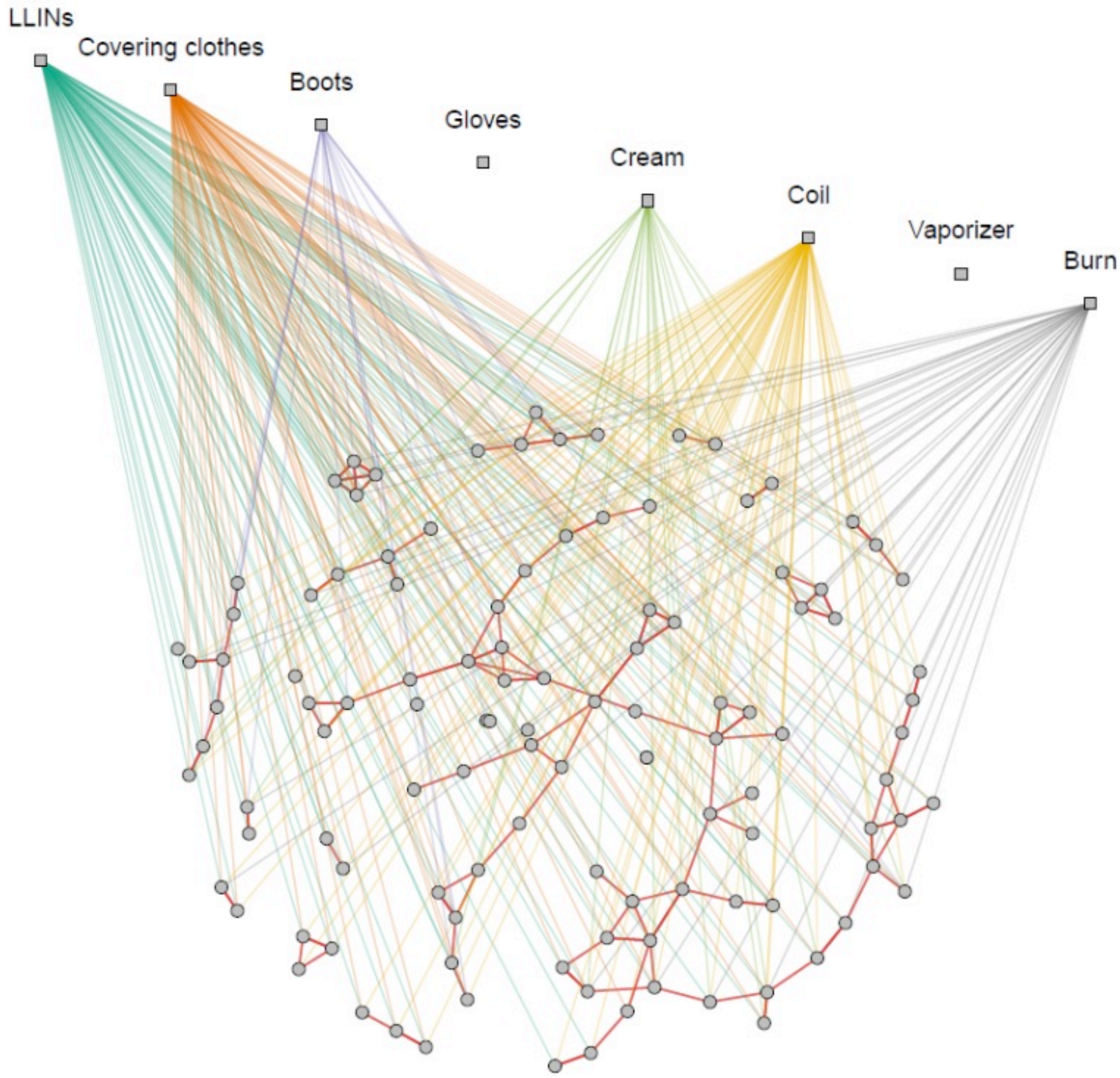

**Figure C6.** Visualisation of the multilevel network of discussion ties and measure use in WJ3.

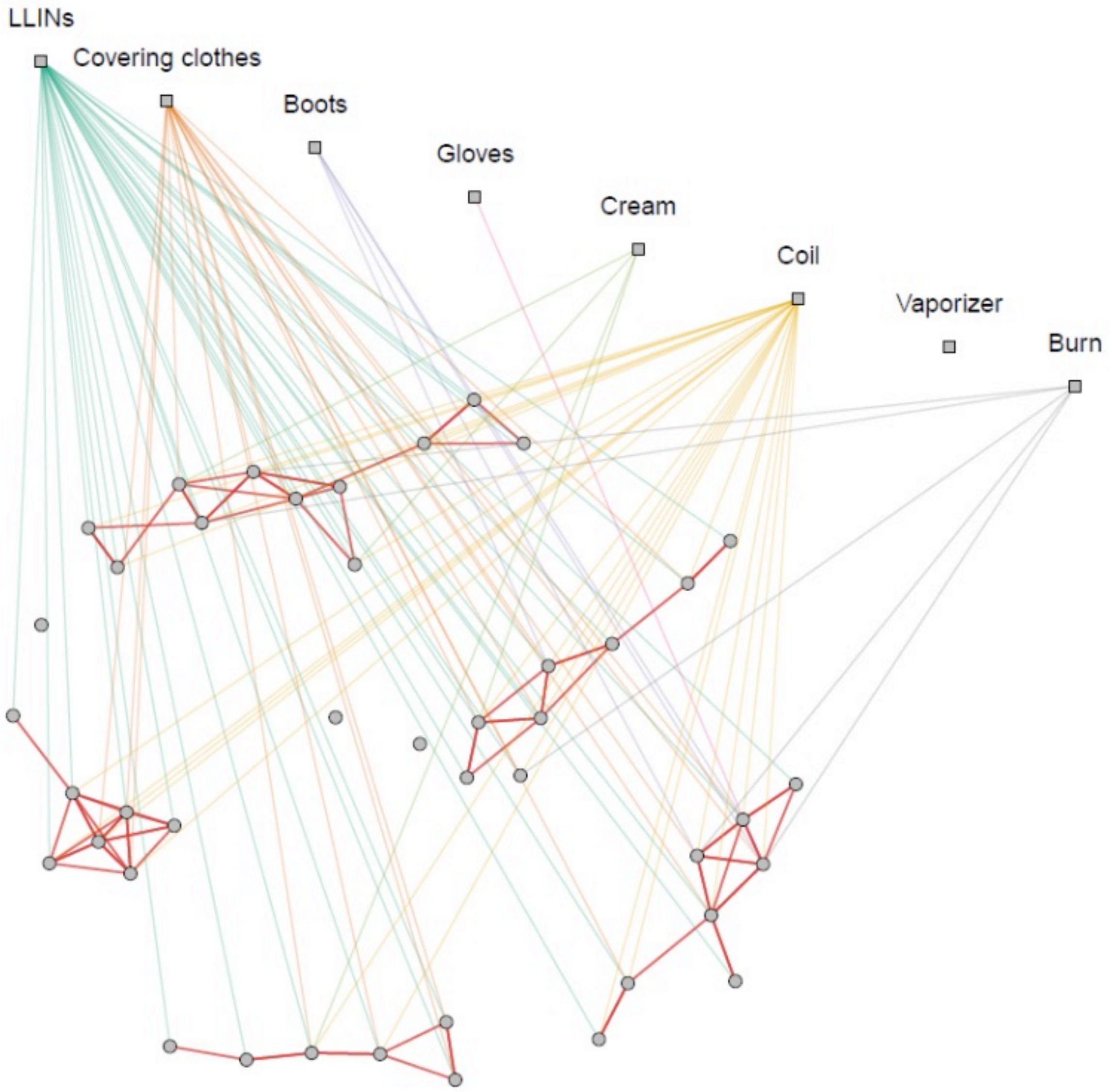

Figure C7. Visualisation of the multilevel network of discussion ties and measure use in SG1.

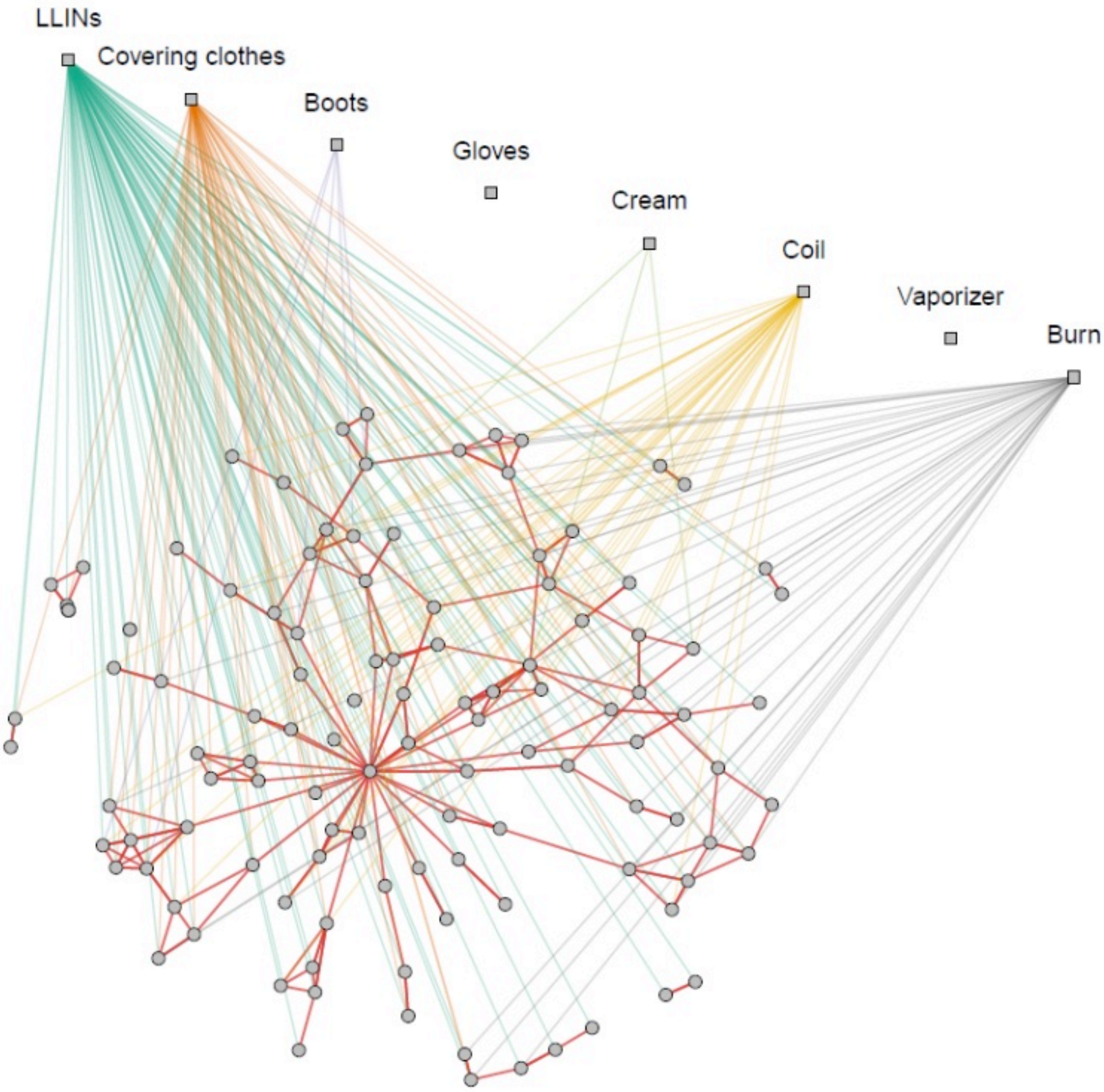

Figure C8. Visualisation of the multilevel network of discussion ties and measure use in SG2.

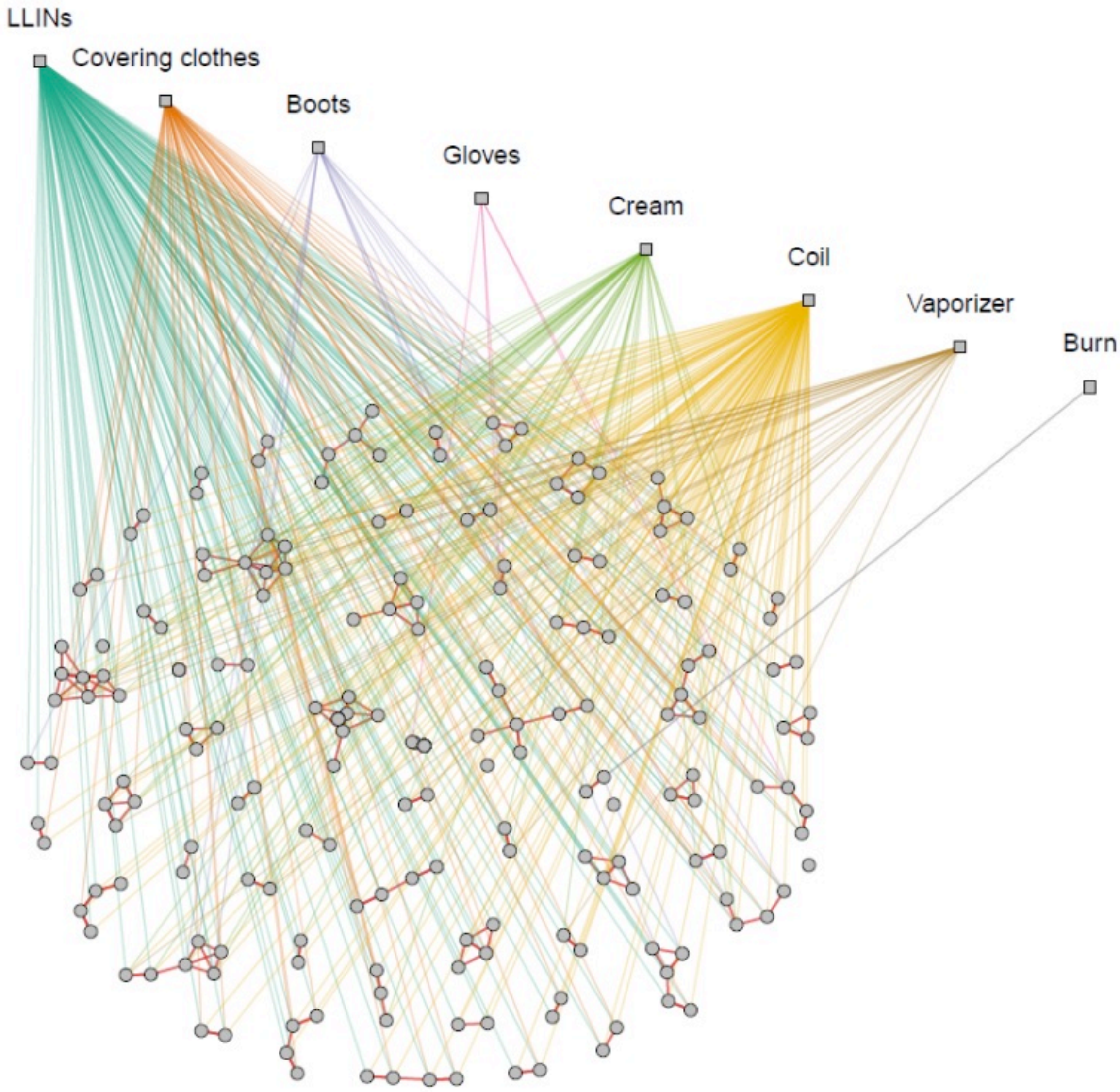

**Figure C9.** Visualisation of the multilevel network of discussion ties and measure use in SG3.

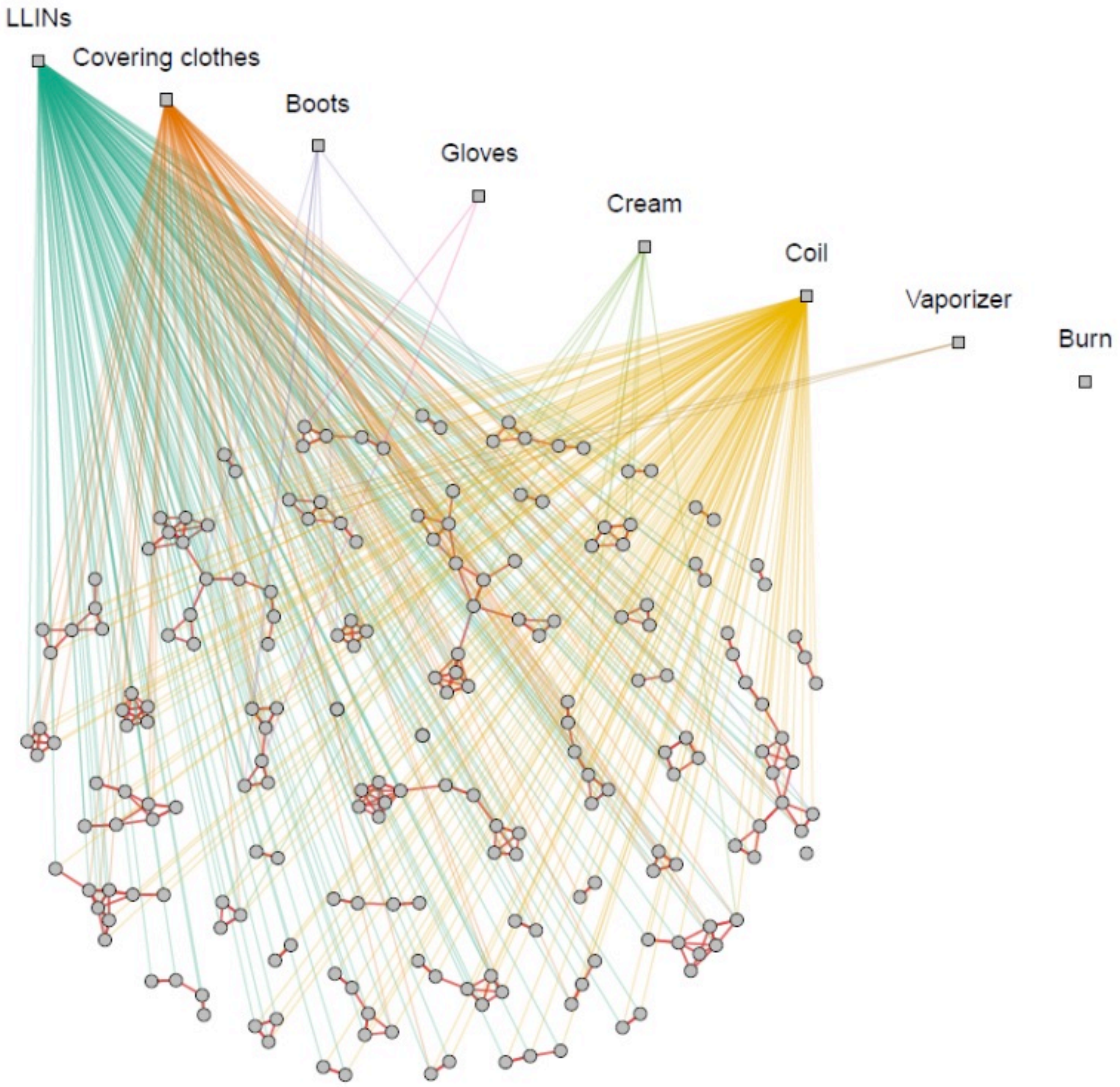

**Figure C10.** Visualisation of the multilevel network of discussion ties and measure use in SG4.

### Appendix D. Results of the analyses using logistic regression models

#### D.1 Model selection I: Backward variable selection in each village

**Table D1** presents the explanatory variables we considered for each dependent variable (preventive measure) and village when fitting our logit regression models. Due to the size of the villages and the often heavily skewed distribution of the dependent variables, most of the village-level datasets did not have sufficient statistical power to fit models that included all of these explanatory factors. To reduce the set of explanatory variables, we carried out a backward model selection procedure, based on estimate p-values, as described in the Methods section of the main text (Procedure 1, Stage I). **Tables D2-9** present the results of this process, one model for each village and each of the eight binary dependent variables.

**Table D1**. Description of explanatory variables considered in the logistic models.

| Explanatory Variable | Description |
|---|---|
| (1) Female | Respondent is female (1=yes, 0=no) |
| (2) Head of household | Respondent is head of their household (1=yes, 0=no) |
| (3) Carer for a sick person | Respondent is the carer for a sick person (1=yes, 0=no) |
| (4) Works in fields | Respondent works in fields (1=yes, 0=no) |
| (5) Age | Respondent's age in years (1=18-29, 2=30-49, 3=50+) |
| (6) Education | Respondent's level of education (1=none, …, 7=graduate) |
| (7) Talks to Asha | Respondent talks to their village's Asha (1=yes, 0=no) |
| (8) Talks to Healer | Respondent talks to their village's Healer (1=yes, 0=no) |
| (9) Network exposure | Percentage of users of a given measure among those the respondent talks to in the village (0-100%) |
| (10) Network outdegree in village | No. of people respondent talks to in their village |
| (11) Network outdegree out of village | No. of people respondent talks to outside their village |
| (12) Household talks to Asha | Someone in respondent's household talks to their village's Asha (1=yes, 0=no) |
| (13) Household talks to Healer | Someone in respondent's household talks to their village's Healer (1=yes, 0=no) |
| (14) Household exposure | Someone in respondent's household, excl. respondent, uses a given preventive measure (1=yes, 0=no) |

**Table D2:** Results of village-level backward model selection procedures for logistic regressions explaining the use of LLINs. Parameters are reported with standard errors (in parentheses), along with the number of observations (N) and goodness of fit statistics at the bottom of the table.

| Village | WK1 | WK2 | WK3 | WJ1 | WJ2 | WJ3 | SG1 | SG3 | SG4 |
|---|---|---|---|---|---|---|---|---|---|
| Intercept | -1.62* (0.77) | -0.81 (0.60) | 2.23*** (0.40) | 2.29** (0.75) | 4.98*** (0.71) | -0.94 (0.97) | 8.04* (3.34) | 1.82 (1.10) | -1.44 (0.79) |
| Female | | | | | | 1.76* (0.82) | | | |
| Head of household | | | | | | | | 2.06* (0.98) | |
| Carer for a sick person | | | | | | 2.63 ** (0.99) | | | |
| Works in fields | | | | | | | | | 1.85 ** (0.70) |
| Age | 1.58** (0.50) | | | | | | | | |
| Talks to Asha | 2.61* (1.07) | | | | | | | | |
| Network exposure | | | | 2.31* (0.96) | | | | 3.98*** (1.04) | 3.84*** (0.81) |
| Outdegree in village | | | | | | | -1.62* (0.79) | | |
| Household talks to Asha | | | | | | 2.74* (1.38) | | | |
| Household exposure | | 4.34*** (1.18) | | | | | | -2.56* (1.30) | |
| N | 102 | 48 | 72 | 317 | 293 | 113 | 41 | 198 | 208 |
| AIC | 83.37 | 29.13 | 47.93 | 50.94 | 25.93 | 55.87 | 17.57 | 55.52 | 74.62 |
| BIC | 91.25 | 32.87 | 50.20 | 58.46 | 29.61 | 66.77 | 21.00 | 68.67 | 84.63 |
| Pseudo R2 | 0.36 | 0.61 | 0.00 | 0.09 | 0.00 | 0.29 | 0.43 | 0.33 | 0.34 |

*** $p < 0.001$; ** $p < 0.01$; * $p < 0.05$.

**Table D3:** Results of village-level backward model selection procedures for logistic regressions explaining the use of covering clothes. Parameters are reported with standard errors (in parentheses), along with the number of observations (N) and goodness of fit statistics at the bottom of the table.

| Village | WK1 | WK2 | WK3 | WJ1 | WJ2 | WJ3 | SG1 | SG2 | SG3 | SG4 |
|---|---|---|---|---|---|---|---|---|---|---|
| Intercept | -1.70*<br>(0.83) | -0.42<br>(0.59) | -0.46<br>(0.48) | -0.31<br>(0.58) | -0.01<br>(0.23) | 1.03<br>(0.73) | -4.00**<br>(1.24) | -1.62**<br>(0.50) | -0.04<br>(0.23) | -0.99<br>(0.85) |
| Female | 1.77*<br>(0.74) | | | | | -1.47*<br>(0.70) | | | | |
| Head of house-hold | 1.45*<br>(0.72) | | | | | | | | -1.31***<br>(0.32) | -1.50*<br>(0.61) |
| Works in fields | | | | 0.82*<br>(0.35) | | | | | | -1.21*<br>(0.59) |
| Age | | | | -0.50*<br>(0.22) | | | | | | |
| Edu-cation | | | | | | | | | | -0.29*<br>(0.14) |
| Talks to Asha | | | | -0.81*<br>(0.39) | | 2.74***<br>(0.75) | | | 0.86*<br>(0.38) | 1.94***<br>(0.59) |
| Talks to Healer | | | -3.20*<br>(1.29) | | | | | | | |
| Network exposure | | 2.16*<br>(0.86) | | 1.79***<br>(0.49) | | 2.87***<br>(0.86) | | | | |
| Network size in village | | | | | | -0.67*<br>(0.31) | 0.83*<br>(0.35) | 0.44**<br>(0.16) | | 0.76***<br>(0.21) |
| Network size out of village | | | | 0.35*<br>(0.16) | 0.31**<br>(0.12) | | 1.37*<br>(0.54) | 0.96**<br>(0.30) | | 0.83***<br>(0.21) |
| House-hold talks to healer | | -2.20*<br>(0.91) | | | | | | | | |
| House-hold exposure | 1.14*<br>(0.46) | | 2.90***<br>(0.71) | 1.29***<br>(0.33) | 0.86 *<br>(0.26) | | | | | |
| N | 102 | 48 | 72 | 317 | 293 | 113 | 40 | 117 | 195 | 204 |
| AIC | 130.03 | 53.71 | 61.69 | 287.06 | 348.31 | 104.95 | 40.27 | 147.74 | 240.85 | 150.98 |
| BIC | 140.53 | 59.32 | 68.52 | 313.37 | 359.35 | 118.58 | 45.34 | 156.02 | 250.66 | 174.20 |
| Pseudo R2 | 0.13 | 0.43 | 0.46 | 0.28 | 0.09 | 0.25 | 0.54 | 0.21 | 0.14 | 0.68 |

*** $p < 0.001$; ** $p < 0.01$; * $p < 0.05$.

**Table D4:** Results of village-level backward model selection procedures for logistic regressions explaining the use of boots. Parameters are reported with standard errors (in parentheses), along with the number of observations (N) and goodness of fit statistics at the bottom of the table.

| Village | WK1 | WK2 | WK3 | WJ1 | WJ2 | WJ3 | SG1 | SG2 | SG3 | SG4 |
|---|---|---|---|---|---|---|---|---|---|---|
| Intercept | -0.61 (0.34) | -3.82 * (1.49) | -0.41 (0.36) | -0.71 (0.52) | -1.42*** (0.27) | -0.22 (0.39) | -2.51*** (0.60) | -4.94*** (1.18) | -1.43** (0.45) | -5.63*** (1.31) |
| Female | -2.16*** (0.61) | | -1.70** (0.63) | -1.32*** (0.30) | -0.98** (0.30) | -1.81*** (0.54) | | | | |
| Works in fields | | | | 0.98** (0.34) | | | | | | |
| Age | | | | -0.42* (0.19) | | | | | | |
| Edu-cation | | | | | | | | 0.74* (0.31) | | |
| Talks to Asha | | 3.00* (1.46) | | | | | | | | |
| Network exposure | | 4.20* (2.04) | | 1.64*** (0.38) | 1.44** (0.46) | | | | 2.04* (1.03) | |
| Network size in village | | | | | | | | | -1.11** (0.39) | |
| Network size out of village | | | | | | | | | | 0.94* (0.45) |
| House-hold talks to Healer | | | 2.46* (0.98) | | | | | | | |
| House-hold exposure | 1.92** (0.63) | | | 0.95*** (0.27) | 0.80* (0.34) | | | | 1.86** (0.68) | |
| N | 102 | 28 | 72 | 317 | 293 | 96 | 40 | 102 | 194 | 163 |
| AIC | 101.71 | 25.77 | 78.45 | 365.84 | 292.03 | 90.60 | 23.31 | 37.38 | 115.78 | 28.88 |
| BIC | 109.59 | 29.77 | 85.28 | 388.39 | 306.75 | 95.73 | 25.00 | 42.63 | 128.85 | 35.06 |
| Pseudo R2 | 0.29 | 0.44 | 0.26 | 0.32 | 0.17 | 0.18 | 0.00 | 0.19 | 0.31 | 0.18 |

*** $p < 0.001$; ** $p < 0.01$; * $p < 0.05$.

**Table D5:** Results of village-level backward model selection procedures for logistic regressions explaining the use of gloves. Parameters are reported with standard errors (in parentheses), along with the number of observations (N) and goodness of fit statistics at the bottom of the table.

| Village | WK1 | WK2 | WK3 | WJ1 | WJ2 | SG1 | SG3 | SG4 |
|---|---|---|---|---|---|---|---|---|
| Intercept | -2.68 ** (0.87) | -1.87 * (0.76) | -0.35 (0.85) | -3.94 *** (0.41) | -3.65 *** (0.38) | -21.57 (N/A)+ | -12.12 ** (3.73) | -4.38 *** (0.71) |
| Age | | | | | | | 1.85 * (0.94) | |
| Edu-cation | 0.79 ** (0.26) | | | | | | 1.15 * (0.49) | |
| Network exposure | | | | 5.03 ** (1.55) | | | | |
| Network size in village | -0.59 * (0.25) | | -0.47 * (0.23) | | | | | |
| Network size out of village | 0.40 * (0.19) | | | | | | | |
| House-hold exposure | | 2.56 ** (0.98) | 1.70 * (0.73) | | 2.47 *** (0.69) | | | |
| N | 102 | 27 | 72 | 317 | 293 | 39 | 187 | 161 |
| AIC | 90.60 | 31.06 | 61.48 | 69.79 | 87.81 | 2.00 | 41.29 | 23.53 |
| BIC | 101.10 | 33.65 | 68.31 | 77.31 | 95.17 | 3.66 | 50.98 | 26.61 |
| Pseudo R2 | 0.29 | 0.37 | 0.21 | 0.13 | 0.12 | 0.00 | 0.26 | 0.00 |

*** p < 0.001; * p < 0.01; * p < 0.05; + large (>1000) std. error suggesting uncertainty cannot be precisely estimated

**Table D6:** Results of village-level backward model selection procedures for logistic regressions explaining the use of insecticide cream. Parameters are reported with standard errors (in parentheses), along with the number of observations (N) and goodness of fit statistics at the bottom of the table.

| Village | WK1 | WK2 | WK3 | WJ1 | WJ2 | WJ3 | SG1 | SG2 | SG3 | SG4 |
|---|---|---|---|---|---|---|---|---|---|---|
| Intercept | -1.88*** | -3.81*** | -1.41** | -2.80*** | -2.46*** | -1.41* | -1.97*** | -7.16** | -3.64*** | -0.31 |
| | (0.30) | (1.01) | (0.48) | (0.26) | (0.43) | (0.63) | (0.48) | (2.44) | (0.98) | (1.17) |
| Head of house-hold | | | | | | | | | 1.97*** | |
| | | | | | | | | | (0.54) | |
| Carer for a sick person | | | | | | -1.49* | | | | |
| | | | | | | (0.67) | | | | |
| Works in fields | | | -1.39* | | | | | | | -2.36* |
| | | | (0.68) | | | | | | | (1.04) |
| Age | | | | | | | | | -0.92* | -1.97* |
| | | | | | | | | | (0.41) | (0.84) |
| Edu-cation | | | | | 0.25* | | | 1.10* | 0.41** | |
| | | | | | (0.10) | | | (0.55) | (0.13) | |
| Talks to Healer | | | | | 1.30* | | | | | |
| | | | | | (0.53) | | | | | |
| Network size in village | | | | | -0.35* | | | | | |
| | | | | | (0.15) | | | | | |
| Network size out of village | | | | | | 0.93*** | | | | |
| | | | | | | (0.25) | | | | |
| House-hold exposure | | | 1.85** | 2.07*** | 1.92*** | | | | 2.95*** | 4.73*** |
| | | | (0.63) | (0.44) | (0.38) | | | | (0.47) | (1.08) |
| N | 98 | 46 | 72 | 317 | 293 | 113 | 41 | 117 | 194 | 207 |
| AIC | 78.71 | 11.64 | 69.60 | 173.28 | 220.79 | 104.52 | 32.41 | 24.65 | 157.25 | 45.36 |
| BIC | 81.30 | 13.46 | 76.43 | 180.80 | 239.19 | 112.70 | 34.12 | 30.18 | 173.59 | 58.69 |
| Pseudo R2 | 0.00 | 0.00 | 0.28 | 0.14 | 0.21 | 0.26 | 0.00 | 0.28 | 0.48 | 0.62 |

*** $p < 0.001$; ** $p < 0.01$; * $p < 0.05$.

**Table D7:** Results of village-level backward model selection procedures for logistic regressions explaining the use of coils. Parameters are reported with standard errors (in parentheses), along with the number of observations (N) and goodness of fit statistics at the bottom of the table.

| Village | WK1 | WK2 | WK3 | WJ1 | WJ2 | WJ3 | SG1 | SG2 | SG3 | SG4 |
|---|---|---|---|---|---|---|---|---|---|---|
| Intercept | -2.24** (0.81) | -0.94 (1.71) | -2.55** (0.88) | -0.33 (0.47) | -0.69 (0.42) | 1.10 (0.57) | 6.60 (3.66) | -3.12*** (0.67) | 4.42*** (1.00) | -2.46 (1.31) |
| Female | | -4.34* (1.92) | | -0.71* (0.32) | | | | | | |
| Head of house-hold | | | | | | | | | -3.06** (1.03) | |
| Carer for a sick person | 1.92* (0.76) | 3.15* (1.43) | | -0.90* (0.46) | | | | | | 3.89** (1.34) |
| Works in fields | -1.89** (0.73) | | -1.25* (0.63) | | | | -6.19* (2.93) | | | |
| Age | | -2.15* (0.90) | | | | | | | | -0.91* (0.46) |
| Edu-cation | | | | | 0.21* (0.10) | | -1.40* (0.65) | 0.58*** (0.15) | | |
| Talks to Asha | | | 2.24** (0.69) | | | 1.07* (0.53) | | | | |
| Network exposure | | | 4.43*** (1.28) | 1.88*** (0.38) | 0.90* (0.42) | 2.60*** (0.73) | | | | 4.36*** (1.08) |
| Network size in village | | | | | | -0.79** (0.25) | 3.37** (1.28) | 0.45* (0.17) | | -1.13*** (0.28) |
| Network size out of village | | | | | 0.31* (0.13) | | | | | |
| House-hold talks to Asha | | | | -0.63* (0.32) | | | -6.27** (2.40) | | | |
| House-hold talks to Healer | | | | | -1.11** (0.40) | | | | | |
| House-hold exposure | 3.43*** (0.62) | 6.59** (2.14) | | 1.72*** (0.31) | 1.12** (0.35) | | | | | 3.12*** (0.87) |
| N | 102 | 46 | 72 | 317 | 293 | 113 | 41 | 117 | 201 | 208 |
| AIC | 94.10 | 36.04 | 78.15 | 353.40 | 288.58 | 109.88 | 35.01 | 130.84 | 133.59 | 84.91 |
| BIC | 104.60 | 45.19 | 87.26 | 375.96 | 310.66 | 120.79 | 43.58 | 139.13 | 140.19 | 104.94 |
| Pseudo R2 | 0.52 | 0.72 | 0.43 | 0.35 | 0.18 | 0.27 | 0.68 | 0.26 | 0.19 | 0.68 |

*** $p < 0.001$; ** $p < 0.01$; * $p < 0.05$.

**Table D8:** Results of village-level backward model selection procedures for logistic regressions explaining the use of vaporisers. Parameters are reported with standard errors (in parentheses), along with the number of observations (N) and goodness of fit statistics at the bottom of the table.

| Village | WK1 | WK2 | WK3 | WJ1 | WJ2 | SG3 | SG4 |
|---|---|---|---|---|---|---|---|
| Intercept | -3.20 ***<br>(0.51) | -3.99 ***<br>(1.12) | -2.66 ***<br>(0.52) | -2.31 ***<br>(0.31) | -4.24 ***<br>(0.54) | -6.52 ***<br>(1.28) | -4.22 ***<br>(0.58) |
| Head of household | | | | -0.74 *<br>(0.36) | | 3.01 ***<br>(0.83) | |
| Education | | | | | | 0.50 **<br>(0.16) | |
| Network exposure | | 4.74 **<br>(1.45) | | 1.10 *<br>(0.47) | 3.16 *<br>(1.51) | | |
| Network size out of village | | | | | | -1.25 *<br>(0.51) | |
| Household talks to Healer | | | | | 1.49 *<br>(0.73) | | |
| Household exposure | | | 2.47 **<br>(0.80) | 2.34 ***<br>(0.37) | | 2.96 ***<br>(0.57) | |
| N | 102 | 46 | 72 | 317 | 293 | 201 | 208 |
| AIC | 35.75 | 22.96 | 48.69 | 257.55 | 72.24 | 116.56 | 33.39 |
| BIC | 38.38 | 26.62 | 53.24 | 272.58 | 83.28 | 133.08 | 36.73 |
| Pseudo R2 | 0.00 | 0.56 | 0.24 | 0.36 | 0.11 | 0.49 | 0.00 |

*** p < 0.001; ** p < 0.01; * p < 0.05.

**Table D9:** Results of village-level backward model selection procedures for logistic regressions explaining burning materials. Parameters are reported with standard errors (in parentheses), along with the number of observations (N) and goodness of fit statistics at the bottom of the table.

| Village | WK1 | WK2 | WK3 | WJ1 | WJ2 | WJ3 | SG1 | SG2 | SG3 |
|---|---|---|---|---|---|---|---|---|---|
| Intercept | -2.94*** (0.85) | -2.12*** (0.61) | -1.69*** (0.48) | -1.14*** (0.20) | -2.33*** (0.33) | -1.39*** (0.36) | -2.22*** (0.53) | -3.96*** (1.01) | -4.60*** (0.71) |
| Female | | | | | 0.68* (0.32) | | | | |
| Head of household | | | | | | | | -2.43*** (0.65) | |
| Works in fields | 1.61* (0.81) | | | | | | | | |
| Age | | | | | | | | 1.33** (0.47) | |
| Talks to Asha | | | | | | | | 1.46* (0.68) | |
| Talks to Healer | | | | | 1.01* (0.39) | | | | |
| Network exposure | 1.81** (0.65) | | 3.73** (1.21) | 0.96* (0.46) | | | | 3.14** (0.98) | |
| Network size out of village | | | | | | 0.45* (0.19) | | | |
| Household talks to Asha | | | | -0.65* (0.27) | | | | | |
| Household exposure | | 2.81*** (0.79) | | 1.19*** (0.33) | 1.83*** (0.30) | 1.03* (0.50) | | | |
| N | 102 | 46 | 72 | 317 | 293 | 113 | 41 | 116 | 201 |
| AIC | 120.39 | 45.98 | 87.83 | 357.08 | 294.23 | 144.70 | 28.21 | 96.44 | 24.42 |
| BIC | 128.26 | 49.64 | 92.39 | 372.12 | 308.95 | 152.89 | 29.93 | 110.21 | 27.72 |
| Pseudo R2 | 0.16 | 0.41 | 0.20 | 0.16 | 0.23 | 0.11 | 0.00 | 0.51 | 0.00 |

*** $p < 0.001$; ** $p < 0.01$; * $p < 0.05$.

#### D.2 Model selection II: Identical model specifications across villages

As the above model specifications differ between villages for every measure, these models cannot be used in a meta-analysis of regressions across villages. To enable this, we first had to identify a common model specification, one per dependent variable, for which all parameters could be estimated in every village. We do so by following a systematic trial-and-error strategy, as described in the Methods section of the main text (Procedure 1, Stage II). For each dependent variable, our procedure finds the largest set of explanatory variables identified in the backward selection stage for which the model estimation converges in every village and none of the estimates are affected by the Donner-Hauck phenomenon leading to a very large standard error of the estimate (Hauck & Donner, 1977). In a small number of cases, it was necessary to exclude an explanatory variable from one of the village models to achieve this result. However, this was restricted to a single variable and village. If this step did not fix the issue, the given village model was excluded from the meta-analysis in the following step. **Table**

**D10-D17** show the village-level logistic regression results, one for each preventive measure based on the identified common specifications across the villages.

**Table D10.** Results of village-level logistic regressions with the same specification in each village explaining the use of LLINs. Parameters are reported with standard errors (in parentheses), along with the number of observations (N) and goodness of fit statistics at the bottom of the table.

| Village | WK1 | WK2 | WK3 | WJ1 | WJ2 | WJ3 | SG1 | SG2 | SG3 | SG4 |
|---|---|---|---|---|---|---|---|---|---|---|
| Intercept | 0.19 | -2.21* | -0.44 | 2.30** | - | 1.38 | 1.23 | - | 2.98** | -0.53 |
| | (0.62) | (1.06) | (1.41) | (0.82) | - | (0.94) | (1.58) | - | (1.02) | (0.63) |
| Network | 0.95 | 3.82* | 2.41 . | 2.32* | - | 1.91 | 0.19 | - | 3.03*** | 2.63*** |
| exposure | (0.91) | (1.80) | (1.41) | (1.10) | - | (1.16) | (1.63) | - | (0.84) | (0.80) |
| Household | 0.64 | 2.38 | 0.60 | -0.03 | - | -0.99 | 0.95 | - | -2.11 | 1.60* |
| exposure | (0.61) | (1.51) | (0.90) | (1.07) | - | (0.83) | (1.00) | - | (1.19) | (0.70) |
| N | 102 | 48 | 73 | 317 | - | 114 | 44 | - | 205 | 212 |
| AIC | 105.24 | 24.98 | 53.40 | 52.94 | - | 65.84 | 36.29 | - | 61.81 | 78.74 |
| BIC | 113.12 | 30.60 | 60.28 | 64.22 | - | 74.05 | 41.64 | - | 71.78 | 88.80 |
| Pseudo R2 | 0.07 | 0.73 | 0.08 | 0.09 | - | 0.06 | 0.04 | - | 0.20 | 0.30 |

*** p < 0.001; ** p < 0.01; * p < 0.05.

**Table D11.** Results of village-level logistic regressions with the same specification in each village explaining the use of covering clothes. Parameters are reported with standard errors (in parentheses), along with the number of observations (N) and goodness of fit statistics at the bottom of the table.

| Village | WK1 | WK2 | WK3 | WJ1 | WJ2 | WJ3 | SG1 | SG2 | SG3 | SG4 |
|---|---|---|---|---|---|---|---|---|---|---|
| Intercept | 0.69 | 0.08 | -2.04* | -0.69 . | -0.04 | -0.04 | -5.94** | -2.06** | -0.51 | -2.98*** |
| | (0.69) | (1.21) | (1.04) | (0.40) | (0.34) | (0.78) | (2.15) | (0.80) | (0.49) | (0.71) |
| Head of | 0.13 | -0.73 | 0.23 | -0.02 | -0.04 | 0.12 | 0.24 | 0.14 | -1.26*** | -1.3 * |
| house-hold | (0.44) | (0.72) | (0.71) | (0.32) | (0.27) | (0.54) | (1.01) | (0.45) | (0.34) | (0.54) |
| Talks to | -0.17 | 0.59 | 0.14 | -1.00* | 0.04 | 2.15** | -1.64 | 0.42 | 1.13** | 1.59* |
| Asha | (0.48) | (1.14) | (0.76) | (0.41) | (0.31) | (0.69) | (1.39) | (0.64) | (0.42) | (0.68) |
| Network | -0.25 | 1.82 | 1.88 | 1.73*** | 0.47 | 2.34** | 2.78 | 0.34 | 0.77 | 0.86 |
| exposure | (0.81) | (1.11) | (1.36) | (0.48) | (0.42) | (0.84) | (1.98) | (0.91) | (0.57) | (0.85) |
| Network | -0.07 | -0.44 | -0.05 | 0.19 | -0.12 | -0.56 . | 1.29** | 0.46* | -0.07 | 0.70** |
| size in village | (0.15) | (0.44) | (0.20) | (0.16) | (0.10) | (0.31) | (0.48) | (0.21) | (0.22) | (0.24) |
| Network size out | -0.10 | 0.02 | 0.18 | 0.32* | 0.32** | 0.09 | 1.70* | 0.92** | 0.53* | 0.74*** |
| of village | (0.13) | (0.23) | (0.19) | (0.16) | (0.12) | (0.23) | (0.69) | (0.31) | (0.26) | (0.22) |
| House- | 0.87 | 0.72 | 1.98* | 1.15*** | 0.79* | 0.30 | -1.47 | -0.34 | 0.04 | 0.11 |
| hold exposure | (0.56) | (0.86) | (0.84) | (0.33) | (0.31) | (0.61) | (1.18) | (0.59) | (0.46) | (0.59) |
| N | 102 | 48 | 73 | 317 | 293 | 114 | 43 | 117 | 198 | 207 |
| AIC | 141.01 | 65.61 | 73.73 | 294.90 | 353.92 | 113.93 | 44.89 | 152.35 | 244.04 | 158.46 |
| BIC | 159.39 | 78.71 | 89.77 | 321.21 | 379.68 | 133.08 | 57.22 | 171.69 | 267.06 | 181.79 |
| Pseudo R2 | 0.07 | 0.35 | 0.43 | 0.25 | 0.10 | 0.19 | 0.65 | 0.25 | 0.19 | 0.66 |

*** p < 0.001; ** p < 0.01; * p < 0.05.

**Table D12.** Results of village-level logistic regressions with the same specification in each village explaining the use of boots. Parameters are reported with standard errors (in parentheses), along with the number of observations (N) and goodness of fit statistics at the bottom of the table.

| Village | WK1 | WK2 | WK3 | WJ1 | WJ2 | WJ3 | SG1 | SG2 | SG3 | SG4 |
|---|---|---|---|---|---|---|---|---|---|---|
| Intercept | -0.62 . | -2.01* | -0.73 | -0.64 * | -1.42*** | -0.47 | -3.08** | -3.10*** | -2.81*** | - |
| | (0.35) | (0.82) | (0.48) | (0.28) | (0.27) | (0.45) | (1.03) | (0.67) | (0.43) | - |
| Female | -2.16*** | -1.79 | -1.65** | -1.34*** | -0.98** | -1.69** | 0.08 | -0.78 | 0.31 | - |
| | (0.61) | (1.45) | (0.59) | (0.29) | (0.30) | (0.55) | (1.20) | (1.22) | (0.50) | - |
| Network | 0.23 | 1.88 | 0.45 | 1.62*** | 1.44** | -0.08 | 2.52 | -6.13 | 1.79 | - |
| exposure | (1.31) | (2.36) | (1.49) | (0.37) | (0.46) | (1.09) | (3.44) | (5.63) | (0.93) | - |
| House- | 1.80 | 2.44. | 0.89 | 0.90*** | 0.80* | 1.02 | 1.48 | 3.00* | 1.73** | - |
| hold exposure | (0.92) | (1.41) | (0.66) | (0.27) | (0.34) | (0.79) | (2.25) | (1.30) | (0.63) | - |
| N | 102 | 28 | 73 | 317 | 293 | 96 | 43 | 102 | 201 | - |
| AIC | 103.68 | 28.76 | 85.02 | 375.18 | 292.03 | 92.50 | 28.82 | 41.78 | 125.74 | - |
| BIC | 114.18 | 34.09 | 94.18 | 390.22 | 306.75 | 102.76 | 35.87 | 52.28 | 138.96 | - |
| Pseudo R2 | 0.29 | 0.40 | 0.19 | 0.27 | 0.17 | 0.21 | 0.27 | 0.18 | 0.23 | - |

*** p < 0.001; ** p < 0.01; * p < 0.05.

**Table D13.** Results of village-level logistic regressions with the same specification in each village explaining the use of gloves. Parameters are reported with standard errors (in parentheses), along with the number of observations (N) and goodness of fit statistics at the bottom of the table.

| Village | WK1 | WK2 | WK3 | WJ1 | WJ2 | WJ3 | SG1 | SG2 | SG3 | SG4 |
|---|---|---|---|---|---|---|---|---|---|---|
| Intercept | -2.00* | -3.78* | -0.83 | -2.65** | -4.06*** | - | - | - | -9.14* | - |
| | (0.80) | (1.83) | (1.07) | (0.87) | (0.80) | - | - | - | (3.56) | - |
| Education | 0.78** | 0.14 | 0.15 | -0.25 | -0.14 | - | - | - | 1.42* | - |
| | (0.26) | (0.29) | (0.20) | (0.31) | (0.21) | - | - | - | (0.70) | - |
| Network | -0.53* | 0.57 | -0.50* | -0.44 | 0.30 | - | - | - | -1.89 | - |
| size in village | (0.27) | (0.43) | (0.24) | (0.33) | (0.20) | - | - | - | (1.03) | - |
| Household | 0.27 | 2.79* | 1.63* | 2.79** | 2.41*** | - | - | - | 6.02** | - |
| exposure | (0.60) | (1.09) | (0.73) | (0.94) | (0.70) | - | - | - | (2.15) | - |
| N | 102 | 27 | 72 | 317 | 293 | - | - | - | 194 | - |
| AIC | 95.52 | 33.02 | 62.90 | 74.92 | 89.39 | - | - | - | 32.42 | - |
| BIC | 106.02 | 38.21 | 72.01 | 89.96 | 104.11 | - | - | - | 45.49 | - |
| Pseudo R2 | 0.23 | 0.44 | 0.22 | 0.12 | 0.15 | - | - | - | 0.50 | - |

*** p < 0.001; ** p < 0.01; * p < 0.05.

**Table D14.** Results of village-level logistic regressions with the same specification in each village explaining the use of insecticide cream. Parameters are reported with standard errors (in parentheses), along with the number of observations (N) and goodness of fit statistics at the bottom of the table.

| Village | WK1 | WK2 | WK3 | WJ1 | WJ2 | WJ3 | SG1 | SG2 | SG3 | SG4 |
|---|---|---|---|---|---|---|---|---|---|---|
| Intercept | -5.89** | - | -1.37 | -4.01*** | -3.88*** | -0.85 | - | - | -3.84*** | -0.90 |
| | (2.26) | - | (1.66) | (1.02) | (0.91) | (0.94) | - | - | (0.95) | (1.80) |
| Works in fields | 1.33 | - | -0.85 | 0.54 | 0.16 | -0.50 | - | - | 0.95* | -2.21* |
| | (1.23) | - | (0.80) | (0.59) | (0.40) | (0.50) | - | - | (0.43) | (1.08) |
| Age | 0.50 | - | -0.47 | 0.26 | 0.34 | 0.02 | - | - | -0.42 | -1.87* |
| | (0.52) | - | (0.57) | (0.31) | (0.29) | (0.37) | - | - | (0.33) | (0.87) |
| Education | 0.69* | - | 0.13 | 0.13 | 0.33* | -0.14 | - | - | 0.48*** | 0.11 |
| | (0.33) | - | (0.22) | (0.18) | (0.13) | (0.19) | - | - | (0.13) | (0.27) |
| Household exposure | 1.35 . | - | 1.77** | 2.04*** | 1.73*** | 0.82 | - | - | 2.71*** | 4.65*** |
| | (0.74) | - | (0.64) | (0.44) | (0.36) | (0.58) | - | - | (0.43) | (1.08) |
| N | 98 | - | 72 | 317 | 293 | 113 | - | - | 201 | 209 |
| AIC | 78.89 | - | 71.96 | 177.54 | 227.27 | 126.28 | - | - | 173.11 | 47.18 |
| BIC | 91.81 | - | 83.35 | 196.33 | 245.68 | 139.92 | - | - | 189.63 | 63.89 |
| Pseudo R2 | 0.14 | - | 0.31 | 0.15 | 0.17 | 0.04 | - | - | 0.42 | 0.62 |

*** p < 0.001; ** p < 0.01; * p < 0.05.

**Table D15.** Results of village-level logistic regressions with the same specification in each village explaining the use of coils. Parameters are reported with standard errors (in parentheses), along with the number of observations (N) and goodness of fit statistics at the bottom of the table.

| Village | WK1 | WK2 | WK3 | WJ1 | WJ2 | WJ3 | SG1 | SG2 | SG3 | SG4 |
|---|---|---|---|---|---|---|---|---|---|---|
| Intercept | -2.02 | -4.11 | -1.68 | -0.35 | -0.33 | 1.51 | 0.93 | -3.71** | - | -3.44* |
| | (1.24) | (2.12) | (1.39) | (0.60) | (0.70) | (1.10) | (2.74) | (1.38) | - | (1.55) |
| Carer for a sick person | 1.76* | 1.96 | -0.14 | -1.13** | 0.04 | 0.51 | 0.57 | 1.96 | - | 3.31** |
| | (0.77) | (1.16) | (0.69) | (0.44) | (0.52) | (0.82) | (1.44) | (1.25) | - | (1.26) |
| Works in fields | -2.03** | 1.44 | -0.96 | -0.17 | 0.02 | -1.06 | -1.91 | -1.17 | - | -0.50 |
| | (0.74) | (1.52) | (0.66) | (0.33) | (0.33) | (0.63) | (1.44) | (0.81) | - | (0.78) |
| Edu-cation | 0.18 | -0.16 | -0.03 | -0.11 | 0.19 | 0.14 | -0.37 | 0.58*** | - | 0.11 |
| | (0.25) | (0.39) | (0.20) | (0.11) | (0.11) | (0.21) | (0.34) | (0.18) | - | (0.21) |
| Network exposure | 1.48 | 3.95* | 2.60* | 1.80*** | 0.97* | 2.01* | 1.06 | -0.39 | - | 4.78*** |
| | (1.15) | (1.97) | (1.13) | (0.37) | (0.44) | (0.79) | (1.50) | (0.88) | - | (1.07) |
| Network size in village | -0.21 | -0.42 | 0.14 | -0.01 | -0.11 | -0.64* | 1.13* | 0.43* | - | -1.22*** |
| | (0.19) | (0.43) | (0.17) | (0.11) | (0.11) | (0.25) | (0.46) | (0.20) | - | (0.29) |
| House-hold exposure | 2.77*** | 1.09 | 1.43* | 1.49*** | 0.96** | 0.35 | -1.90 | -0.02 | - | 2.96*** |
| | (0.79) | (1.31) | (0.64) | (0.28) | (0.33) | (0.59) | (1.11) | (0.50) | - | (0.82) |
| N | 102 | 46 | 72 | 317 | 293 | 113 | 41 | 117 | - | 210 |
| AIC | 95.74 | 48.12 | 89.17 | 361.55 | 301.30 | 116.12 | 49.86 | 134.53 | - | 90.27 |
| BIC | 114.12 | 60.92 | 105.11 | 387.86 | 327.06 | 135.21 | 61.86 | 153.86 | - | 113.70 |
| Pseudo R2 | 0.56 | 0.60 | 0.36 | 0.33 | 0.13 | 0.27 | 0.46 | 0.30 | - | 0.67 |

*** p < 0.001; ** p < 0.01; * p < 0.05.

**Table D16.** Results of village-level logistic regressions with the same specification in each village explaining the use of vaporisers. Parameters are reported with standard errors (in parentheses), along with the number of observations (N) and goodness of fit statistics at the bottom of the table.

| Village | WK1 | WK2 | WK3 | WJ1 | WJ2 | WJ3 | SG1 | SG2 | SG3 | SG4 |
|---|---|---|---|---|---|---|---|---|---|---|
| Intercept | - | -3.77** | -2.69*** | -2.31*** | - | - | - | - | -4.31*** | - |
| | - | (1.28) | (0.59) | (0.31) | - | - | - | - | (0.70) | - |
| Head of household | - | -0.12 | -1.29 | -0.74* | - | - | - | - | 2.31*** | - |
| | - | (1.43) | (1.07) | (0.36) | - | - | - | - | (0.69) | - |
| Network exposure | - | 3.51 | 4.22 | 1.10* | - | - | - | - | -0.83 | - |
| | - | (2.01) | (2.45) | (0.47) | - | - | - | - | (0.90) | - |
| Household | - | 1.69 | 1.45 | 2.34*** | - | - | - | - | 2.98*** | - |
| exposure | - | (1.92) | (1.17) | (0.37) | - | - | - | - | (0.73) | - |
| N | - | 46 | 73 | 317 | - | - | - | - | 208 | - |
| AIC | - | 25.91 | 47.63 | 257.55 | - | - | - | - | 134.01 | - |
| BIC | - | 33.23 | 56.79 | 272.58 | - | - | - | - | 147.36 | - |
| Pseudo R2 | - | 0.59 | 0.35 | 0.36 | - | - | - | - | 0.37 | - |

*** p < 0.001; ** p < 0.01; * p < 0.05.

**Table D17.** Results of village-level logistic regressions with the same specification in each village explaining burning materials. Parameters are reported with standard errors (in parentheses), along with the number of observations (N) and goodness of fit statistics at the bottom of the table.

| Village | WK1 | WK2 | WK3 | WJ1 | WJ2 | WJ3 | SG1 | SG2 | SG3 | SG4 |
|---|---|---|---|---|---|---|---|---|---|---|
| Intercept | -1.47*** | -2.07*** | -1.77*** | -1.41*** | -1.65*** | -0.89*** | -2.45*** | -1.97*** | - | - |
| | (0.36) | (0.62) | (0.53) | (0.17) | (0.22) | (0.25) | (0.61) | (0.35) | - | - |
| Network | 1.71* | -0.67 | 3.73** | 1.00* | -0.25 | 0.87 | -0.74 | 2.70* | - | - |
| exposure | (0.78) | (1.42) | (1.25) | (0.46) | (0.49) | (0.68) | (2.75) | (1.21) | - | - |
| Household | -0.06 | 3.15** | 0.12 | 1.02** | 1.89*** | 0.38 | 2.38 | 0.80 | - | - |
| exposure | (0.56) | (1.09) | (0.55) | (0.32) | (0.37) | (0.60) | (1.56) | (0.66) | - | - |
| N | 102 | 46 | 73 | 317 | 293 | 114 | 44 | 116 | - | - |
| AIC | 125.68 | 47.76 | 90.12 | 360.95 | 302.06 | 150.13 | 33.81 | 118.45 | - | - |
| BIC | 133.55 | 53.24 | 96.99 | 372.22 | 313.10 | 158.34 | 39.16 | 126.71 | - | - |
| Pseudo R2 | 0.10 | 0.42 | 0.21 | 0.14 | 0.18 | 0.06 | 0.14 | 0.27 | - | - |

*** p < 0.001; ** p < 0.01; * p < 0.05.

#### D.3 Meta-analyses of village-level logistic regression models

In the main text of the paper, we present meta-analyses of the village-level logistic regression models presented in section D.2. A meta-analysis of estimates is performed for each parameter reported in **Tables D10-D17**. **Table D18** provides an overview of the village-level models that were excluded from the meta-analysis due to the issues discussed in the previous section. We used the metafor package in R (Viechtbauer, 2010) to perform the meta-analysis of the village-level models reported in the main text. Full results of the meta-analyses are reported in **Tables D19-26**. In each table, we report the estimated mean of the village-level parameters (μ), its standard error (se), the p-value of the test μ=0 (p: μ=0), the estimated between-village standard deviation of parameters (τ), Cohran's Q that reflects the variability of village-level parameters around the fixed-effects estimate of their mean (Q), the p-value of the test Q=0 (p: Q=0), and the number of village-level used in each parameter-wise meta-analysis (n).

Results of the meta-analysis show that the use of **LLINS** is positively associated with network exposure. Household exposure was the only other variable included in the model but it is not significant across villages, and therefore does not constitute a generalizable factor. The use of **Covering clothes** significantly increases with higher network and household exposure, but also with a higher degree outside village. There is significant variability in talking to the Asha and in the overall number of people villagers talk to within their own village, but both variables do not overall increase the use of clothes. Also use of **Boots** significantly increases with network and household exposure, while women tend to use them less. **Gloves** and **Vaporizers** are only used more when someone in the household also uses them, like **Insecticide creams**, although these are also used more by people with a higher level of education. Like in the case of covering clothes and boots, **Burning materials** and the use of **Coils** increase with network and household exposure, but coils are overall less common between people working in fields.

**Table D18**. Overview of village-level models included in the meta-analysis of logistic regressions explaining different preventive measures.

<table>
<tr><td></td><td>WK1</td><td>WK2</td><td>WK3</td><td>WJ1</td><td>WJ2</td><td>WJ3</td><td>SG1</td><td>SG2</td><td>SG3</td><td>SG4</td></tr>
<tr><td>LLINS</td><td></td><td></td><td></td><td></td><td style="background:#d0d0d0"></td><td></td><td></td><td style="background:#d0d0d0"></td><td></td><td></td></tr>
<tr><td>Covering clothes</td><td></td><td></td><td></td><td></td><td></td><td></td><td></td><td></td><td></td><td></td></tr>
<tr><td>Boots</td><td></td><td></td><td></td><td></td><td></td><td></td><td></td><td></td><td></td><td style="background:#d0d0d0"></td></tr>
<tr><td>Gloves</td><td></td><td></td><td></td><td></td><td></td><td style="background:#d0d0d0"></td><td style="background:#d0d0d0"></td><td style="background:#d0d0d0"></td><td></td><td style="background:#d0d0d0"></td></tr>
<tr><td>Insecticide Cream</td><td></td><td style="background:#d0d0d0"></td><td></td><td></td><td></td><td></td><td style="background:#d0d0d0"></td><td style="background:#d0d0d0"></td><td></td><td></td></tr>
<tr><td>Coils</td><td></td><td></td><td></td><td></td><td></td><td></td><td></td><td></td><td style="background:#d0d0d0"></td><td></td></tr>
<tr><td>Vaporizers</td><td style="background:#d0d0d0"></td><td></td><td></td><td></td><td style="background:#d0d0d0"></td><td style="background:#d0d0d0"></td><td style="background:#d0d0d0"></td><td style="background:#d0d0d0"></td><td></td><td style="background:#d0d0d0"></td></tr>
<tr><td>Burning materials</td><td></td><td></td><td></td><td></td><td></td><td></td><td></td><td></td><td style="background:#d0d0d0"></td><td style="background:#d0d0d0"></td></tr>
</table>

<table>
<tr><td></td><td>Village-level models used in meta-analysis</td></tr>
<tr><td style="background:#d0d0d0"></td><td>Village-level models not used in meta-analysis</td></tr>
</table>

**Table D19.** Full results of the meta-analysis of logistic regression models explaining the use of LLINs. See the current text for a description of the reported statistics.

| | μ | se | p: μ=0 | τ | Q | p: Q=0 | n |
|---|---|---|---|---|---|---|---|

| | | | | | | | |
|---|---|---|---|---|---|---|---|
| Intercept | 0.61 | 0.60 | 0.31 | 1.37 | 21.67 | 0.00 | 8 |
| Network exposure | 2.21 | 0.38 | 0.00 | 0.00 | 5.59 | 0.59 | 8 |
| Household exposure | 0.39 | 0.42 | 0.36 | 0.76 | 12.52 | 0.08 | 8 |

**Table D20.** Full results of the meta-analysis of logistic regression models explaining the use of covering clothes. See the current text for a description of the reported statistics.

| | μ | se | p: μ=0 | τ | Q | p: Q=0 | n |
|---|---|---|---|---|---|---|---|
| Intercept | -0.96 | 0.42 | 0.02 | 1.07 | 29.74 | 0.00 | 10 |
| Head of household | -0.28 | 0.21 | 0.17 | 0.43 | 16.97 | 0.05 | 10 |
| Talks to Asha | 0.38 | 0.33 | 0.25 | 0.82 | 28.12 | 0.00 | 10 |
| Network exposure | 1.04 | 0.28 | 0.00 | 0.43 | 11.32 | 0.25 | 10 |
| Network size in village | 0.10 | 0.13 | 0.44 | 0.32 | 27.94 | 0.00 | 10 |
| Network size out of village | 0.32 | 0.10 | 0.00 | 0.25 | 24.56 | 0.00 | 10 |
| Household exposure | 0.56 | 0.21 | 0.01 | 0.34 | 14.15 | 0.12 | 10 |

**Table D21.** Full results of the meta-analysis of logistic regression models explaining the use of boots. See the current text for a description of the reported statistics.

| | μ | se | p: μ=0 | τ | Q | p: Q=0 | n |
|---|---|---|---|---|---|---|---|
| Intercept | -1.51 | 0.35 | 0.00 | 0.92 | 37.45 | 0.00 | 9 |
| Female | -1.15 | 0.28 | 0.00 | 0.57 | 14.96 | 0.06 | 9 |
| Network exposure | 1.39 | 0.26 | 0.00 | 0.00 | 5.51 | 0.70 | 9 |
| Household exposure | 1.04 | 0.18 | 0.00 | 0.00 | 6.04 | 0.64 | 9 |

**Table D22.** Full results of the meta-analysis of logistic regression models explaining the use of gloves. See the current text for a description of the reported statistics.

| | μ | se | p: μ=0 | τ | Q | p: Q=0 | n |
|---|---|---|---|---|---|---|---|
| Intercept | -2.78 | 0.61 | 0.00 | 0.94 | 10.27 | 0.07 | 6 |
| Education | 0.21 | 0.18 | 0.25 | 0.33 | 12.87 | 0.02 | 6 |
| Network size in village | -0.22 | 0.22 | 0.32 | 0.42 | 15.91 | 0.01 | 6 |
| Household exposure | 2.08 | 0.56 | 0.00 | 1.01 | 13.17 | 0.02 | 6 |

**Table D23.** Full results of the meta-analysis of logistic regression models explaining the use of insecticide cream. See the current text for a description of the reported statistics.

| | μ | se | p: μ=0 | τ | Q | p: Q=0 | n |
|---|---|---|---|---|---|---|---|
| Intercept | -2.93 | 0.64 | 0.00 | 1.14 | 11.85 | 0.07 | 7 |
| Works in fields | 0.04 | 0.34 | 0.90 | 0.61 | 12.86 | 0.05 | 7 |
| Age | 0.00 | 0.17 | 0.98 | 0.20 | 9.91 | 0.13 | 7 |
| Education | 0.24 | 0.09 | 0.01 | 0.16 | 10.29 | 0.11 | 7 |
| Household exposure | 1.98 | 0.31 | 0.00 | 0.59 | 14.40 | 0.03 | 7 |

**Table D24.** Full results of the meta-analysis of logistic regression models explaining the use of coils. See the current text for a description of the reported statistics.

| | μ | se | p: μ=0 | τ | Q | p: Q=0 | n |
|---|---|---|---|---|---|---|---|
| Intercept | -1.25 | 0.62 | 0.04 | 1.32 | 16.96 | 0.03 | 9 |
| Carer for a sick person | 0.71 | 0.47 | 0.13 | 1.08 | 24.29 | 0.00 | 9 |
| Works in fields | -0.61 | 0.26 | 0.02 | 0.45 | 12.22 | 0.14 | 9 |
| Education | 0.10 | 0.09 | 0.25 | 0.17 | 15.34 | 0.05 | 9 |
| Network exposure | 1.81 | 0.48 | 0.00 | 1.06 | 18.91 | 0.02 | 9 |
| Network size in village | -0.12 | 0.19 | 0.53 | 0.51 | 37.37 | 0.00 | 9 |
| Household exposure | 1.07 | 0.41 | 0.01 | 1.00 | 25.82 | 0.00 | 9 |

**Table D25.** Full results of the meta-analysis of logistic regression models explaining the use of vaporisers. See the current text for a description of the reported statistics.

| | μ | se | p: μ=0 | τ | Q | p: Q=0 | n |
|---|---|---|---|---|---|---|---|
| Intercept | -3.06 | 0.51 | 0.00 | 0.77 | 7.63 | 0.05 | 4 |
| Head of household | 0.10 | 0.87 | 0.91 | 1.49 | 16.56 | 0.00 | 4 |
| Network exposure | 1.25 | 1.01 | 0.22 | 1.55 | 7.48 | 0.06 | 4 |
| Household exposure | 2.38 | 0.31 | 0.00 | 0.00 | 1.44 | 0.70 | 4 |

**Table D26.** Full results of the meta-analysis of logistic regression models explaining burning materials. See the current text for a description of the reported statistics.

| | μ | se | p: μ=0 | τ | Q | p: Q=0 | n |
|---|---|---|---|---|---|---|---|
| Intercept | -1.56 | 0.16 | 0.00 | 0.28 | 12.06 | 0.10 | 8 |
| Network exposure | 1.06 | 0.46 | 0.02 | 0.90 | 15.59 | 0.03 | 8 |
| Household exposure | 0.98 | 0.33 | 0.00 | 0.68 | 17.77 | 0.01 | 8 |

#### D.4 Assessment of the goodness of fit of village-level logistic regression models

Finally, we compare the accuracy of the village-level models to assess the relative importance of various factors in explaining the use of the different measures. **Table D27** reports the average accuracy (correct classification rate) of village-level models by preventive measure. The figures reported in the main text can be found in the "Total" column. **Table D28** provides results of paired-sample t-tests of the difference of average accuracy statistics between different model specifications presented in **Table D27**. **Tables D29-38** present the accuracy of models in each village. In each table, the assessed model specifications include the same set of variables as described in the main text:

A. intercept,
B. individual characteristics,
C. opinion leaders (Asha, Healer),
D. network size,
E. network exposure,
F. household exposure.

**Table D27.** Average accuracy of different village-level logistic model specifications explaining the use of each preventive measure.

| | LLINs | Covering clothes | Boots | Gloves | Insecticide cream | Coils | Vaporisers | Burning materials | Total |
|---|---|---|---|---|---|---|---|---|---|
| Empty model (A) | 0.90 | 0.63 | 0.78 | 0.86 | 0.84 | 0.68 | 0.84 | 0.71 | 0.78 |
| Individual model (A+B) | 0.90 | 0.65 | 0.80 | 0.87 | 0.84 | 0.69 | 0.84 | 0.71 | 0.79 |
| Opinion leader model (A+B+C) | 0.90 | 0.69 | 0.80 | 0.87 | 0.84 | 0.69 | 0.84 | 0.71 | 0.79 |
| Network size model (A+B+C+D) | 0.90 | 0.74 | 0.80 | 0.87 | 0.84 | 0.71 | 0.84 | 0.71 | 0.80 |
| Network exposure model (A+B+C+D+E) | 0.91 | 0.76 | 0.81 | 0.87 | 0.84 | 0.77 | 0.86 | 0.73 | 0.82 |
| Household exposure model (A+B+C+D+F) | 0.91 | 0.75 | 0.81 | 0.88 | 0.87 | 0.79 | 0.88 | 0.73 | 0.83 |
| Full model (A+B+C+D+E+F) | 0.92 | 0.76 | 0.82 | 0.88 | 0.87 | 0.81 | 0.88 | 0.75 | 0.84 |

**Table D28.** Paired-sample t-tests comparing the average accuracy of logistic model specifications. Model names are identical to those in Table D27; difference refers to Mean 1 – Mean 2; the t-statistics are the results of paired-sample t-tests with H0: Mean 1 – Mean 2 = 0; p refers to two-sided p-values of the t-test; df are the degrees of freedom (number of fitted models as in Table D18 – 1).

| Model 1 | Model 2 | Mean 1 | Mean 2 | Difference | t-statistic | p | df |
|---|---|---|---|---|---|---|---|
| Individual | Empty | 0.77 | 0.77 | 0.01 | 2.35 | 0.02 | 60 |
| Opinion leader | Individual | 0.78 | 0.77 | 0.01 | 1.71 | 0.09 | 60 |
| Network size | Opinion leader | 0.79 | 0.78 | 0.01 | 2.72 | 0.01 | 60 |
| Network exposure | Network size | 0.81 | 0.79 | 0.02 | 3.82 | 0.00 | 60 |
| Household exposure | Network size | 0.82 | 0.79 | 0.03 | 4.42 | 0.00 | 60 |
| Full | Network size | 0.83 | 0.79 | 0.04 | 5.16 | 0.00 | 60 |
| Household exposure | Network exposure | 0.82 | 0.81 | 0.01 | 1.75 | 0.09 | 60 |
| Full | Household exposure | 0.83 | 0.82 | 0.01 | 3.46 | 0.00 | 60 |
| Full | Network exposure | 0.83 | 0.81 | 0.02 | 4.00 | 0.00 | 60 |

**Table D29.** Accuracy of different village-level logistic model specifications explaining the use of each preventive measure in village WK1.

| | LLINs | Covering clothes | Boots | Gloves | Insecticide cream | Coils | Vaporisers | Burning materials |
|---|---|---|---|---|---|---|---|---|
| Empty model (A) | 0.79 | 0.65 | 0.74 | 0.79 | 0.87 | 0.62 | - | 0.69 |
| Individual model (A+B) | 0.79 | 0.65 | 0.74 | 0.80 | 0.87 | 0.61 | - | 0.69 |
| Opinion leader model (A+B+C) | 0.79 | 0.65 | 0.74 | 0.80 | 0.87 | 0.61 | - | 0.69 |
| Network size model (A+B+C+D) | 0.79 | 0.67 | 0.74 | 0.80 | 0.87 | 0.63 | - | 0.69 |
| Network exposure model (A+B+C+D+E) | 0.79 | 0.66 | 0.78 | 0.80 | 0.87 | 0.75 | - | 0.69 |
| Household exposure model (A+B+C+D+F) | 0.79 | 0.69 | 0.79 | 0.79 | 0.86 | 0.84 | - | 0.69 |
| Full model (A+B+C+D+E+F) | 0.79 | 0.69 | 0.79 | 0.79 | 0.86 | 0.85 | - | 0.69 |

**Table D30.** Accuracy of different village-level logistic model specifications explaining the use of each preventive measure in village WK2.

| | LLINs | Covering clothes | Boots | Gloves | Insecticide cream | Coils | Vaporisers | Burning materials |
|---|---|---|---|---|---|---|---|---|
| Empty model (A) | 0.79 | 0.54 | 0.79 | 0.64 | - | 0.64 | 0.87 | 0.66 |
| Individual model (A+B) | 0.79 | 0.54 | 0.79 | 0.64 | - | 0.68 | 0.87 | 0.66 |
| Opinion leader model (A+B+C) | 0.79 | 0.54 | 0.79 | 0.64 | - | 0.68 | 0.87 | 0.66 |
| Network size model (A+B+C+D) | 0.79 | 0.62 | 0.79 | 0.68 | - | 0.68 | 0.87 | 0.66 |
| Network exposure model (A+B+C+D+E) | 0.94 | 0.73 | 0.79 | 0.68 | - | 0.85 | 0.94 | 0.66 |
| Household exposure model (A+B+C+D+F) | 0.90 | 0.67 | 0.82 | 0.74 | - | 0.83 | 0.93 | 0.80 |
| Full model (A+B+C+D+E+F) | 0.96 | 0.73 | 0.86 | 0.74 | - | 0.85 | 0.93 | 0.80 |

**Table D31.** Accuracy of different village-level logistic model specifications explaining the use of each preventive measure in village WK3.

| | LLINs | Covering clothes | Boots | Gloves | Insecticide cream | Coils | Vaporisers | Burning materials |
|---|---|---|---|---|---|---|---|---|
| Empty model (A) | 0.89 | 0.73 | 0.71 | 0.84 | 0.77 | 0.58 | 0.88 | 0.63 |
| Individual model (A+B) | 0.89 | 0.73 | 0.71 | 0.83 | 0.76 | 0.57 | 0.88 | 0.63 |
| Opinion leader model (A+B+C) | 0.89 | 0.73 | 0.71 | 0.83 | 0.76 | 0.57 | 0.88 | 0.63 |
| Network size model (A+B+C+D) | 0.89 | 0.75 | 0.71 | 0.82 | 0.76 | 0.58 | 0.88 | 0.63 |
| Network exposure model (A+B+C+D+E) | 0.89 | 0.82 | 0.73 | 0.82 | 0.76 | 0.67 | 0.86 | 0.71 |
| Household exposure model (A+B+C+D+F) | 0.89 | 0.85 | 0.73 | 0.82 | 0.82 | 0.74 | 0.88 | 0.63 |
| Full model (A+B+C+D+E+F) | 0.89 | 0.85 | 0.73 | 0.82 | 0.82 | 0.76 | 0.86 | 0.71 |

**Table D32.** Accuracy of different village-level logistic model specifications explaining the use of each preventive measure in village WJ1.

| | LLINs | Covering clothes | Boots | Gloves | Insecticide cream | Coils | Vaporisers | Burning materials |
|---|---|---|---|---|---|---|---|---|
| Empty model (A) | 0.98 | 0.78 | 0.51 | 0.97 | 0.91 | 0.52 | 0.78 | 0.70 |
| Individual model (A+B) | 0.98 | 0.78 | 0.62 | 0.97 | 0.91 | 0.54 | 0.78 | 0.70 |
| Opinion leader model (A+B+C) | 0.98 | 0.78 | 0.62 | 0.97 | 0.91 | 0.54 | 0.78 | 0.70 |
| Network size model (A+B+C+D) | 0.98 | 0.78 | 0.62 | 0.97 | 0.91 | 0.56 | 0.78 | 0.70 |
| Network exposure model (A+B+C+D+E) | 0.98 | 0.78 | 0.67 | 0.97 | 0.91 | 0.69 | 0.78 | 0.71 |
| Household exposure model (A+B+C+D+F) | 0.98 | 0.76 | 0.67 | 0.97 | 0.91 | 0.72 | 0.84 | 0.71 |
| Full model (A+B+C+D+E+F) | 0.98 | 0.79 | 0.70 | 0.97 | 0.91 | 0.72 | 0.84 | 0.73 |

**Table D33.** Accuracy of different village-level logistic model specifications explaining the use of each preventive measure in village WJ2.

| | LLINs | Covering clothes | Boots | Gloves | Insecticide cream | Coils | Vaporisers | Burning materials |
|---|---|---|---|---|---|---|---|---|
| Empty model (A) | - | 0.69 | 0.76 | 0.96 | 0.85 | 0.77 | - | 0.74 |
| Individual model (A+B) | - | 0.69 | 0.76 | 0.96 | 0.85 | 0.77 | - | 0.74 |
| Opinion leader model (A+B+C) | - | 0.69 | 0.76 | 0.96 | 0.85 | 0.77 | - | 0.74 |
| Network size model (A+B+C+D) | - | 0.69 | 0.76 | 0.96 | 0.85 | 0.77 | - | 0.74 |
| Network exposure model (A+B+C+D+E) | - | 0.71 | 0.78 | 0.96 | 0.85 | 0.77 | - | 0.74 |
| Household exposure model (A+B+C+D+F) | - | 0.71 | 0.77 | 0.96 | 0.86 | 0.77 | - | 0.75 |
| Full model (A+B+C+D+E+F) | - | 0.71 | 0.78 | 0.96 | 0.86 | 0.78 | - | 0.75 |

**Table D34.** Accuracy of different village-level logistic model specifications explaining the use of each preventive measure in village WJ3.

| | LLINs | Covering clothes | Boots | Gloves | Insecticide cream | Coils | Vaporisers | Burning materials |
|---|---|---|---|---|---|---|---|---|
| Empty model (A) | 0.92 | 0.80 | 0.79 | - | 0.78 | 0.75 | - | 0.64 |
| Individual model (A+B) | 0.92 | 0.80 | 0.79 | - | 0.78 | 0.76 | - | 0.64 |
| Opinion leader model (A+B+C) | 0.92 | 0.80 | 0.79 | - | 0.78 | 0.76 | - | 0.64 |
| Network size model (A+B+C+D) | 0.92 | 0.80 | 0.79 | - | 0.78 | 0.76 | - | 0.64 |
| Network exposure model (A+B+C+D+E) | 0.92 | 0.81 | 0.78 | - | 0.78 | 0.81 | - | 0.65 |
| Household exposure model (A+B+C+D+F) | 0.92 | 0.80 | 0.80 | - | 0.78 | 0.81 | - | 0.65 |
| Full model (A+B+C+D+E+F) | 0.92 | 0.82 | 0.80 | - | 0.78 | 0.80 | - | 0.68 |

**Table D35.** Accuracy of different village-level logistic model specifications explaining the use of each preventive measure in village SG1.

| | LLINs | Covering clothes | Boots | Gloves | Insecticide cream | Coils | Vaporisers | Burning materials |
|---|---|---|---|---|---|---|---|---|
| Empty model (A) | 0.89 | 0.51 | 0.91 | - | - | 0.68 | - | 0.89 |
| Individual model (A+B) | 0.89 | 0.51 | 0.91 | - | - | 0.68 | - | 0.89 |
| Opinion leader model (A+B+C) | 0.89 | 0.65 | 0.91 | - | - | 0.68 | - | 0.89 |
| Network size model (A+B+C+D) | 0.89 | 0.84 | 0.91 | - | - | 0.73 | - | 0.89 |
| Network exposure model (A+B+C+D+E) | 0.89 | 0.84 | 0.88 | - | - | 0.73 | - | 0.89 |
| Household exposure model (A+B+C+D+F) | 0.89 | 0.84 | 0.91 | - | - | 0.78 | - | 0.89 |
| Full model (A+B+C+D+E+F) | 0.89 | 0.84 | 0.88 | - | - | 0.78 | - | 0.89 |

**Table D36.** Accuracy of different village-level logistic model specifications explaining the use of each preventive measure in village SG2.

| | LLINs | Covering clothes | Boots | Gloves | Insecticide cream | Coils | Vaporisers | Burning materials |
|---|---|---|---|---|---|---|---|---|
| Empty model (A) | - | 0.5 | 0.95 | - | - | 0.67 | - | 0.72 |
| Individual model (A+B) | - | 0.53 | 0.95 | - | - | 0.75 | - | 0.72 |
| Opinion leader model (A+B+C) | - | 0.61 | 0.95 | - | - | 0.75 | - | 0.72 |
| Network size model (A+B+C+D) | - | 0.66 | 0.95 | - | - | 0.78 | - | 0.72 |
| Network exposure model (A+B+C+D+E) | - | 0.66 | 0.95 | - | - | 0.79 | - | 0.78 |
| Household exposure model (A+B+C+D+F) | - | 0.64 | 0.95 | - | - | 0.78 | - | 0.73 |
| Full model (A+B+C+D+E+F) | - | 0.65 | 0.95 | - | - | 0.79 | - | 0.78 |

**Table D37.** Accuracy of different village-level logistic model specifications explaining the use of each preventive measure in village SG3.

| | LLINs | Covering clothes | Boots | Gloves | Insecticide cream | Coils | Vaporisers | Burning materials |
|---|---|---|---|---|---|---|---|---|
| Empty model (A) | 0.96 | 0.63 | 0.89 | 0.98 | 0.74 | - | 0.85 | - |
| Individual model (A+B) | 0.96 | 0.66 | 0.89 | 0.98 | 0.75 | - | 0.85 | - |
| Opinion leader model (A+B+C) | 0.96 | 0.67 | 0.89 | 0.98 | 0.75 | - | 0.85 | - |
| Network size model (A+B+C+D) | 0.96 | 0.72 | 0.89 | 0.98 | 0.75 | - | 0.85 | - |
| Network exposure model (A+B+C+D+E) | 0.96 | 0.72 | 0.91 | 0.98 | 0.75 | - | 0.85 | - |
| Household exposure model (A+B+C+D+F) | 0.96 | 0.69 | 0.89 | 0.98 | 0.86 | - | 0.87 | - |
| Full model (A+B+C+D+E+F) | 0.96 | 0.69 | 0.90 | 0.98 | 0.86 | - | 0.87 | - |

**Table D38.** Accuracy of different village-level logistic model specifications explaining the use of each preventive measure in village SG4.

| | LLINs | Covering clothes | Boots | Gloves | Insecticide cream | Coils | Vaporisers | Burning materials |
|---|---|---|---|---|---|---|---|---|
| Empty model (A) | 0.94 | 0.51 | - | - | 0.94 | 0.85 | - | - |
| Individual model (A+B) | 0.94 | 0.62 | - | - | 0.95 | 0.86 | - | - |
| Opinion leader model (A+B+C) | 0.94 | 0.81 | - | - | 0.95 | 0.86 | - | - |
| Network size model (A+B+C+D) | 0.94 | 0.86 | - | - | 0.95 | 0.86 | - | - |
| Network exposure model (A+B+C+D+E) | 0.94 | 0.86 | - | - | 0.95 | 0.90 | - | - |
| Household exposure model (A+B+C+D+F) | 0.94 | 0.85 | - | - | 0.98 | 0.88 | - | - |
| Full model (A+B+C+D+E+F) | 0.92 | 0.85 | - | - | 0.98 | 0.92 | - | - |

#### Appendix E. Results of the analyses using Stochastic Actor-oriented Models (SAOMs)E.1 Overview of modelling approach

This section reports the village-level stationary stochastic actor oriented models (SAOMs), the SAOM meta-analysis reported in the main text, goodness of fit tests for village-level models, and robustness checks for the model rate parameters. In our SAOMs, we model the co-evolution (or rather: the joint short-term equilibrium) of two networks: one representing discussion ties between villagers and one representing the use of different measures by villagers. We used the RSiena package in R to fit the village-level models (Ripley et al., 2020), and the metafor package (Viechtbauer, 2010) to perform the meta-analysis of results.

The main network of interest is a two-mode network connecting villagers to preventive measures. The presence of a tie in this network means that a given villager reported using the measure in question, whereas a tie's absence means they did not. This way, we do not distinguish between preventive measures in the model, like we did in earlier analyses. This is not a shortcoming of the SAOM framework: we could study processes specific to certain measures by the use of nodal covariates. However, the advantage of assuming that social processes are comparable across measures is more statistical power – which is something we lacked in our logistic models presented earlier.

Our second modelled network is the one-mode discussion network about health-related issues connecting villagers. The presence of a tie here means that a given villager reported talking to the other villager in question; tie absence means a lack of such mention. This network is directed as it is possible that either or both members of a pair (dyad) of villagers recalls or finds a discussion relevant to mention.

The two-mode and one-mode networks jointly define a multilevel network of discussions and measure use in each village. These networks are visualised in Appendix C. We apply the SAOM to explore which social mechanisms may keep the structure of these multilevel networks stable over a short period of time. While changes in the discussion network are explicitly modelled in our approach, we focus on interpreting results about the use of measures as these are focal to our research questions. Results for the discussion network are viewed as "control" variables, which allow better estimation of parameters explaining the use of measures. We ensured nonetheless that the final models provided a reasonable representation of the discussion networks as well.

#### E.2 Model specification and definition of effects

The specification of effects in the fitted models was determined by our research questions (network size and exposure to use of measures), earlier empirical findings (role of individual characteristics and households), and established practices in the field of dynamic network modelling with SAOMs (see Ripley et al., 2020; Block, 2015). **Table E1** and **E2** list the effects that were included in the full models, for the use and the talk networks respectively, along with their "short names" based on which they can be exactly identified in section 12 of the RSiena Manual (Ripley et al., 2020). Effects 1-21 are identical to those presented in the main text.

Where relevant, linear effects of network statistics (as opposed weighted sums) were used by setting appropriate effect parameters to 1 in RSiena.

**Table E1**. Description of SAOM effects used for modelling the use of preventive measures network.

| *Effects of network structural variables* | | *Rsiena "short names"* |
|---|---|---|
| 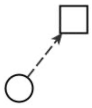 | (1) Outdegree (intercept) | density |
| 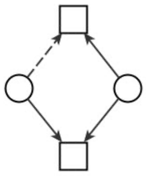 | (2) Agreement among villagers (4-cycles) x 100 | cycle4 |
| 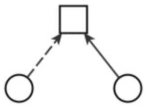 | (3) Popularity of measures | inPop |
| 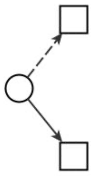 | (4) Activity of villagers | outAct |
| ***Effects of individual variables on villagers' use of measures*** | | ***Rsiena "short names"*** |
| 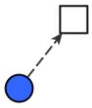 | (5) Female | egoX<br>(interaction: Female) |
| 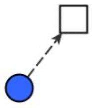 | (6) Head of household | egoX<br>(interaction: Head of Household) |
| 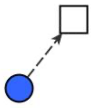 | (7) Carer for a sick person | egoX<br>(interaction: Carer for a sick person) |
| 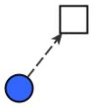 | (8) Works in fields | egoX<br>(interaction: Works in fields) |
| 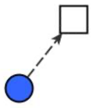 | (9) Age | egoX<br>(interaction: Age) |

| | | |
|---|---|---|
| 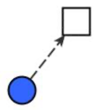 | (10) Education | egoX<br>(interaction: Education) |
| ***Effects of opinion leaders on villagers' use of measures*** | | ***Rsiena "short names"*** |
| 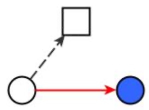 | (11) Talking to the Asha | egoX<br>(interaction: talks to Asha) |
| 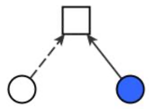 | (12) Asha's use of a specific measure | altX<br>(interaction Asha uses) |
| 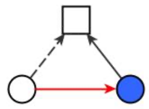 | (13) Asha's use of a measure if talking to Asha | egoX * altX<br>(interaction: talks to Asha, Asha uses) |
| 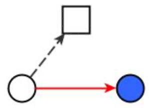 | (14) Talking to Healer | egoX<br>(interaction: talks to Healer) |
| 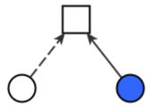 | (15) Healer's use of a specific measures | altX<br>(interaction Healer uses) |
| 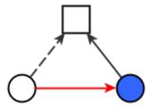 | (16) Healer's use of a measure if talking to Healer | egoX * altX<br>(interaction: talks to Healer, Healer uses) |
| ***Effects of talking to others on villagers' use of measures*** | | ***Rsiena "short names"*** |
| 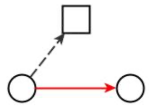 | (17) Number of other villagers one talks to | outActIntn<br>(interaction: talk) |
| 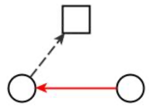 | (18) Number of other villagers who talk to one | inActIntn<br>(interaction: talk) |
| 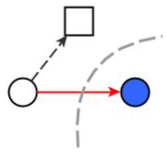 | (19) Number of non-villagers one talks to | egoX<br>(interaction: outdegree out of village) |
| ***Effects from exposure*** | ***Rsiena "short names"*** | |

| | | |
|---|---|---|
| 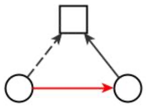 | (20) Using same measures as those one talks to | to<br>(interaction: talk) |
| 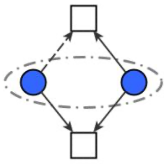 | (21) Using same measures as household members | sameXCycle4<br>(interaction: same household) |

**Table E2**. Description of SAOM effects used for modelling the health-related discussion network.

| *Effects of network structural variables* | | *Rsiena "short names"* |
|---|---|---|
| 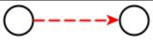 | (1) Outdegree (intercept) | density |
| 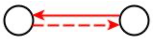 | (2) Reciprocity | recip |
| 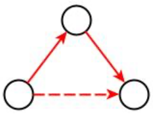 | (3) Transitivity | transTrip |
| 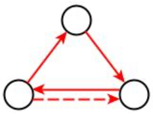 | (4) Transitivity * reciprocity | transRecTrip |
| 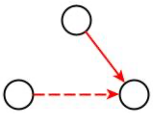 | (5) Popularity of villagers | inPop |
| 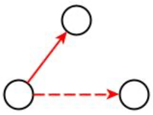 | (6) Activity of villagers | outAct |
| ***Effects of individual variables on discussion ties*** | | ***Rsiena "short names"*** |
| 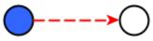 | (7) Sender Female | egoX<br>(interaction: Female) |
| 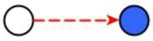 | (8) Receiver Female | altX<br>(interaction: Female) |
| 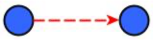 | (9) Both Female | sameX<br>(interaction: Female) |
| 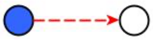 | (10) Sender Head of household | egoX<br>(interaction: Head of Household) |
| 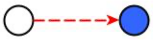 | (11) Receiver Head of household | altX<br>(interaction: Head of Household) |

| | | |
|---|---|---|
| 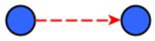 | (12) Both Head of household | sameX<br>(interaction: Head of Household) |
| 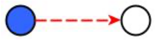 | (13) Sender carer for a sick person | egoX<br>(interaction: Carer for a sick person) |
| 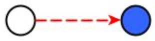 | (14) Receiver carer for a sick person | altX<br>(interaction: Carer for a sick person) |
| 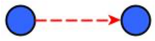 | (15) Both carer for a sick person | sameX<br>(interaction: Carer for a sick person) |
| 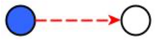 | (16) Sender Works in fields | egoX<br>(interaction: Works in fields) |
| 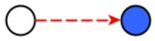 | (17) Receiver Works in fields | atX<br>(interaction: Works in fields) |
| 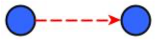 | (18) Both Work in fields | sameX<br>(interaction: Works in fields) |
| 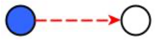 | (19) Sender Age | egoX<br>(interaction: Age) |
| 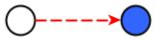 | (20) Receiver Age | altX<br>(interaction: Age) |
| 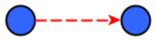 | (21) Both of similar Age | simX<br>(interaction: Age) |
| 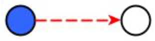 | (22) Sender Education | egoX<br>(interaction: Education) |
| 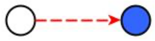 | (23) Receiver Education | altX<br>(interaction: Education) |
| 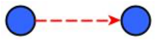 | (24) Both of similar Education | simX<br>(interaction: Education) |
| ***Effects of talking to opinion leaders on discussion ties*** | | ***Rsiena "short names"*** |
| 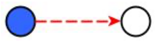 | (25) Sender Talks to the Asha | egoX<br>(interaction: talks to Asha) |

| | | |
|---|---|---|
| 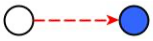 | (26) Receiver Talks to the Asha | altX<br>(interaction talks to Asha) |
| 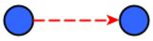 | (27) Both Talk to the Asha | sameX<br>(interaction: talks to Asha) |
| 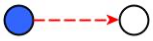 | (28) Sender Talks to the Healer | egoX<br>(interaction: talks to Healer) |
| 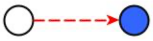 | (29) Receiver Talks to the Healer | altX<br>(interaction talks to Healer) |
| 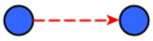 | (30) Both Talk to the Healer | sameX<br>(interaction: talks to Healer) |
| *Effects of household and people outside village on discussion ties* | | *Rsiena "short names"* |
| 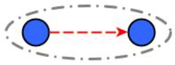 | (31) Both in same Household | sameX<br>(interaction: Household) |
| 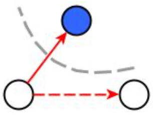 | (32) Sender's Number of non-villagers they talk to | egoX<br>(interaction: outdegree out of village) |
| 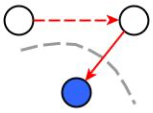 | (33) Receiver's Number of non-villagers they talk to | altX<br>(interaction: outdegree out of village) |
| 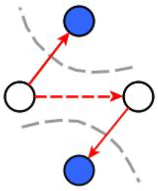 | (34) Similarity in Number of non-villagers they talk to | simX<br>(interaction: outdegree out of village) |
| *Effects from use of measures* | | *Rsiena "short names"* |
| 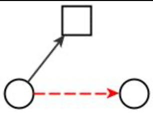 | (35) Sender's Number of measures used | outAct52<br>(interaction: use) |
| 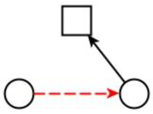 | (36) Receiver's Number of measures used | outPop<br>(interaction: use) |
| 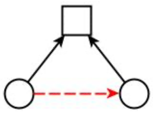 | (37) Both using the same measures | From<br>(interaction: use) |

#### E.3 Village-level SAOM results

We first attempted to fit a model with the above specification in each of the ten villages. In cases when models did not converge, we fixed parameters for effects with problematic estimates and tested these using score-type tests (Ripley et al., 2020). Effect fixing was done sequentially, fixing one additional effect at a time, until the model converged. Individual covariate ego, alter, and similarity effects were jointly tested. The score tests were all non-significant at a 5% level, with one exception: a mixed-network degree effect in village 10; since this effect was used to explain discussion ties, it is unlikely to affect the overall results on measure use. Further details about effect fixing and score-type tests are available from the authors.

Table E1 and E2 present the results of Stochastic Actor-oriented Models (SAOMs), one in each village. All of the presented models converged according to the criteria that all effect-wise convergence t-statistics are smaller than 0.1 in absolute value and the overall maximum convergence ratio is smaller than 0.25 (see Ripley et al., 2020). The models were estimated using rate parameters of 2.5 for both networks – this choice is further discussed below.
